# Non-enzymatic detection of SARS-CoV-2 RNA using DNA nanoswitches

**DOI:** 10.1101/2023.05.31.23290613

**Authors:** Javier Vilcapoma, Asmer Aliyeva, Andrew Hayden, Arun Richard Chandrasekaran, Lifeng Zhou, Jibin Abraham Punnoose, Darren Yang, Clinton H. Hansen, Simon Chi-Chin Shiu, Alexis Russell, Kirsten St. George, Wesley P. Wong, Ken Halvorsen

**Affiliations:** The RNA Institute, University at Albany, State University of New York, Albany, NY 12222; Department of Nanoscale Science and Engineering, University at Albany, State University of New York, Albany, NY 12222; Program in Cellular and Molecular Medicine, Boston Children’s Hospital, Boston, MA 02115; Wyss Institute for Biologically Inspired Engineering, Harvard University, Boston, MA 02115; Department of Biological Chemistry and Molecular Pharmacology, Blavatnik Institute, Harvard Medical School, Boston, MA 02115; Laboratory of Viral Diseases, Wadsworth Center, New York State Department of Health, Albany, NY 12208; Department of Biomedical Science, University at Albany, State University of New York, Albany, NY 12222

## Abstract

The emergence of a highly contagious novel coronavirus in 2019 led to an unprecedented need for large scale diagnostic testing. The associated challenges including reagent shortages, cost, deployment delays, and turnaround time have all highlighted the need for an alternative suite of low-cost tests. Here, we demonstrate a test for SARS-CoV-2 RNA that provides direct detection of viral RNA and eliminates the need for costly enzymes. We employ DNA nanoswitches that respond to segments of the viral RNA by a change in shape that is readable by gel electrophoresis. A new multi-targeting approach samples 120 different viral regions to improve the limit of detection and provide robust detection of viral variants. We apply our approach to a cohort of clinical samples, positively identifying a subset of samples with high viral loads. Since our method directly detects multiple regions of viral RNA without amplification, it eliminates the risk of amplicon contamination and renders the method less susceptible to false positives. This new tool can benefit diagnostic options for COVID-19 and future emerging outbreaks, providing a third option between amplification-based RNA detection and protein antigen detection. Ultimately, we believe this tool can be adapted both for low-resource onsite testing as well as for monitoring viral loads in recovering patients.

## Introduction

A novel coronavirus, SARS-CoV-2 (severe acute respiratory syndrome coronavirus 2), caused an outbreak of the disease COVID-19,[1,2] with over 600 million reported cases and 6.7 million deaths globally in the first three years of the pandemic.[3] The pandemic has highlighted the need for the development of rapid and low-cost alternate detection strategies for large scale and frequent testing.[4,5] The gold-standard method for viral detection in clinical samples is RT-qPCR (reverse transcription quantitative polymerase chain reaction), which involves nucleic acid amplification and its monitoring via fluorescent-labeled probes. The tests can be performed in a couple of hours, but logistical issues and the need for specialized laboratories makes typical turnaround times several days, creating a gap in preventing COVID positive individuals from transmitting the virus before the results are obtained. Antigen tests can be used to detect the various proteins of SARS-CoV-2 virions using lateral flow assay technology [5]. These tests have been developed and marketed as rapid home-based tests and are now widely used. They have less sensitivity than RT-qPCR tests and are commonly negative for two to three days following onset of symptoms, but provide results quicker and with less expense. At various points in the pandemic, large scale global testing strained supply chains and created backorders of reagents and consumables required for RT-qPCR assays with consequent bottlenecks in testing [6,7], or general scarcity of rapid antigen tests. These limitations in testing negatively affected our ability to control the pandemic. Further, in the acute phase of the pandemic, the frequency and speed of testing was found to be more practically relevant than test sensitivity for large scale COVID-19 testing.[8] Thus, the focus of new diagnostic assays shifted from solely high sensitivity to also include aspects of cost, turnaround time, and operation outside a lab by non-specialists.[9] These alternate testing strategies provide rapid results that aid in quick diagnosis and thus control of disease spread in contrast to highly sensitive tests with longer turnaround times.[10,11]

The challenges faced by traditional in vitro diagnostic techniques and the need for more frequent testing accelerated the development of alternative COVID-19 testing strategies [4,5]. Many new tests have been developed [12], most of which rely on RT-qPCR while a few use alternate approaches such as CRISPR [13,14] or isothermal amplification methods [15,16]. Several nanotechnology-based approaches have also been developed for detecting SARS-CoV-2 [17,18], including those based on nanoparticles [19], molecular switches [20], carbon nanotubes [21], graphene [22], and quantum dots [23]. These approaches all rely on enzymatic amplification for detection, or the use of proteins and antibodies for signal generation, which add cost and logistical challenges for transportation, storage, and use. To address some of these challenges, a few methods have been developed for non-enzymatic detection of viral RNA [24-26]. One promising approach for non-enzymatic detection of viral RNAs is with DNA nanotechnology, which allows for construction and dynamic control of nanoscale objects built from DNA [27,28]. Precise spatial positioning and programmable control over the shape of DNA nanostructures has been used extensively for many applications including biosensing and drug delivery [29]. For applications to viruses, DNA nanostructures have been employed to elucidate viral vaccine design principles [30] and in viral diagnostics [31] and therapeutics [32]. In our own work, we previously established proof of concept for detection of Zika virus RNA [33]. Here, we considerably expand on our prior work to develop a DNA nanoswitch-based assay for non-enzymatic, direct detection of SARS-CoV-2 by targeting multiple fragments of the SARS-CoV-2 genome.

## Results

### DNA nanoswitch design and operation

The DNA nanoswitch is constructed based on DNA origami principles. Typically, in DNA origami, a long single stranded DNA is folded into specific two- or three-dimensional shapes using short complementary strands [34]. Here, we create a simple linear duplex (the “off” state) using a 7249-nt long scaffold strand (commercially available M13 bacteriophage viral genome) tiled with short complementary backbone oligonucleotides (**Figure S1**) [35,36]. Two of the oligonucleotides contain single stranded extensions (detectors) that are complementary to parts of a target nucleic acid (**Figure 1a**). On binding the target sequence, the nanoswitch is reconfigured from the linear “off” state to a looped “on” state [37]. This conformational change is easily read out on an agarose gel where the on and off states of the nanoswitch migrate differently due to the topological difference (**Figure 1a**, inset). Importantly, this approach requires no complex equipment or enzymatic amplification. The signal comes from the intercalation of thousands of dye molecules from regularly used DNA gel stains (GelRed in this case), providing a high detection signal due to the length of the nanoswitch. In previous work, we used similar DNA nanoswitches for detection of microRNAs [38,39], viral RNAs [33], antigens [40] and enzymes [41].

**Figure 1.**
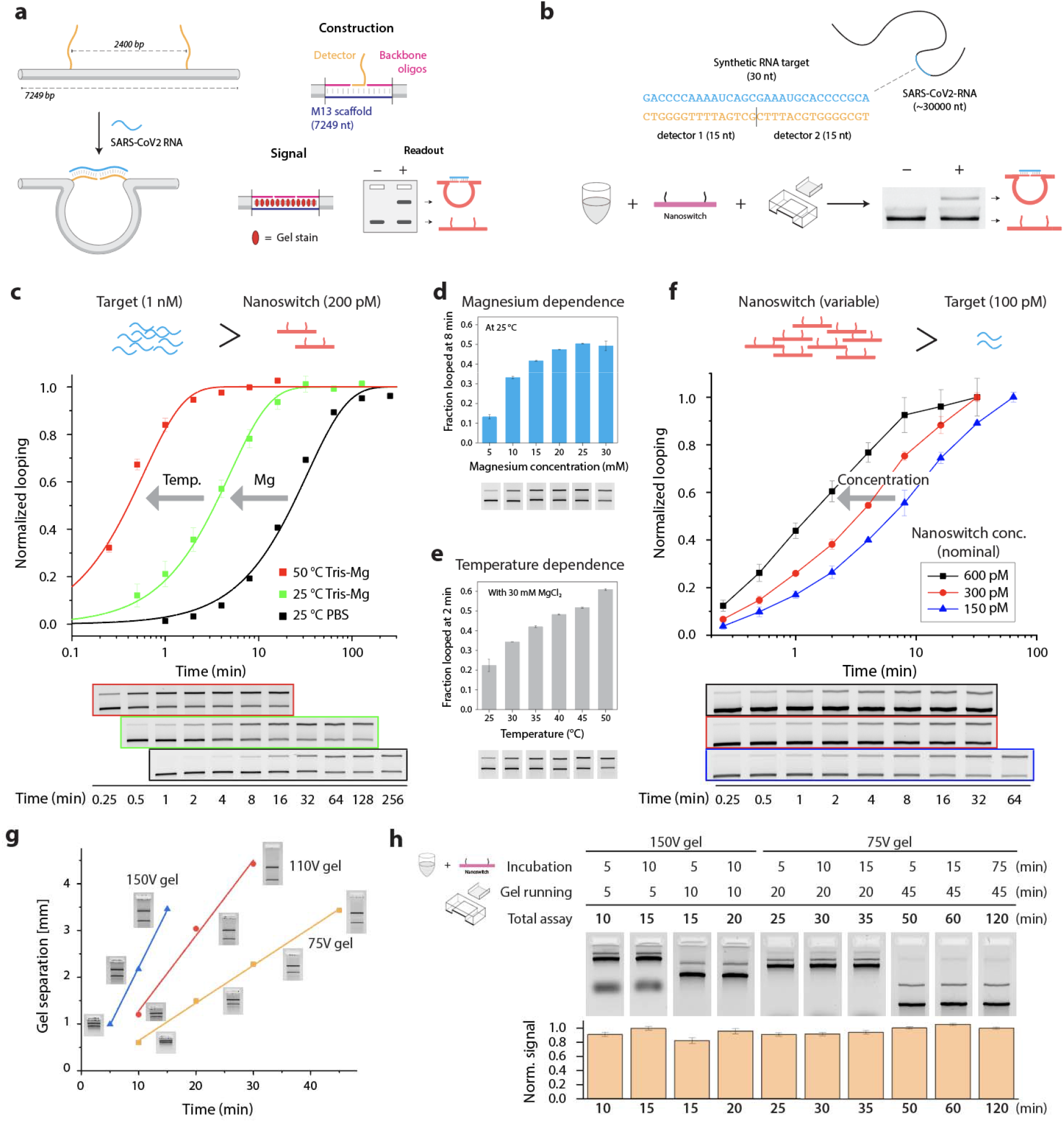
DNA nanoswitch detection of SARS-CoV-2 RNA. A) The DNA nanoswitch is a self-assembled DNA construct designed to form a loop upon interacting with a specific target sequence. The nanoswitch is stained with intercalating dye and imaged on a gel for detection readout. B) Design and validation of a DNA nanoswitch targeting a 30 nt portion of the N-gene. C) Reaction kinetics for a 30 nt RNA target in excess shows nearly two orders of magnitude improvement with optimal magnesium and temperature. D) Effect of various magnesium concentrations on room temperature kinetics. E) Effect of various temperatures on kinetics. F) Reaction kinetics with limited target. G) Gel separation of nanoswitch with different times and voltages. H) Overall assay time for a 50 pM target.

In this work, we considerably expand on our previous knowledge to accomplish direct, non-enzymatic detection of SARS-CoV-2 viral RNA. As a first step, we designed a nanoswitch with detectors that respond to a target sequence in SARS-CoV-2 that was expanded from one of the original CDC N2 primer targets [42]. Using a synthetic RNA target, we confirmed that our nanoswitches could detect this SARS-CoV-2 fragment (**Figure 1b**). Next, we moved to address one of the key challenges in detection of SARS-CoV-2 RNA: achieving a short detection time for a low concentration target. Previous nanoswitch work with proteins had shown rapid detection with a 30 minute incubation [40], but this goal had not yet been achieved for nucleic acids, where secondary structures can slow reaction kinetics. To address this issue, we screened the kinetics of the nanoswitch assay for a variety of conditions using a 30 nt synthetic RNA target that corresponds to a region in the SARS-CoV-2 genome nucleocapsid (N) gene (**Figure 1c**). First, we performed kinetic experiments at room temperature in phosphate buffered saline (PBS), which took nearly 2 hours to reach completion (**Figure 1c**). We then considered the addition of magnesium, which is known to facilitate nucleic acid hybridization [43,44]. Using a Tris-HCl buffer, we tested different concentrations of magnesium. We found that achieving the highest detection signal required at least 10 mM MgCl_2_ and that kinetics were enhanced up to 30 mM MgCl_2_ (**Figure 1d** and **Figure S2**). Choosing 30 mM MgCl_2_, we then measured kinetics at different temperatures, which showed a continued increase until ∼50°C (**Figure 1e** and **Figure S3**). With a 1 nM target RNA concentration, the magnesium and elevated temperature enabled complete reactions in less than 2 minutes, nearly two orders of magnitude faster than our starting condition (**Figure 1c**).

In a typical viral detection assay, we expect the target concentration (∼fM level) to generally be lower than the nanoswitch concentration (∼200-500 pM level). To assess more realistic conditions, we evaluated the kinetics for reactions with excess nanoswitch (**Figure 1f**). First, we confirmed that the rate of the signal accumulation relative to maximum signal is independent of the target concentration under these conditions (**Figure S4**). Next, we evaluated the kinetics of the reactions for 40 pM target RNA with varying nanoswitch concentrations. As expected, we found that increasing the nanoswitch concentration increased the reaction rate. At the highest nanoswitch concentration tested (∼600 pM), it took less than 2 minutes to reach half signal and less than 10 minutes to reach 90% signal (**Figure 1f**). To ensure a short end-to-end assay time, we additionally optimized the gel running conditions. We maximized band separation at a given voltage by minimizing buffer height above the gel (**Figure S5)**, which nearly doubled the separation. We then imaged looped nanoswitches at different voltages and running times, reducing the gel running time from 45 minutes to 5-20 minutes with minimal signal loss (**Figure 1g)**. These optimizations enabled detection of a 50 pM RNA target with an end-to-end assay time of as little as 10 minutes without sacrificing more than 20% of the signal (**Figure 1h**).

### Targeting multiple viral RNA fragments for signal enhancement

Next, we turned our attention to improving the analytical sensitivity of the assay to be suitable for viral targets. For SARS-CoV-2, the viral load can vary widely among patients from ∼10^3^ to 10^11^ copies/mL [45,46] due to various factors including viral variant, patient profile (age, health, vaccination status, e.g.), and time since infection [47]. Viral load is often considered a proxy for infectiousness, with most infectious individuals having excess of 10^6^ copies/mL and one study [45] reporting a median viral load as 10^6.8^ copies/mL (roughly 10 fM). Our previous sensitivity for small RNA targets such as microRNA has been reported in the ∼100 fM range. To improve sensitivity, we developed a new strategy for simultaneous targeting of different fragments of viral RNA. The concept is based on the idea that fragmenting the viral RNA produces many discrete targets from a single viral RNA genome (**Figure 2a**). A typical assay with a single target sequence can capture one target RNA per viral RNA, but an assay with multiple target sequences can capture multiple target RNAs per viral RNA to increase signal. In a nanoswitch assay, this can be accomplished by targeting several different SARS-CoV-2 sequences with identical loop sizes to essentially add the signal from multiple targets (**Figure 2b**). To achieve this, we developed a new nanoswitch design where multiple detectors that target distinct regions of the viral genome can be integrated onto a single nanoswitch, improving on previous ideas from combining individual single target nanoswitches [39].

**Figure 2.**
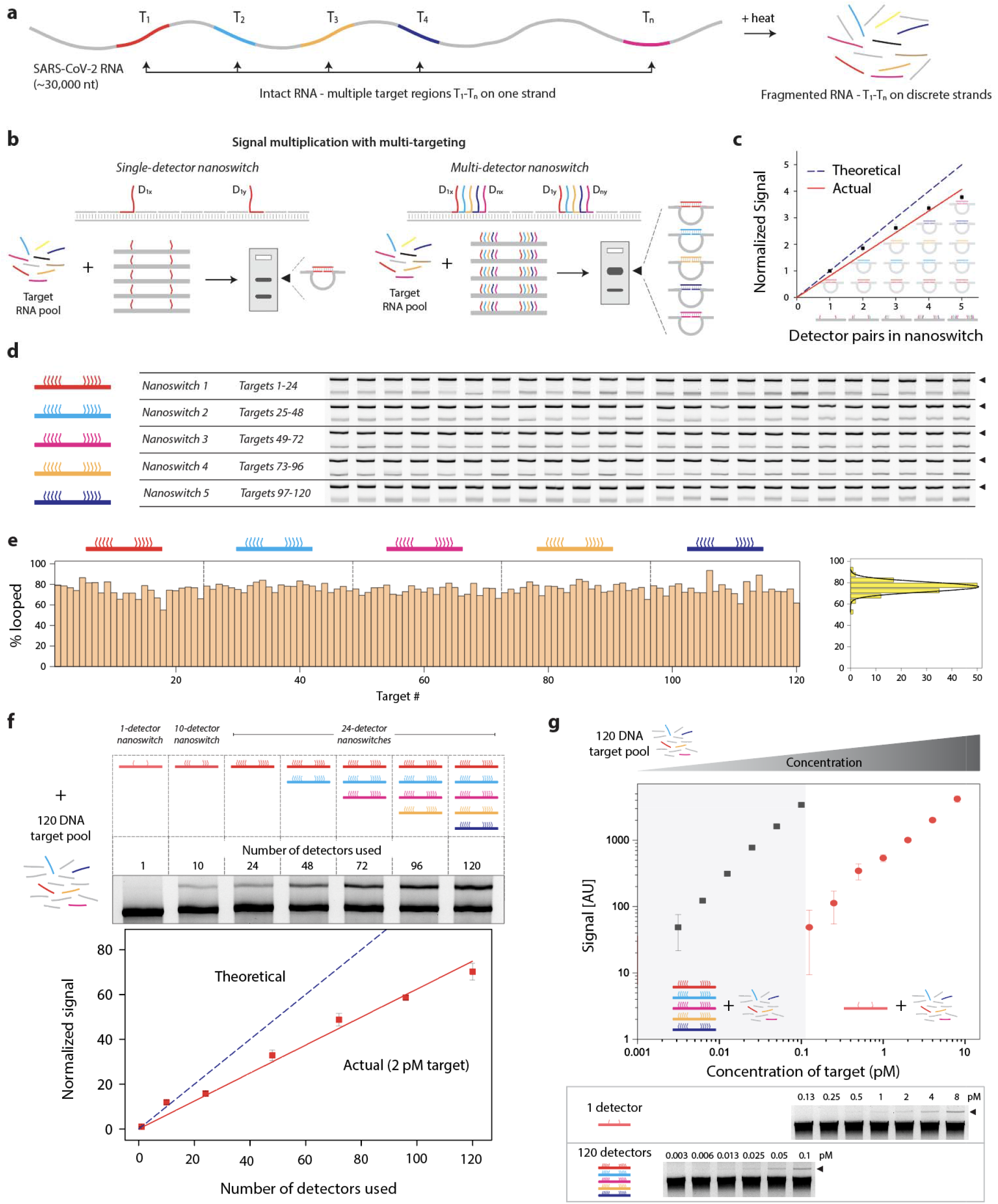
Improving sensitivity with multi-detector nanoswitches. A) Long viral RNA can have many target regions that separate into discrete strands after fragmentation. B) Concept of single-detector and multi-detector sensing. C) Validation of multi-detector sensing concept by targeting one to five sequences in a five-sequence pool. D) Development and validation of five 24 detector nanoswitches to enable targeting of 120 different regions. Gels demonstrate detection of each individual target. E) Quantified looped % of each nanoswitch target, with a histogram showing the distribution. F) Detection of a mock viral RNA using 120 equimolar targets and corresponding signal increase with increased detectors. G) Overall sensitivity of the 120 detector nanoswitch approach compared with single detector.

The detection signal for the nanoswitch assay is based on the looping of the nanoswitch when a target RNA binds a pair of detectors on the nanoswitch. To design a multi-detector nanoswitch, we incorporated pairwise detectors on the nanoswitch, with each pair offset along the length of the nanoswitch. We designed these pairwise detectors to be a fixed distance apart so that they all form the same loop size on binding their specific target (**Figure S1**). Under typical conditions for SARS-CoV-2 detection (when nanoswitch concentration >> target concentration), this results in the ability to detect multiple SARS-CoV-2 fragments with an additive signal (**Figure 2b)**. To implement this concept, we first identified loops that could be repositioned along the length of the nanoswitch with similar gel migrations. We screened different loop sizes and positions and found that a nearly centered loop of ∼2400 bp provided a constant signal when shifted (**Figure S6**). The identified design provided two 720 nt regions to place detector pairs. As a first proof of concept, we designed and built a five target nanoswitch to recognize five 30 nt regions of the SARS-CoV-2 genome. We confirmed that the nanoswitch responded to each of the five targets individually with an indistinguishable signal (**Figure S7**). We also verified that the kinetics of binding a single RNA target is identical between a multi-detector nanoswitch and a single target nanoswitch as expected (**Figure S8**). To demonstrate the signal multiplication concept, we used an equimolar pool of the 5 RNA targets representing a perfectly fragmented viral RNA and tested them against 5 different nanoswitches with different numbers of detectors that can target from 1 to all 5 of the targets. We show that for a constant target pool, we achieved a nearly stoichiometric increase in the signal as we increase the targeting capability (**Figure 2c**).

Having proven the concept, we sought to expand the number of targets. Considering the long length of viral RNAs such as SARS-CoV-2 (∼30,000 nt) and the short length required for detection (∼20-60 nt), this idea can in principle be expanded to hundreds of targets and achieve a corresponding level of signal amplification. We divided the 720 nt nanoswitch regions into 30 nt segments to enable development of 24-target nanoswitches. We designed and built five such 24-target nanoswitches to provide detection of up to 120 fragments that span the SARS-CoV-2 viral genome. We confirmed the positive detection of each of the 120 individual targets using DNA oligonucleotides (**Figure 2d**) and showed that maximum detection efficiency appears to be a gaussian distribution with a mean 76% (**Figure 2e)**. To measure the signal enhancement, we used an equimolar pool of the 120 targets and measured the detection signal using single nanoswitches with 1, 10, or 24 detector pairs and mixtures of 1, 2, 3, 4, and 5 24-target nanoswitches. We found that the signal increases nearly linearly with the number of detectors in the mixture (**Figure 2f**). Using serial dilutions of the 120-target mixture, we tested the sensitivity of the assay with the five multi-detector nanoswitch mixture and a single target nanoswitch. Compared to the LoD of 122 fM or 0.76 amol for a single target sequence, our signal multiplication strategy provides a ∼65-fold improvement to 1.9 fM or 0.01 amol, corresponding to ∼6,000 genome copies (**Figure 2g)**.

### Testing and validation with transcribed viral RNA

Key to our signal enhancement strategy is the ability to robustly fragment the long viral RNA. To develop and test fragmentation protocols, we first made and purified an in vitro transcribed (IVT) RNA of the SARS-CoV-2 N-gene (1279 nt). We tested three fragmentation reagents, a commercial buffer and two homemade buffers and settled on a homemade Tris-Magnesium reagent with 3 mM MgCl_2_ and pH 8.5 (**Figure S9**). Using our homemade fragmentation buffer, we fragmented the N-gene RNA for various lengths of time from 1 minute to ∼1 hour. We ran the products on a 2% agarose gel (**Figure 3b)** and estimated fragment sizes with a reference ladder. We found mean fragment sizes decreasing from ∼800 nt for 1 minute to ∼50 nt for 64 minutes with distribution widths also decreasing with fragmentation time (**Figure 3c)**.

**Figure 3.**
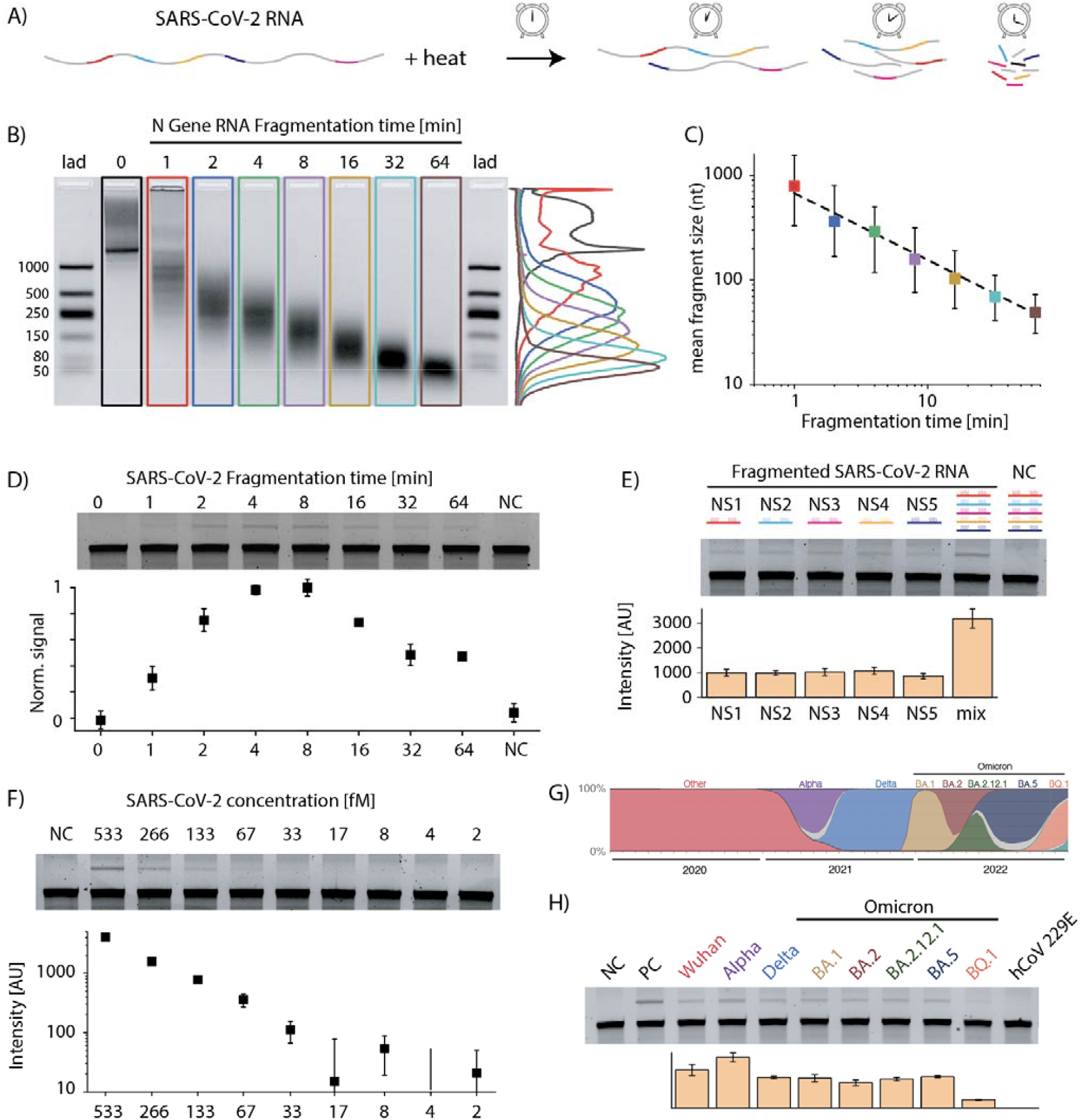
Detection of SARS-CoV-2 RNA. A) Scheme for SARS-CoV-2 fragmentation. B) In vitro transcribed N-gene RNA is fragmented using heat for various times with progressively smaller fragments over time. C) Distribution of apparent fragment sizes as a function of fragmentation time. D) Multi-detector nanoswitch detection of fragmented SARS-CoV-2 full genome RNA controls show optimal performance in the 2-16 minutes range. E) Detection of full genome SARS-CoV-2 with each of five 24 detector nanoswitches and the mixture of all five. F) Analytical sensitivity of the 120 multi-detector assay against full genome SARS-CoV-2 RNA. G) Prominence of different variants during the first 3 years of the pandemic in the United States (data from GISAID). H) Detection of major variants, and no detection of a non-SARS human coronavirus hCoV 229E.

Next we optimized nanoswitch design for the RNA fragments. Long RNAs including viral RNAs are known to contain strong secondary structures, some of which may persist even as the RNA is fragmented into smaller pieces in the 200 nt range [48,49]. Such structured regions could slow or otherwise impede binding to our DNA nanoswitch. During this project, evidence in our lab emerged that longer detector regions enhanced capture of a 401 nt RNA [50]. To see if that result applied to our fragmented viral RNA, we made and tested nanoswitches with detection regions of 15, 20, 25 and 30 nt and found that overall detection signal clearly increased with longer detector lengths (**Figure S10)**. This result was substantial enough to warrant redesign and reconstruction of our five 24 detector nanoswitches with the 30nt detector pairs. We redesigned our target regions and detectors for the new nanoswitches, which we again validated for each target (**Figure S11**). One unintended consequence of this change was false positive detection, likely due to weak cross-reactivity of detectors. These false positives were eliminated by post staining our gels rather than adding the stain to the samples, so we altered our protocol accordingly.

Having optimized our multi-targeting nanoswitches for fragmented N-gene RNA, we expanded the IVT detection to a more realistic target RNA, spanning the full SARS-CoV-2 genome across six strands. We fragmented the IVT RNA and found an optimal fragmentation time of 4-8 minutes based on detection of our five 24 detector nanoswitch mixture, balancing the quality of signal with our time requirements for a short test (**Figure 3d**). Choosing a fragmentation time of four minutes, we were able to detect the SARS-CoV-2 RNA with each of our five multi-detector nanoswitches in roughly equal proportion, with over a 3-fold increase when they were combined (**Figure 3e**). This result confirms good genome coverage for targeting different regions as well as a robust signal multiplication effect. Using this mixture of five 24-detector nanoswitches, we were able to achieve an LOD for this full SARS-CoV-2 genome equivalent of 33 fM or about 10^5^ copies (**Figure 3f**).

### Detection of viral RNA from SARS-CoV-2 variants

Throughout the pandemic, we were challenged with an evolving virus. Several dominant variants have emerged and been later replaced by others. In some cases, mutations in these variants have affected reliability of diagnostic tests [51,52]. To test the robustness of our nanoswitch assay, we obtained full genome RNA controls for the original (Wuhan) strain and each of the 6 other variants that have been dominant in the US during the first 3 years of the pandemic according to data from GISAID [53] **(Figure 3g**). We additionally obtained a non-SARS human coronavirus (hCoV229E) to test cross reactivity. Since our nanoswitch based assay covers the entire SARS-CoV-2 genome, our assay is uniquely indiscriminate to viral variants with different mutations. To demonstrate this, we used the five 24-detector nanoswitch mixture with fragmented RNA from the original strain as well as the six variants and the negative control (**Figure 3h**). We found that our nanoswitch assay was able to similarly detect all SARS-CoV-2 variants but exhibited no reactivity with hCoV229E. We believe that intensity variations in some variants was due to differing concentrations in the source material, which was supported by RT-qPCR data on the same material that showed nearly a 4-fold variation across samples, with BQ.1 having the lowest concentration (**Figure S12)**. Our unique approach of wide genome coverage makes our system robust against new variants.

### Detection of SARS-CoV-2 from clinical samples

To test our assay on clinical samples, we obtained 23 positive and 12 negative samples of RNA extracts from nasal or nasopharyngeal swabs (previously confirmed by RT-qPCR). Our optimized assay is suitable for use with RNA extraction protocols typically used for validation by RT-qPCR, with only an additional fragmentation step. We used 2 µL of RNA extract and fragmented the sample for 8 min, followed by incubation with the five 24-detector nanoswitch mixture. Our assay was able to confirm all 12 negatives and registered positive on 14 out of the 23 samples (**Figure 4 & Figure S13)**. The positive hits were correlated with the Ct values obtained from RT-qPCR, indicating a Ct threshold for our current assay in the mid to low 20s.

**Figure 4.**
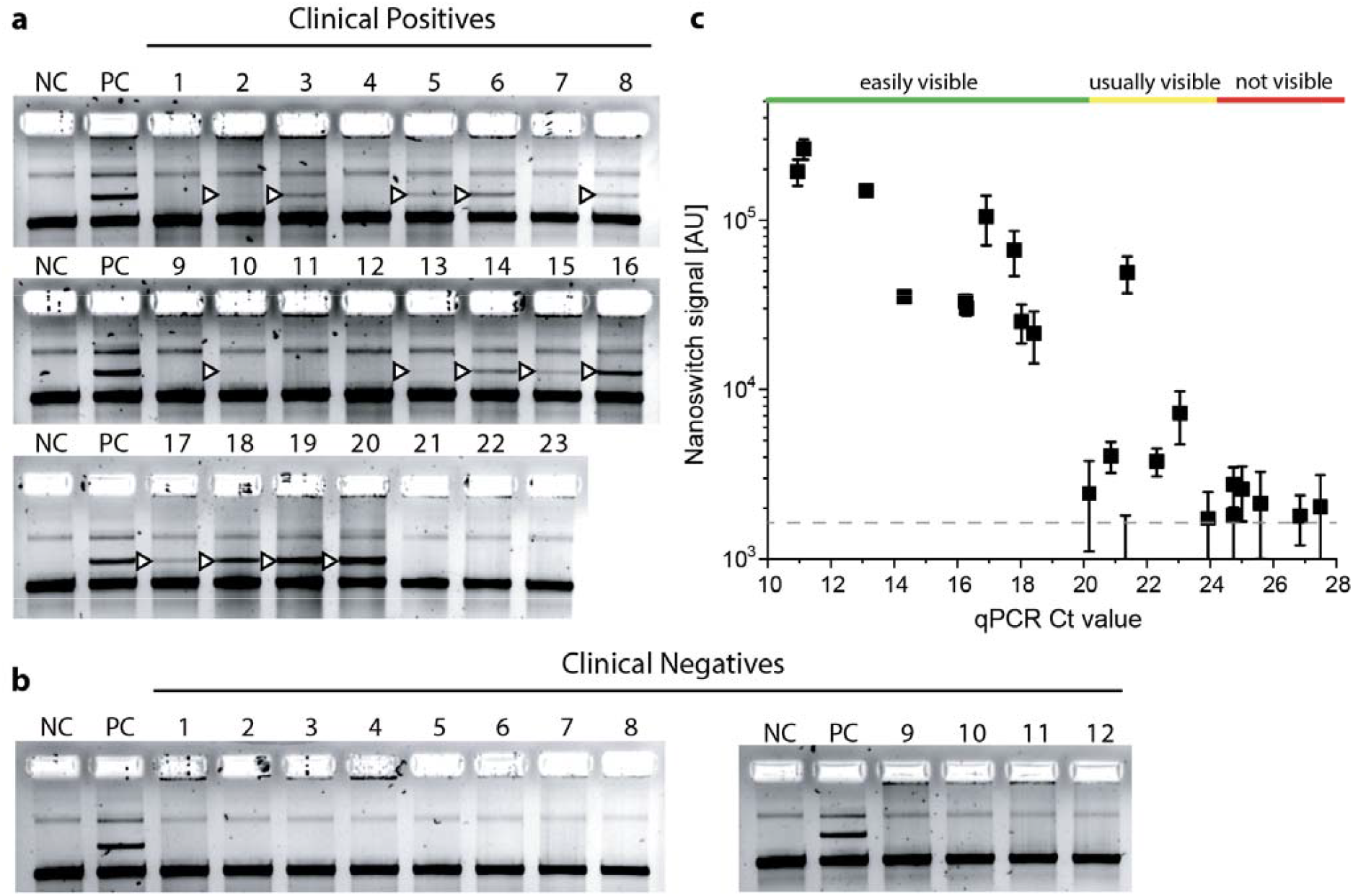
Detection of clinical SARS-CoV-2. A) Nanoswitch based detection of RNA extracts from clinical positive samples and clinical negative samples. B) Integrated band intensity vs Ct values obtained by qPCR. Error bars represent standard deviations from experimental triplicates. The dotted line represents the calculated limit of detection from the mean plus 3 standard deviations of triplicate negative controls.

## Discussion

Overall, our assay provides a unique non-enzymatic approach for direct detection of SARS-CoV-2 RNA. We have shown our method to be sensitive enough to capture a subset of positive clinical samples with moderate to high viral loads. The DNA nanoswitch approach has many advantages. The direct detection without enzymes eliminates high cost and complexity of enzymes including cold chain, and provides a direct linear response that is proportional to viral load, which has been shown to be related to infectivity [54] and severity of disease [55]. A related advantage that became obvious during the pandemic is that our assay uses different reagents than almost all other approaches, helping to diversify against supply shortages or bottlenecks.

Our multi-targeting approach provides coverage over large portions of the genome, which can enable detection even while variants continue to evolve. We have shown robust detection of the reference strain from Wuhan as well as more recent Alpha, Delta, and Omicron variants. This is a distinct advantage over PCR based methods, where some variants have affected the accuracy of certain diagnostic assays [51,52,56,57]. Alternatively, the flexible design of the nanoswitch and the detectors also allows options for designing for specificity between variants, especially in those cases where there are numerous mutations. This same versatility also enables our method to be quickly adapted in response to new diseases.

As it stands, we believe our assay could fill a niche somewhere between diagnostic RT-qPCR and rapid antigen tests. In comparison with RT-qPCR, our test is likely to be substantially less expensive with a potentially faster turnaround, but with lower analytical sensitivity more comparable to rapid antigen tests. However, it has been argued that frequent testing is more important than very high sensitivity in limiting SARS-CoV-2 spread [8]. There is also a clear pathway for improving our test sensitivity by processing more material with our nanoswitches. Currently, we are only testing about 1/1000^th^ of the collected sample (based on typical dilution of a swab into 1-3 mL of media and our use of ∼2 µL of extracted RNA) so a sensitivity increase of 10-100x could be readily achievable with different sample preparation. For example, the swab could be diluted in less initial volume, the RNA extraction volumes could be modified to process more volume and elute less volume, and more volume could be used in the final nanoswitch reaction. Compared to RT-qPCR, our method has another advantage of being more tolerant of certain contaminants that are known to affect RT-qPCR (i.e. salts and detergents) and being operable without the intense cleanliness that is required of RT-qPCR. Some RT-qPCR has been affected by false positives due to sample contamination [58]. It is also worth noting that our test performance improves with degraded RNA (to a point), a unique feature that may potentially reduce stringency of sample collection, transportation, and the testing environment. Compared to rapid antigen tests, our method has the potential to provide similar processing times and analytical sensitivity with similar or lower cost if developed into a commercial product.

This work represents the first demonstration of the DNA nanoswitches with clinical samples, and a first step toward diagnostic use for SARS-CoV-2. Further work includes making the readout more user-friendly, faster, and compatible for home or clinic use. To illustrate the potential for improvements, we demonstrated detection of a positive sample with high viral load in less than 20 minutes (**Figure S14**) Cartridge-based bufferless gels could be a simple way to further improve or streamline the process, and we have previously demonstrated that our test can be ported to the Invitrogen E-gel system with a cell phone camera.[40] Recent development in our lab of a 3D printed mini-gel system has shown resolution of DNA nanoswitches in as little as 2 minutes, in a format more compatible with point-of-care testing.[59] We believe optimization of gel percentage, running voltage, and dye type could improve this method further, bringing it closer to diagnostic use. Other systems such as automated electrophoresis could be also adapted for this purpose, or possibly capillary electrophoresis.

Our assay is also modular, enabling it to be quickly adapted in response to new diseases. Once the sequence information of a newly emerging virus becomes available, the detectors can be designed and synthesized within a day, and the nanoswitch assembly itself takes only a few hours. This plug-and-play design will be useful not just for viral RNA detection, but for a range of biomarkers, and even a combined nucleic acid/antigen test for one or more viral diseases. The nanoswitches can be stored dry or frozen without loss of stability or functionality [38], making the assay useful for potential on-site testing and easy distribution. Our assay is also non-enzymatic and does not require labeling of the target or the nanoswitches, making it low-cost and useful in low-resource settings.

Overall, our method provides an entirely new way to detect SARS-CoV-2 RNA that is in a class of its own for direct detection of RNA without enzymes. The COVID-19 pandemic illustrated how over-reliance on single types of tests can cause reagent shortages and workflow bottlenecks that can negatively impact disease control.[6] The minimalist nature of our approach requires relatively few and low-cost reagents, enabling the assay to be substantially cheaper than $1/test at scale, with the nanoswitch reagent itself costing less than 1 cent per reaction. We envision that this type of test could be adapted and distributed for frequent home testing or deployed for field surveillance testing. The programmability of the nanoswitches ensure that this method can be readily applied to other RNA viruses and make an impact on this and future disease outbreaks.

## Data Availability

All data produced in the present study are available upon reasonable request to the authors

## Author Contributions

J.V., A.A., A.H., L.Z., and S.C.S. performed experiments, aided in experimental design, and analyzed data. A.R.C. and J.A.P. designed and performed experiments, analyzed and visualized data and edited the manuscript. D.Y., and C.H. designed experiments, performed experiments and analyzed data. K.S.G. procured, selected, and provided deidentified clinical SARS-CoV-2 samples and edited the manuscript. A.R. retrieved archived clinical samples, performed RT-PCR to verify positive/negative status, and performed sequencing to determine lineage. W.P.W. supervised the project, designed experiments and edited the manuscript. K.H. conceived and supervised the project, designed experiments, analyzed and visualized data and wrote the manuscript.

## Acknowledgements

Research reported in this publication was supported by the National Science Foundation under award CBET2030279 to K.H. and W.P.W. and by the National Institutes of Health through the National Institute of General Medical Sciences under award R35GM124720 to K.H. and R35GM119537 to W.P.W. Funding for sequencing was provided by the New York Community Trust. We gratefully acknowledge all data contributors, i.e., the Authors and their Originating laboratories responsible for obtaining the specimens, and their Submitting laboratories for generating the genetic sequence and metadata and sharing via the GISAID Initiative, on which the variant plot in Figure 3g is based.

## Materials and Methods

### DNA nanoswitch design

Nanoswitches were designed with “detector” oligos that hybridize 30 nt to the M13 scaffold and 15-30 nt that protrude from the nanoswitch to hybridize to a portion of the target sequence. For single target nanoswitches, a pair of detectors form a sensing element and are spaced 2580 nt away from each other on the M13. The remainder of the M13 scaffold is tiled by 118 oligos of 60 nt length, 2 oligos of 30 nt length and 1 oligo of 49 nt length.

For multi-targeting nanoswitches, multiple pairs of detectors were positioned along the length such that each detector pair was separated by 2610 nt but separate pairs are offset by 30 nt. In this arrangement, the first detector pair occupies position 1921-1950 (with overhang at the 1950 position) and 4561-4590 (with overhang on the 4561 position) on the M13 scaffold. The second detector pair would be shifted by 30 nt pair (occupying 1951-1980 and 4591-4620) and the 24^th^ and final detector pair would occupy 2611-2640 and 5251-5280. Similarly to the single target nanoswitch, the remainder of the M13 is tiled by hybridizing oligonucleotides. For the 24 target nanoswitch, there are the 48 detector oligos which are mixed with 96 oligos of 60 nt length and 1 oligo of 49 nt length.

### Choosing SARS-CoV-2 targets

The first target in figure 1 was chosen based on an original CDC primer target [42], which was expanded to 30 nt. For the first set of multiple targets (from the Wuhan reference genome) with 30 nt length, we used the previously reported Matlab code [33] based on minimum distance between targets and minimal secondary structure. When we redesigned for 60 nt, we simplified the selection by searching for low predicted self folding in a 60 nt sliding window (> -8 kcal/mol) and then selected local areas of lowest predicted self-folding while maintaining at least 100 nt between adjacent targets. Our final 120 target regions have a mean inter-target distance of 250 nt with a minimum of 132 and a maximum of 418. The predicted self folding energies have a mean of -4.9 kcal/mol with a minimum of -8 kcal/mol and maximum of -0.6 kcal/mol.

### DNA nanoswitch construction

Construction of single target DNA nanoswitches have been extensively reported elsewhere [36-41], and we followed those methods here [60]. For multi-target nanoswitches, the construction protocol remained the same but the oligo mixes were altered to include the multiple sets of detection oligos. All oligos were mixed in equimolar ratios. Unless otherwise noted, all nanoswitches were LC purified using our previously established method [61].

### Experimental conditions

Reactions with DNA nanoswitches were typically carried out in 5 uL reaction volumes with 2 uL of LC purified nanoswitches (at ∼1 nM), 1 uL of 5x buffer, 1 uL of H_2_O, and 1 uL of target at 5x final concentration. The typical (1x) reaction buffer unless otherwise noted was 20 mM Tris-HCl, 30 mM MgCl_2_, 0.05% SDS, 1 uM blocking oligo. Samples were incubated for 2 hours at 50ºC unless otherwise noted. For nanoswitches with 15 nt detector regions (Figures 1 and 2), samples were prestained before running the gel with gels imaged immediately after. Samples were brought to 10 uL with water and added 2 uL of 6x loading dye and 1 uL of 10x GelRed (Biotium). For nanoswitches with 30 nt detector regions (Figures 3 and 4), GelRed was omitted from the sample and gels were post-stained in 1x GelRed. For IVT and clinical RNAs, fragmentation at 94ºC for 4-8 minutes (as noted) preceded the reaction. This was carried out in a 1x fragmentation buffer of 20 mM Tris-HCl, 3 mM MgCl_2_, 0.002 N NaOH (to adjust pH to 8.5).

### Gel electrophoresis

Gel electrophoresis was carried out using mini gel boxes (Owl Easy Cast B2) with 0.8% agarose (Sigma A9539) in 25 mL of 0.5x TBE. 5x TBE is made in house and diluted to 0.5x and purified through a 0.2 micron filter before use. Agarose solutions were brought to a boil on a hot plate and held for at least 30 seconds, before being removed from heat, cooled for a few minutes, poured and cured at room temperature for at least 45 minutes. Gels were run for 45 minutes at 75 volts at room temperature unless otherwise noted. Gels with pre-stained samples were imaged immediately using a gel documentation system (Bio-Rad GelDoc XR+ or Azure 400). Other gels were post-stained in a 1x solution of GelRed in water, with gentle shaking for at least 15 minutes. Gels were briefly destained and rinsed with water before imaging.

Gel analysis was performed with ImageLab for measuring the % looped or by ImageJ for quantifying weak signals. In ImageJ, images were first filtered with a 2 pixel median filter to reduce small bright speckles. A fixed size region of interest was applied to each lane to retrieve intensity profiles, which were quantified by peak analysis in OriginLab. For analysis of clinical samples, one replicate of a positive sample was excluded from analysis due to likely pipetting error (replicate one of sample 3). To establish the background signal value, we used three measurements of the negative control.

### Production of SARS-CoV-2 N gene RNA

In vitro transcribed SARS-CoV-2 nucleocapsid (N) gene RNA with a T7 promoter sequence and poly-A tail was prepared by the following method:

To provide sufficient cDNA input for in vitro transcription, N gene amplicon was prepared by PCR from a SARS-CoV-2 Positive Control (N gene) plasmid (NEB Cat No. N2117S). N gene PCR amplicons were purified with Qiagen QIAquick PCR kit following the manufacturer’s protocol with the following modifications: PCR product was eluted with 50 μL of Qiagen Buffer EB preheated to 60°C, allowed to soak on the column for 5 min., centrifuged at max speed (>10,000xg for 5 min.), then the eluate was reapplied and centrifuged again as above to maximize DNA recovery.

Purified N gene PCR product was quantified by Thermo NanoDrop 2000 UV spectrophotometer and analyzed by native 0.8% 0.5X TBE agarose gel electrophoresis post-stained with 1X GelRed to evaluate sizing and integrity.

IVT RNA was prepared from 100 ng of purified N gene PCR product as input using NEB HiScribe T7 Quick Yield RNA Synthesis Kit (NEB Cat. No. E2050S) following the manufacturer’s protocol with the following modifications: 1 μL of Sigma polyvinylsulfonic acid (PVSA) (900 μg/mL) was included as an RNase inhibitor in the 20 μL IVT RNA reaction. N gene IVT RNA was DNase I treated followed by lithium chloride precipitation and resuspended in nuclease-free water following NEB HiScribe T7 Quick Yield RNA Synthesis Kit protocol. Residual LiCl and enzymes were removed from IVT RNA using Zymo RNA Clean & ConcentratorTM-5 kit (Zymo Research, Cat. No. R1015) following the manufacturer’s protocol.

### Analysis of SARS-CoV-2 RNA controls

Eight different variants of IVT RNA controls for SARS-CoV-2 were purchased (Twist Bioscience). Controls #2, 14, 23, 48, 50, 62, 65, and 70 were used, representing common variants Wuhan, Alpha, Delta, BA.1, BA.2, BA.2.1, BA.5, and BQ.1, respectively. For qPCR analysis, we used the Luna SARS-CoV-2 RT-qPCR multiplex assay kit (New England Biolabs) and followed the standard protocol. All controls were diluted from their stocks to 800,000 copies/µL and fragmented for 4 minutes at 94°C in our 1x fragmentation buffer. The kit protocol was followed exactly, using 2 µL of test sample for each 20 µL reaction. The reaction was carried out on a MyGo mini qPCR machine, monitoring HEX and FAM for N1 and N2 regions, respectively. Data was analyzed using auto-quantification with the included software.

### Collection and RNA isolation from clinical Samples

Clinical samples of nasopharyngeal swabs had been previously received at the Virology Laboratory, Wadsworth Center, from numerous clinical sources throughout New York State for clinical COVID-19 testing. Processing and analysis had been performed as previously described [62]. Briefly extraction on the bioMerieux EMAG^®^ was followed by RT-PCR testing on the CDC 2019 nCoV Real-Time RT-PCR Diagnostic Panel. Positive and negative samples were selected, with positive samples across a range of Ct values, and samples were deidentified prior to use in the study. The use of clinical specimens for this study is approved under New York State DOH IRB study # 07-022.

## References

1. Wang, C., Horby, P. W., Hayden, F. G. & Gao, G. F. A novel coronavirus outbreak of global health concern. The Lancet 395, 470–473 (2020).

2. Zhu, N. et al. A Novel Coronavirus from Patients with Pneumonia in China, 2019. New England Journal of Medicine 382, 727–733 (2020).

3. WHO Coronavirus (COVID-19) Dashboard. https://covid19.who.int.

4. Udugama, B. et al. Diagnosing COVID-19: The Disease and Tools for Detection. ACS Nano 14, 3822–3835 (2020).

5. Peeling, Rosanna W., et al. “Diagnostics for COVID-19: moving from pandemic response to control.” The Lancet399.10326 (2022): 757–768.

6. Esbin, M. N. et al. Overcoming the bottleneck to widespread testing: A rapid review of nucleic acid testing approaches for COVID-19 detection. RNA rna.076232.120 (2020) doi:10.1261/rna.076232.120.

7. Behnam, Mohammad, et al. “COVID-19: overcoming supply shortages for diagnostic testing.” McKinsey & Company(2020).

8. Larremore, D. B. et al. Test sensitivity is secondary to frequency and turnaround time for COVID-19 screening. Science Advances 7, eabd5393.

9. Mina, M. J., Parker, R. & Larremore, D. B. Rethinking Covid-19 Test Sensitivity — A Strategy for Containment. New England Journal of Medicine 0, null (2020).

10. Toward COVID-19 Testing Any Time, Anywhere. The Scientist Magazine® https://www.the-scientist.com/news-opinion/toward-covid-19-testing-any-time-anywhere-67906.

11. Even imperfect Covid-19 tests can help control the pandemic. STAT https://www.statnews.com/2020/08/20/even-imperfect-covid-19-tests-can-help-control-the-pandemic/ (2020).

12. Rai, Praveen, et al. “Detection technologies and recent developments in the diagnosis of COVID-19 infection.” Applied microbiology and biotechnology 105 (2021): 441–455.

13. Broughton, James P., et al. “CRISPR–Cas12-based detection of SARS-CoV-2.” Nature biotechnology 38.7 (2020): 870–874.

14. Chandrasekaran, Sita S., et al. “Rapid detection of SARS-CoV-2 RNA in saliva via Cas13.” Nature Biomedical Engineering 6.8 (2022): 944–956.

15. Carter, Jake G., et al. “Ultrarapid detection of SARS-CoV-2 RNA using a reverse transcription– free exponential amplification reaction, RTF-EXPAR.” Proceedings of the National Academy of Sciences 118.35 (2021): e2100347118.

16. Woo, Chang Ha, et al. “Sensitive fluorescence detection of SARS-CoV-2 RNA in clinical samples via one-pot isothermal ligation and transcription.” Nature Biomedical Engineering4.12 (2020): 1168–1179.

17. Xu, Chun, et al. “Nanotechnology for the management of COVID-19 during the pandemic and in the post-pandemic era.” National Science Review 9.10 (2022): nwac124.

18. Weiss, Carsten, et al. “Toward nanotechnology-enabled approaches against the COVID-19 pandemic.” ACS nano 14.6 (2020): 6383–6406.

19. Moitra, P., Alafeef, M., Dighe, K., Frieman, M. B. & Pan, D. Selective Naked-Eye Detection of SARS-CoV-2 Mediated by N Gene Targeted Antisense Oligonucleotide Capped Plasmonic Nanoparticles. ACS Nano 14, 7617–7627 (2020).

20. Sundah, N. R. et al. Catalytic amplification by transition-state molecular switches for direct and sensitive detection of SARS-CoV-2. Science Advances 7, eabe5940 (2021).

21. Pinals, R. L. et al. Rapid SARS-CoV-2 Spike Protein Detection by Carbon Nanotube-Based Near-Infrared Nanosensors. Nano Lett. 21, 2272–2280 (2021).

22. Torrente-Rodríguez, R. M. et al. SARS-CoV-2 RapidPlex: A Graphene-Based Multiplexed Telemedicine Platform for Rapid and Low-Cost COVID-19 Diagnosis and Monitoring. Matter 3, 1981– 1998 (2020).

23. Li, C. et al. Synthesis of polystyrene-based fluorescent quantum dots nanolabel and its performance in H5N1 virus and SARS-CoV-2 antibody sensing. Talanta 225, 122064 (2021).

24. Mohammadniaei, Mohsen, et al. “A non-enzymatic, isothermal strand displacement and amplification assay for rapid detection of SARS-CoV-2 RNA.” Nature Communications 12.1 (2021): 5089.

25. Zhang, Jingjing, et al. “Non-enzymatic signal amplification-powered point-of-care SERS sensor for rapid and ultra-sensitive assay of SARS-CoV-2 RNA.” Biosensors and Bioelectronics 212 (2022): 114379.

26. Xue, Jian, et al. “Highly sensitive electrochemical aptasensor for SARS-CoV-2 antigen detection based on aptamer-binding induced multiple hairpin assembly signal amplification.” Talanta 248 (2022): 123605.

27. Seeman, Nadrian C. “DNA in a material world.” Nature421.6921 (2003): 427–431.

28. DeLuca, Marcello, et al. “Dynamic DNA nanotechnology: toward functional nanoscale devices.” Nanoscale Horizons 5.2 (2020): 182–201.

29. Chen, Yuan-Jyue, et al. “DNA nanotechnology from the test tube to the cell.” Nature nanotechnology 10.9 (2015): 748–760.

30. Veneziano, R. et al. Role of nanoscale antigen organization on B-cell activation probed using DNA origami. Nature Nanotechnology 15, 716–723 (2020).

31. Funck, Timon, et al. “Sensing picomolar concentrations of RNA using switchable plasmonic chirality.” Angewandte Chemie 130.41 (2018): 13683–13686.

32. Kwon, P. S. et al. Designer DNA architecture offers precise and multivalent spatial pattern-recognition for viral sensing and inhibition. Nature Chemistry 12, 26–35 (2020).

33. Zhou, Lifeng, et al. “Programmable low-cost DNA-based platform for viral RNA detection.” Science Advances 6.39 (2020): eabc6246.

34. Rothemund, P. W. K. Folding DNA to create nanoscale shapes and patterns. Nature 440, 297–302 (2006).

35. Halvorsen, Ken, Diane Schaak, and Wesley P. Wong. “Nanoengineering a single-molecule mechanical switch using DNA self-assembly.” Nanotechnology 22.49 (2011): 494005.

36. Koussa, Mounir A., et al. “DNA nanoswitches: a quantitative platform for gel-based biomolecular interaction analysis.” Nature methods 12.2 (2015): 123–126.

37. Chandrasekaran, A. R., Zavala, J. & Halvorsen, K. Programmable DNA Nanoswitches for Detection of Nucleic Acid Sequences. ACS Sens. 1, 120–123 (2016).

38. Chandrasekaran, A. R. et al. Cellular microRNA detection with miRacles: microRNA-activated conditional looping of engineered switches. Science Advances 5, eaau9443 (2019).

39. Chandrasekaran, Arun Richard, and Ken Halvorsen. “DNA-based smart reagent for detecting Alzheimer’s associated MicroRNAs.” ACS sensors 6.9 (2021): 3176–3181.

40. Hansen, C. H., Yang, D., Koussa, M. A. & Wong, W. P. Nanoswitch-linked immunosorbent assay (NLISA) for fast, sensitive, and specific protein detection. PNAS 114, 10367–10372 (2017).

41. Chandrasekaran, A. R., Trivedi, R. & Halvorsen, K. Ribonuclease-Responsive DNA Nanoswitches. Cell Reports Physical Science 1, 100117 (2020).

42. Lu, Xiaoyan, et al. “US CDC real-time reverse transcription PCR panel for detection of severe acute respiratory syndrome coronavirus 2.” Emerging infectious diseases 26.8 (2020): 1654.

43. Misra, Vinod K., and David E. Draper. “On the role of magnesium ions in RNA stability.” Biopolymers: Original Research on Biomolecules 48.2-3 (1998):113–135.

44. Špringer, Tomáš, et al. “Shielding effect of monovalent and divalent cations on solid-phase DNA hybridization: surface plasmon resonance biosensor study.” Nucleic acids research 38.20 (2010): 7343–7351.

45. Jacot, Damien, et al. “Viral load of SARS-CoV-2 across patients and compared to other respiratory viruses.” Microbes and infection 22.10 (2020): 617–621.

46. Pan, Yang, et al. “Viral load of SARS-CoV-2 in clinical samples.” The Lancet infectious diseases 20.4 (2020): 411–412.

47. Puhach, Olha, Benjamin Meyer, and Isabella Eckerle. “SARS-CoV-2 viral load and shedding kinetics.” Nature Reviews Microbiology 21.3 (2023): 147–161.

48. Boerneke, Mark A., Jeffrey E. Ehrhardt, and Kevin M. Weeks. “Physical and functional analysis of viral RNA genomes by SHAPE.” Annual review of virology 6 (2019): 93–117.

49. Jaafar, Zane A., and Jeffrey S. Kieft. “Viral RNA structure-based strategies to manipulate translation.” Nature Reviews Microbiology 17.2 (2019): 110–123.

50. Zhou, Lifeng, et al. “Sequence-selective purification of biological RNAs using DNA nanoswitches.” Cell reports methods 1.8 (2021): 100126.

51. Laine, Pia, et al. “SARS-CoV-2 variant with mutations in N gene affecting detection by widely used PCR primers.” Journal of Medical Virology 94.3 (2022): 1227–1231.

52. Chen, Yuqing, et al. “Impact of SARS-CoV-2 variants on the analytical sensitivity of rRT-PCR assays.” Journal of Clinical Microbiology 60.4 (2022): e02374–21.

53. Khare, Shruti, et al. “GISAID’s role in pandemic response.” China CDC weekly 3.49 (2021): 1049.

54. He, X. et al. Temporal dynamics in viral shedding and transmissibility of COVID-19. Nature Medicine 26, 672–675 (2020).

55. Pujadas, E. et al. SARS-CoV-2 viral load predicts COVID-19 mortality. The Lancet Respiratory Medicine 8, e70 (2020).

56. Jian, Ming-Jr, et al. “SARS-CoV-2 variants with T135I nucleocapsid mutations may affect antigen test performance.” International Journal of Infectious Diseases 114 (2022): 112–114.

57. Del Vecchio, Claudia, et al. “Impact of antigen test target failure and testing strategies on the transmission of SARS-CoV-2 variants.” Nature Communications 13.1 (2022): 5870.

58. Braunstein, G. D., Schwartz, L., Hymel, P. & Fielding, J. False Positive Results With SARS-CoV-2 RT-PCR Tests and How to Evaluate a RT-PCR-Positive Test for the Possibility of a False Positive Result. Journal of Occupational and Environmental Medicine 63, e159 (2021).

59. Morya, V., Hayden, A., Zhou, L., Cole, D., and Halvorsen, K. A 3D printed mini-gel electrophoresis system for rapid and inexpensive DNA nanoswitch biosensing. bioRxiv 10.64898/2026.01.21.700818.

60. Chandrasekaran, Arun Richard, Bijan K. Dey, and Ken Halvorsen. “How to Perform miRacles: A Step-by-Step microRNA Detection Protocol Using DNA Nanoswitches.” Current protocols in molecular biology 130.1 (2020): e114.

61. Halvorsen, Ken, et al. “Shear dependent LC purification of an engineered DNA nanoswitch and implications for DNA origami.” Analytical chemistry 89.11 (2017): 5673–5677.

62. Griesemer, Sara B., Greta Van Slyke, and Kirsten St. George. “Assessment of sample pooling for clinical SARS-CoV-2 testing.” Journal of Clinical Microbiology 59, no. 4 (2021): 10–1128.

