## Supplemental information for "Non-enzymatic detection of SARS-CoV-2 RNA using DNA nanoswitches"

Lifeng Zhou: Department of Advanced Manufacturing and Robotics, College of Engineering, Peking University, Beijing 100871, China

Simon Chi-Chin Shiu: School of Biomedical Sciences, Li Ka Shing Faculty of Medicine, The University of Hong Kong, Hong Kong, China

**Figure S1:**	Schematic of the single and multi-detector DNA nanoswitch

**Figure S2:**	Looping kinetics as a function of magnesium concentration

**Figure S3:**	Looping kinetics as a function of temperature

**Figure S4:**	Looping kinetics as a function of target concentration

**Figure S5:**	Nanoswitch resolution with different buffer volumes

**Figure S6:**	Screening loop positions for multi-detector nanoswitch

**Figure S7:**	Testing individual targets with the five detector nanoswitch

**Figure S8:**	Single target looping kinetics using single or multi-detector nanoswitch

**Figure S9:**	Testing different buffers for RNA fragmentation

**Figure S10:**	Testing IVT RNA detection with different detector lengths

**Figure S11:**	Validating individual targets for the 30nt multi-detector nanoswitches

**Figure S12:**	Figure S12 – qPCR analysis of SARS-CoV-2 IVT controls

**Figure S13:**	Triplicate experiments for clinical samples

**Figure S14:**	Rapid detection of high clinical positive

**Table S1:**	Target oligo sequences

**Table S2:**	Detector oligo sequences

**Table S3:**	Tiling oligo sequences

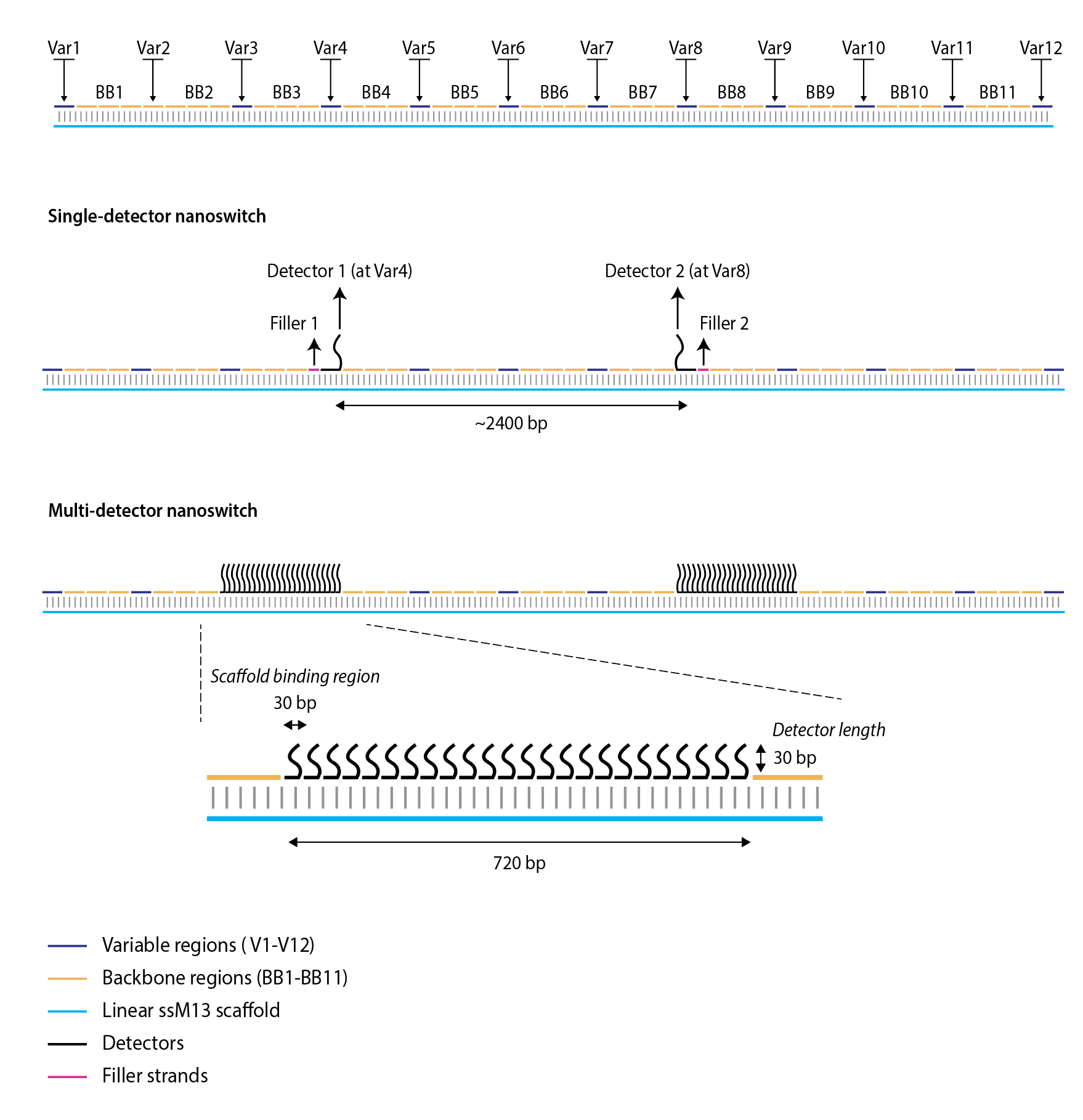

***Figure S1 – Schematic of the single and multi-detector DNA nanoswitch*.**

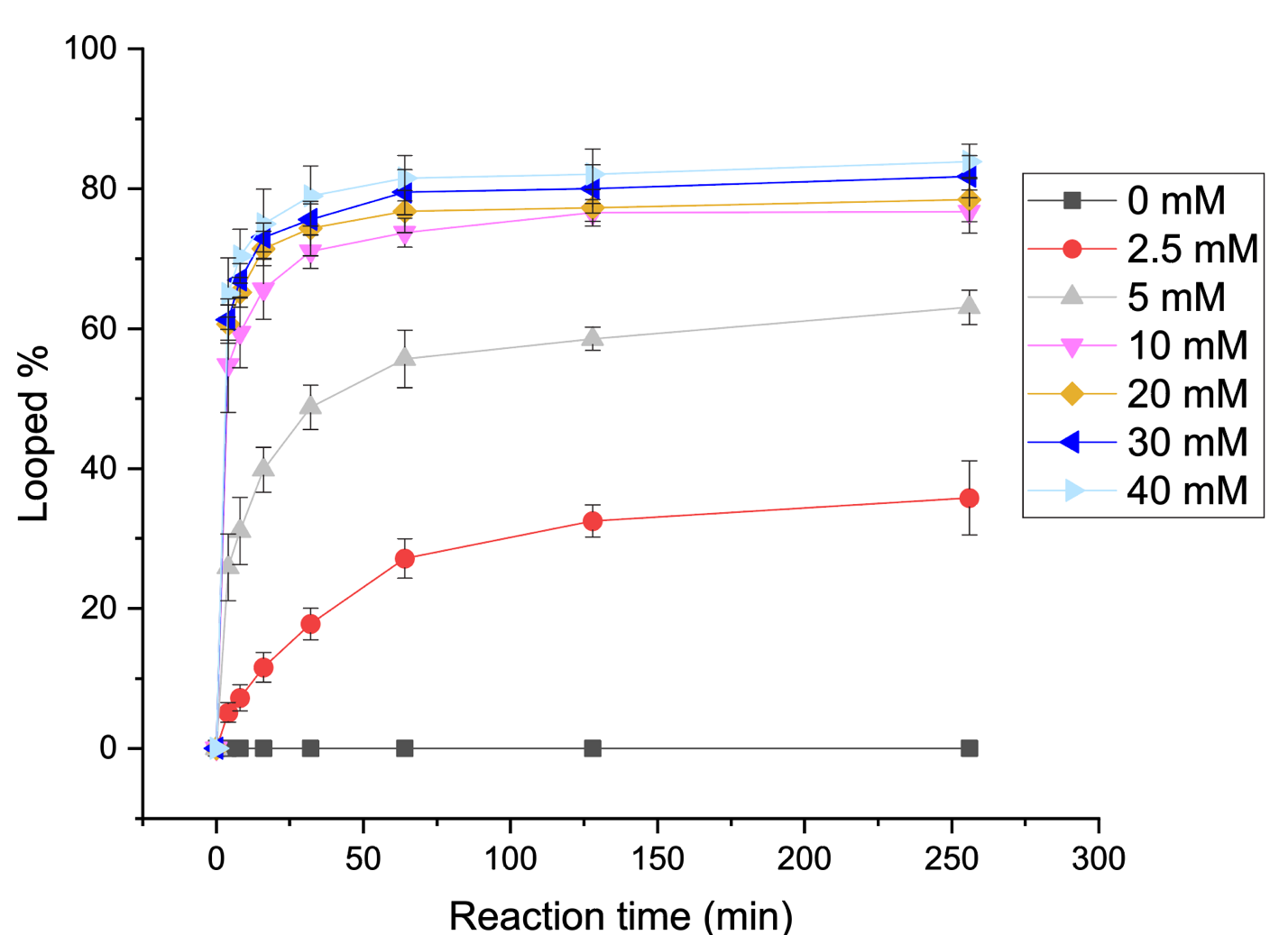

***Figure S2 – Looping kinetics as a function of magnesium concentration.***

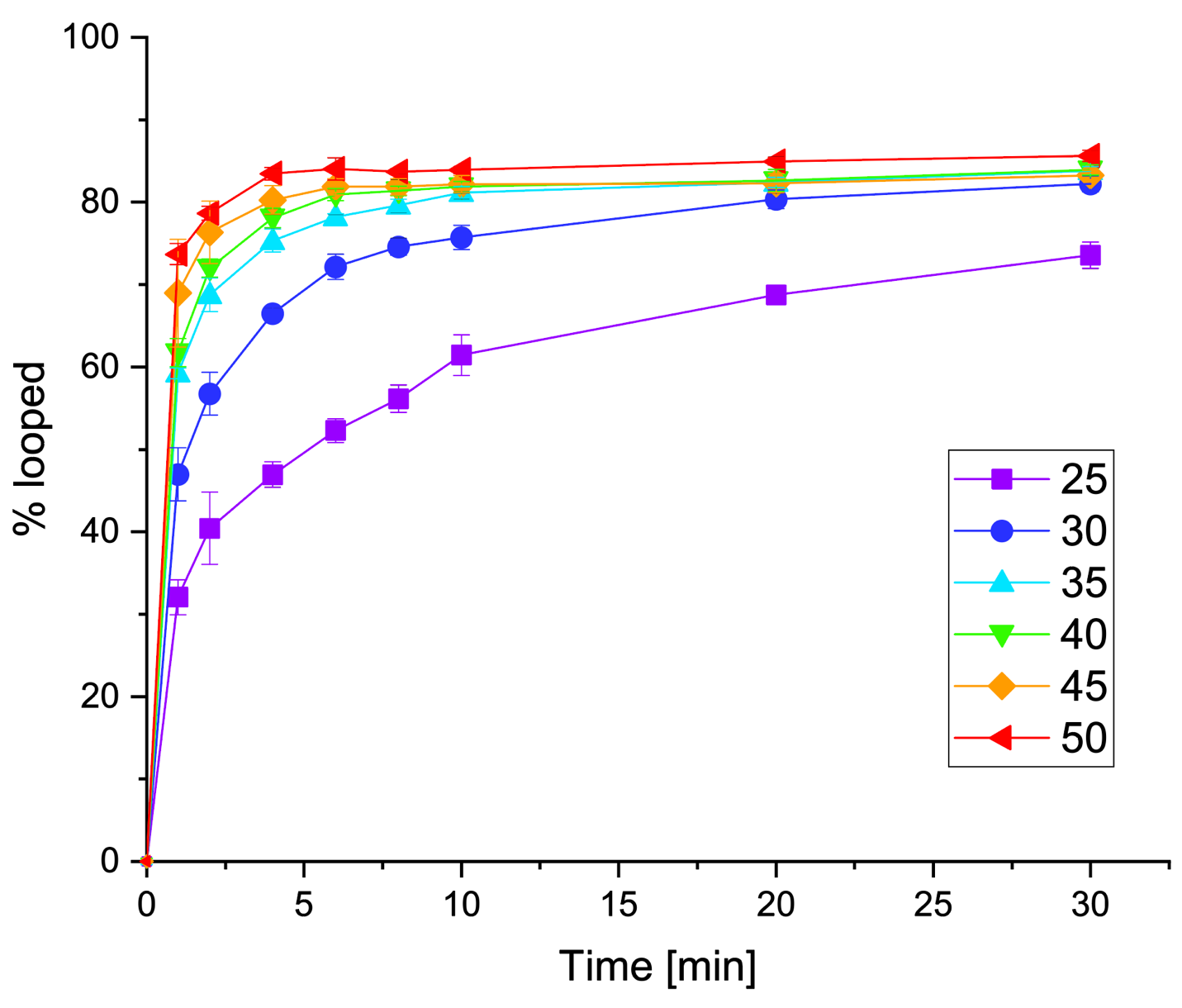

***Figure S3 – Looping kinetics as a function of temperature***

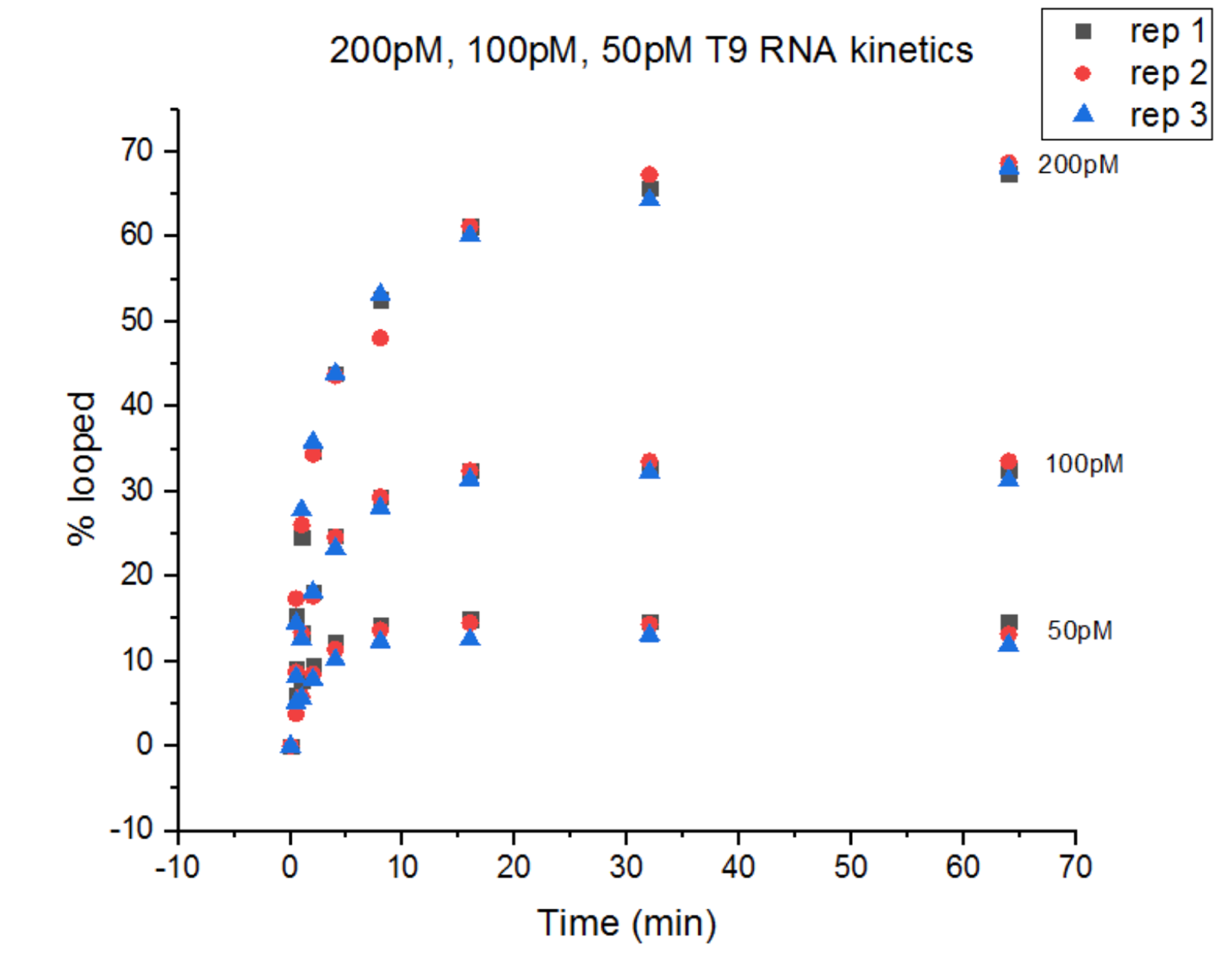

***Figure S4 – Looping kinetics as a function of target concentration***

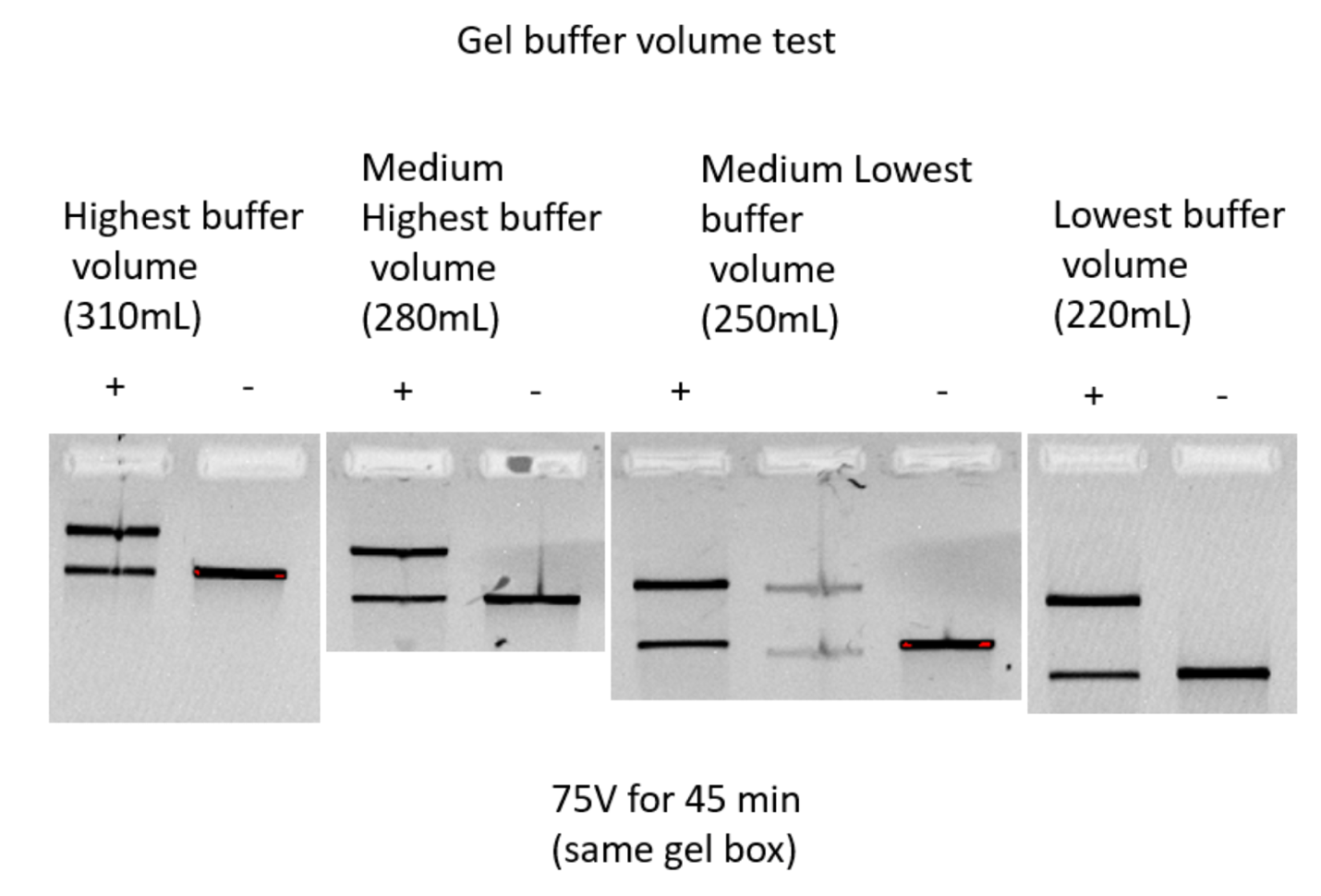

***Figure S5 – Nanoswitch resolution with different buffer volumes***

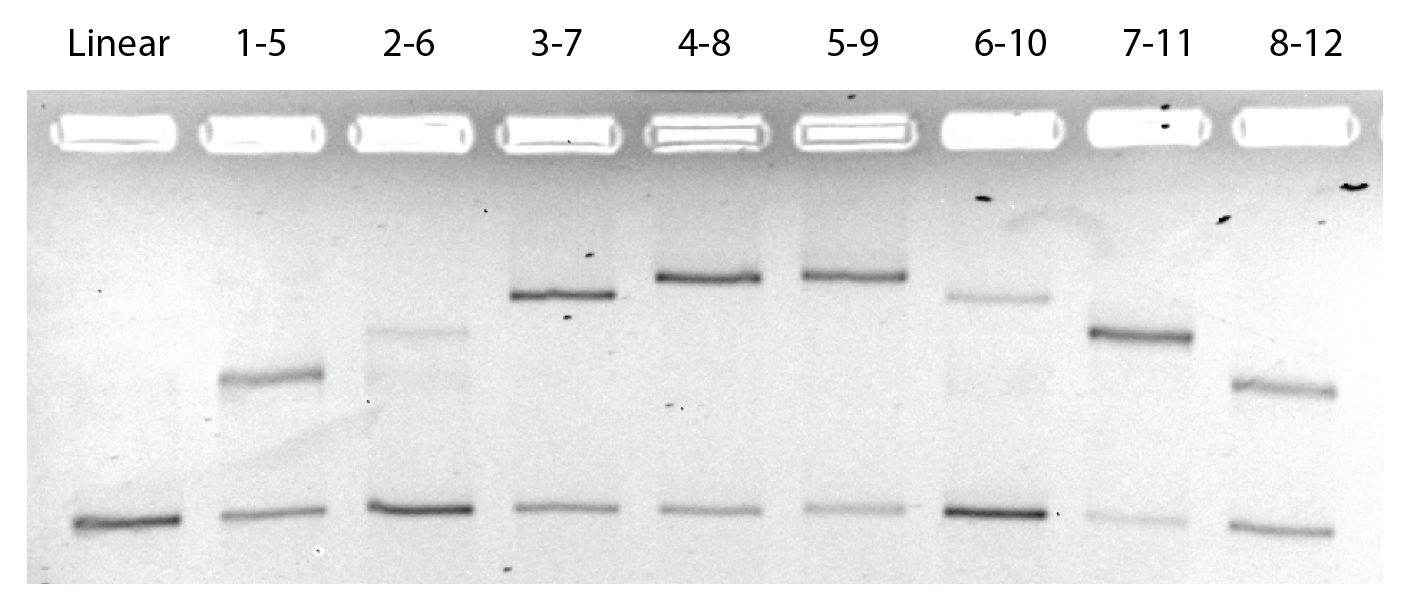

***Figure S6 – Screening loop positions for multi-detector nanoswitch.***

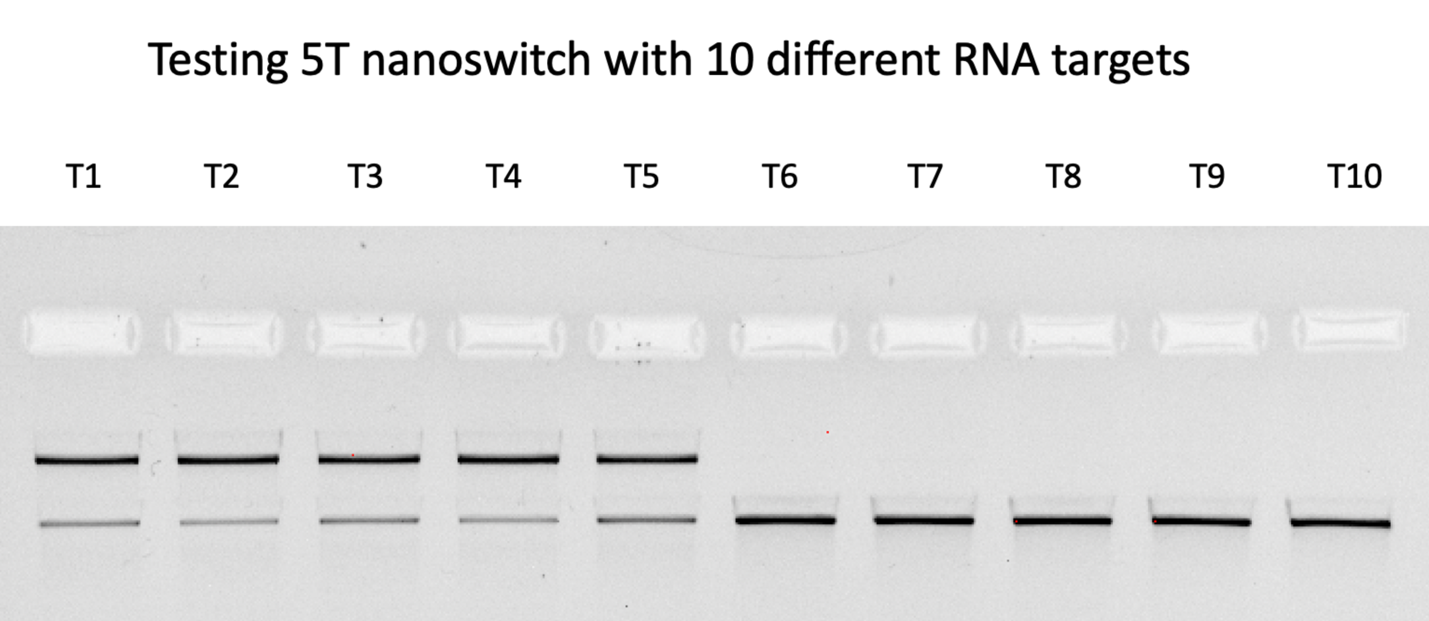

***Figure S7 – Testing individual targets with the five detector nanoswitch.*** Targets T1 through T5 are detected, while off targets T6 through T10 are not.

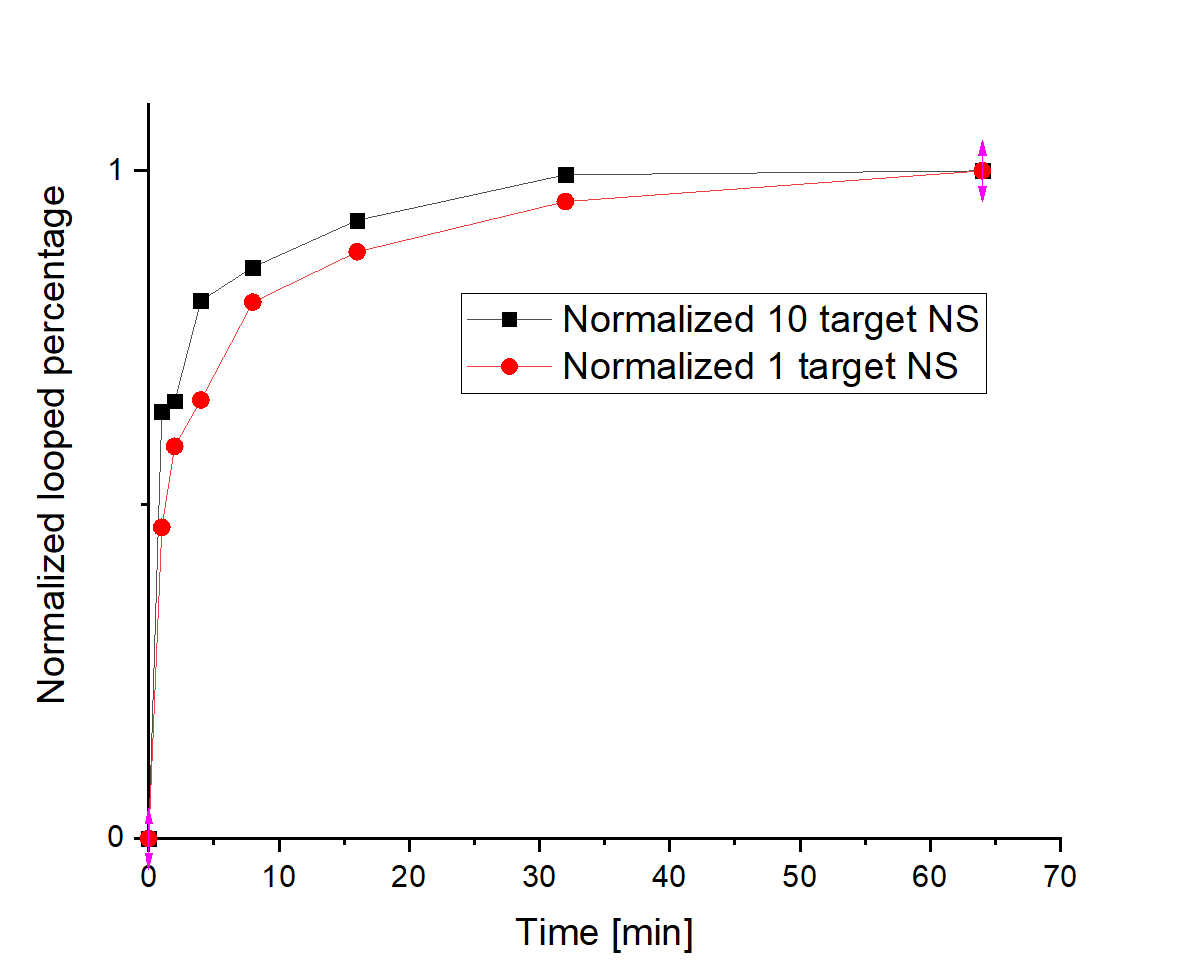

***Figure S8 – Single target looping kinetics using single or multi-detector nanoswitch.***

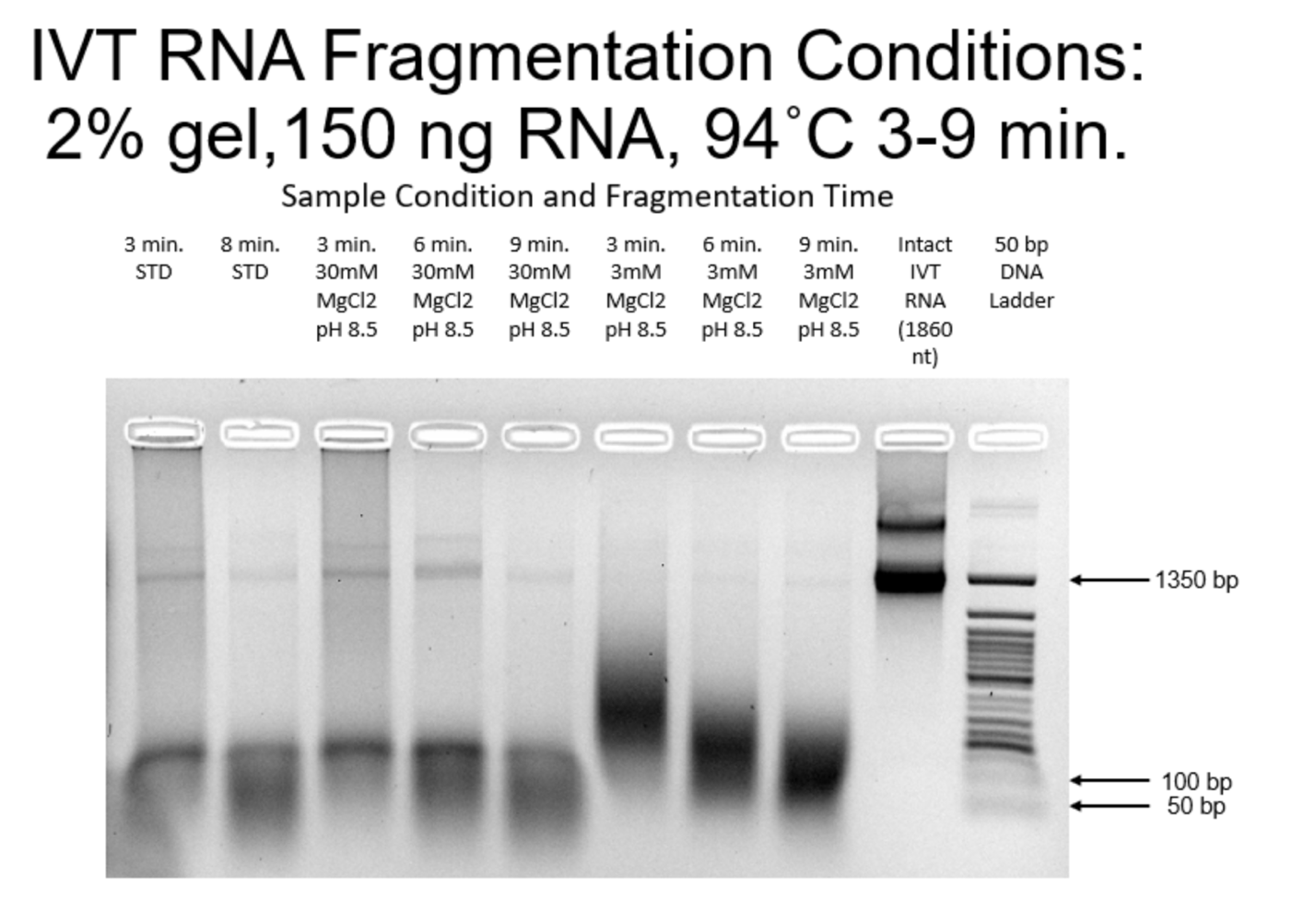

***Figure S9 – Testing different buffers for RNA fragmentation***

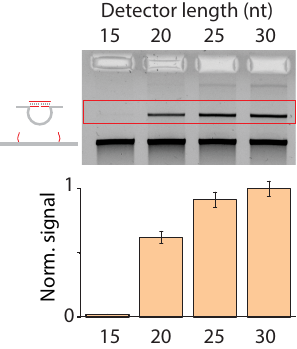

***Figure S10 – Testing IVT RNA detection with different detector lengths***

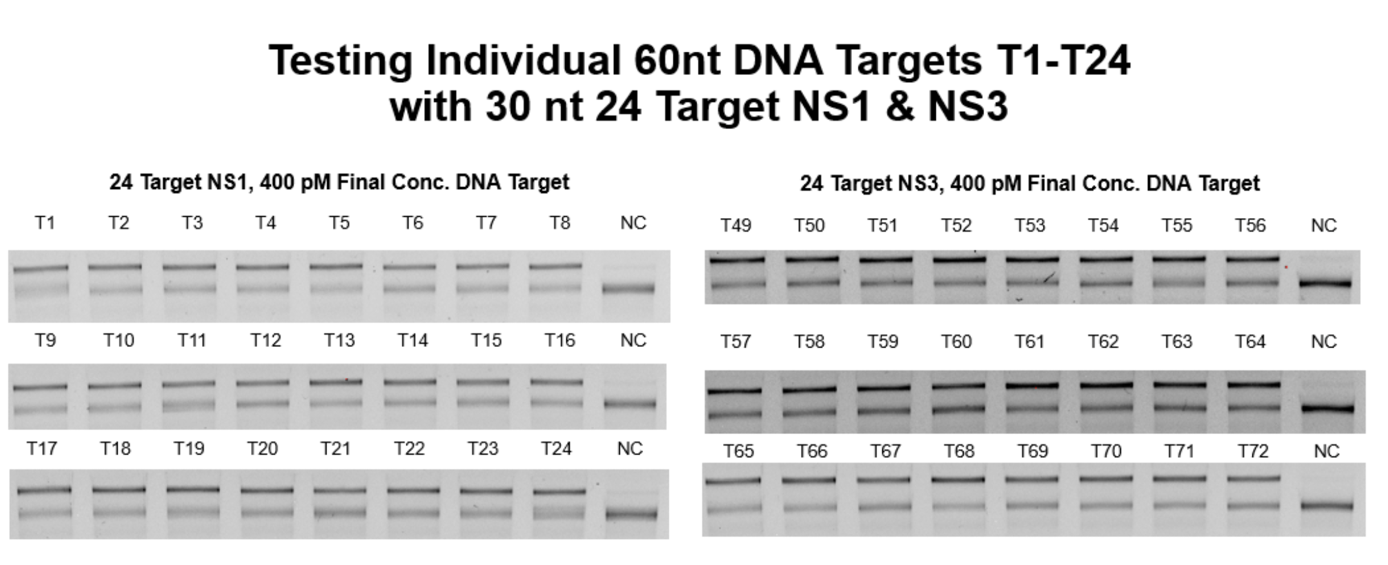

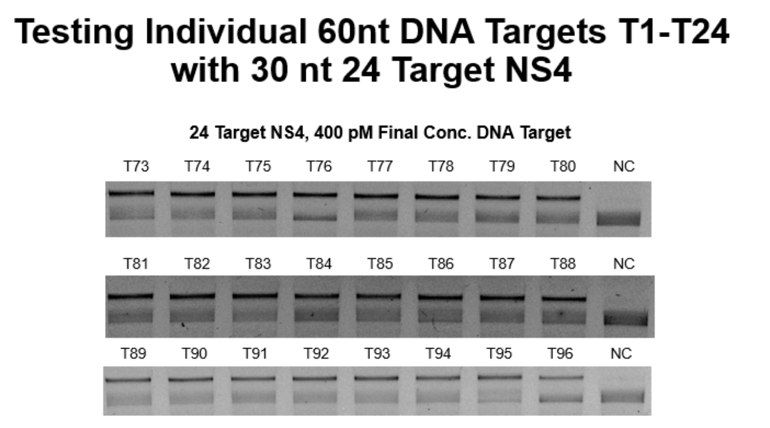

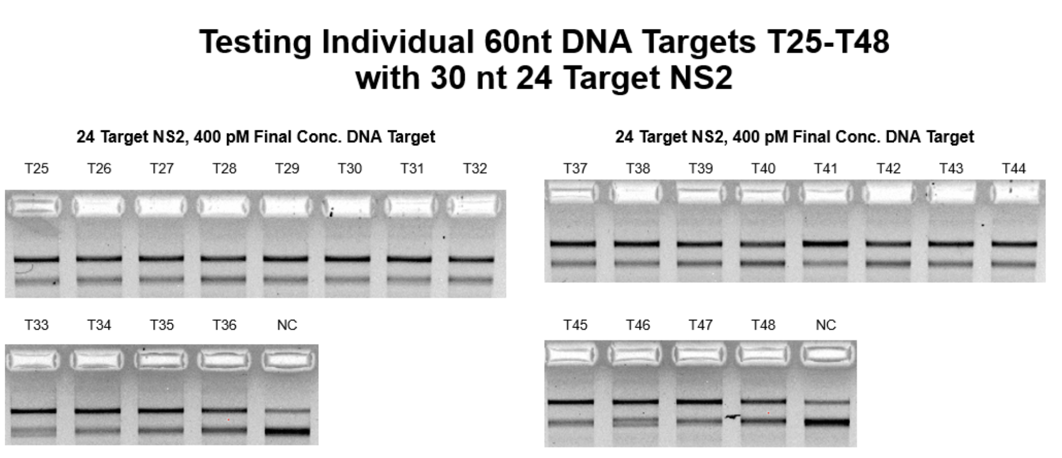

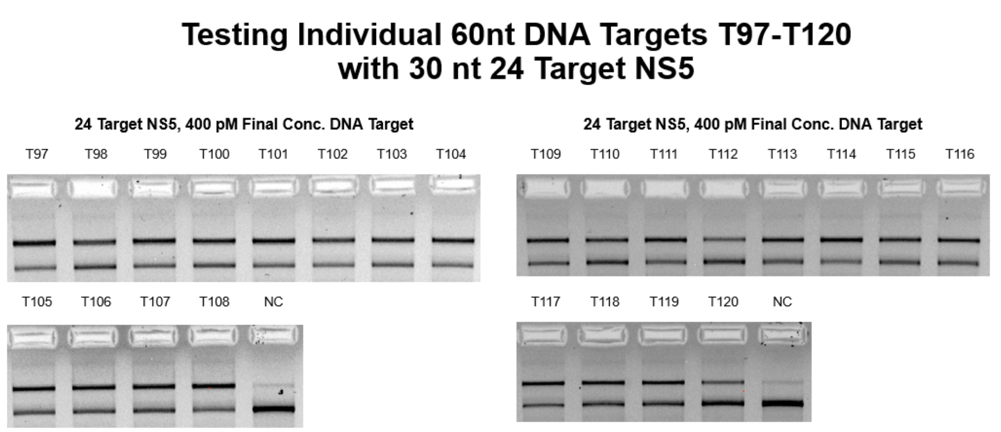

***Figure S11 - Validating individual targets for the 30nt multi-detector nanoswitches***

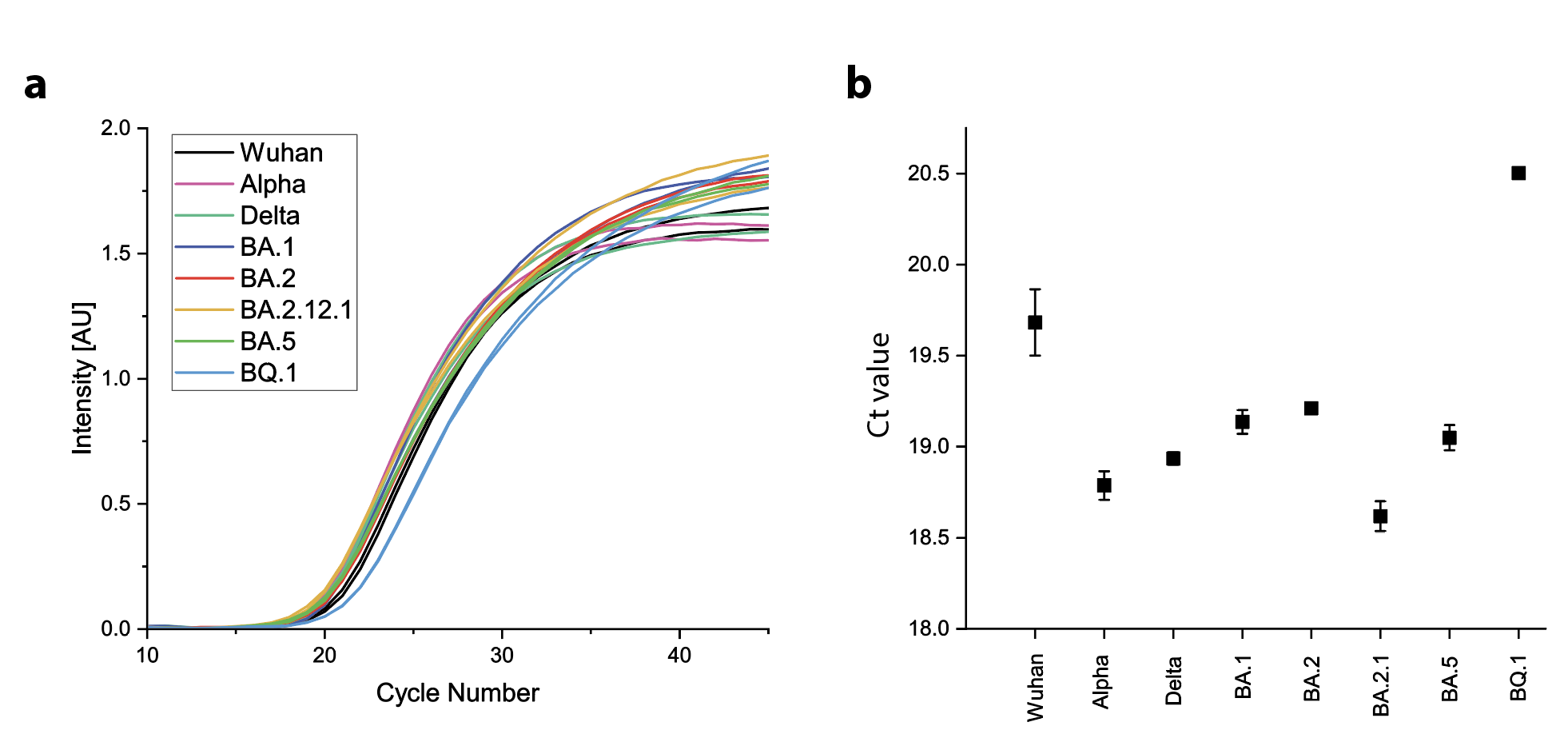

***Figure S12 – qPCR analysis of SARS-CoV-2 IVT controls***

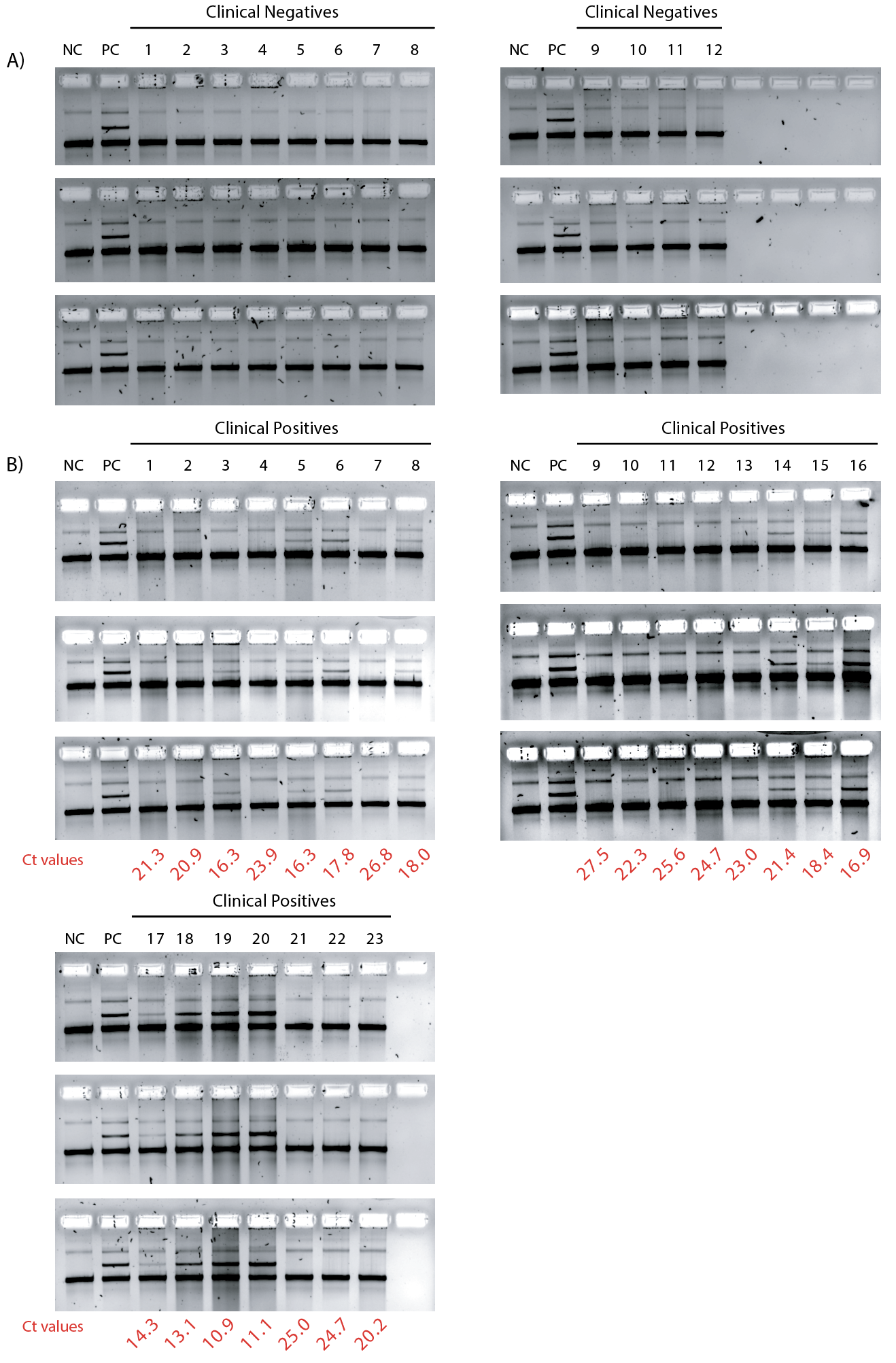

***Figure S13 – Experimental triplicates of clinical samples***

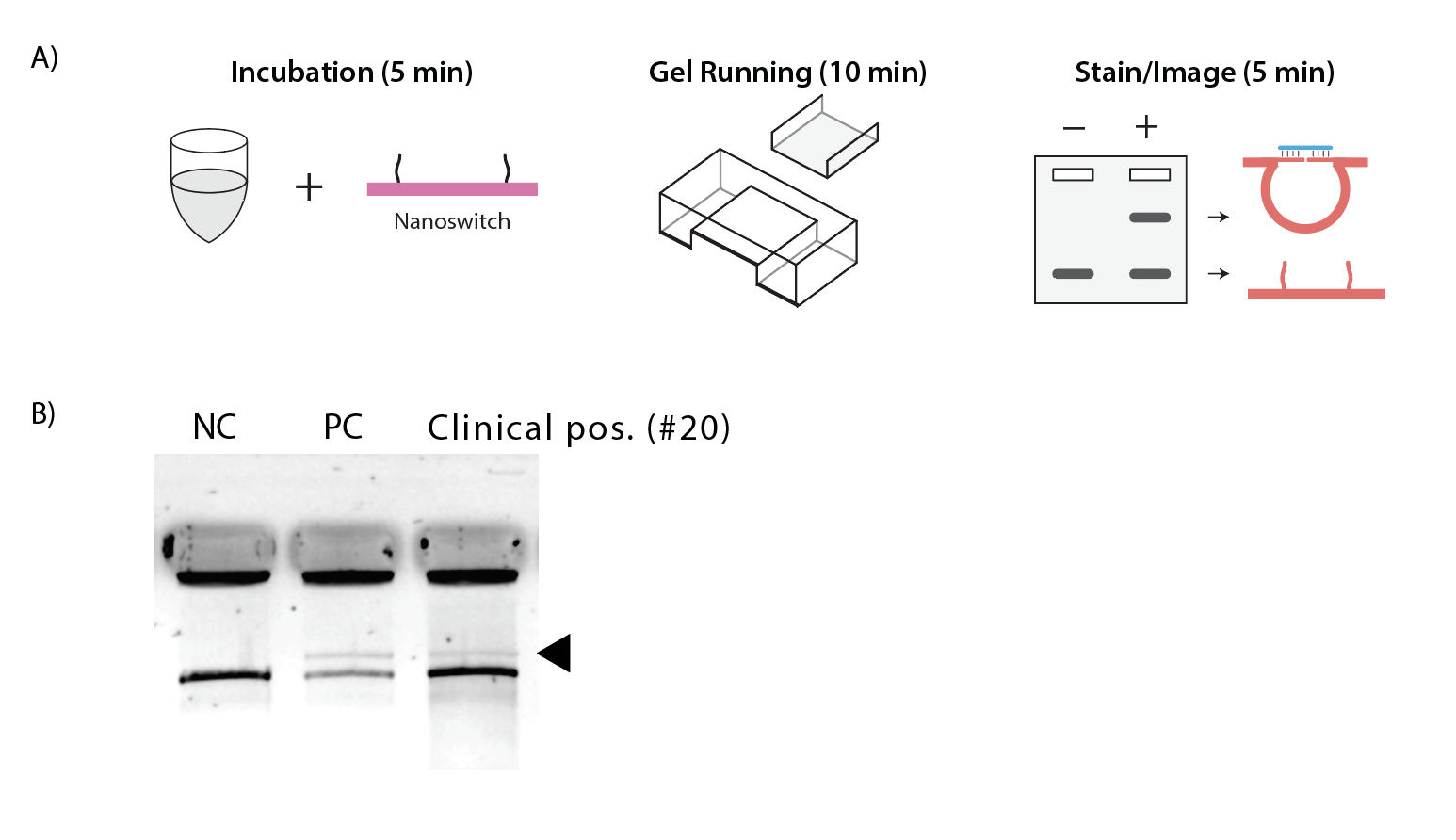

***Figure S14 – Rapid detection of clinical positive***

***Table S1 – Target sequences***

| **Name** | **Sequence** | **Position** |
| --- | --- | --- |
| Single-CDC | GACCCCAAAATCAGCGAAATGCACCCCGCA | 28217 |
| **5 Target** |  |  |
| 5T-1 | GGATGCTCGAACTGCACCTCATGGTCATGT | 487 |
| 5T-2 | GCTATGTCGATAACAACTTCTGTGGCCCTG | 816 |
| 5T-3 | CGACAGATGTCTTGTGCTGCCGGTACTACA | 12713 |
| 5T-4 | ACCGTCTGCGGTATGTGGAAAGGTTATGGC | 13376 |
| 5T-5 | TGATTTCATACAAACCACGCCAGGTAGTGG | 14100 |
| **120 Target (30 nt)** | |  |
| **Nanoswitch 1 30 nt targets** | |  |
| NS1-30nt-T1 | GGATGCTCGAACTGCACCTCATGGTCATGT | 487 |
| NS1-30nt-T2 | GCTATGTCGATAACAACTTCTGTGGCCCTG | 816 |
| NS1-30nt-T3 | CGACAGATGTCTTGTGCTGCCGGTACTACA | 12713 |
| NS1-30nt-T4 | ACCGTCTGCGGTATGTGGAAAGGTTATGGC | 13376 |
| NS1-30nt-T5 | TGATTTCATACAAACCACGCCAGGTAGTGG | 14100 |
| NS1-30nt-T6 | GAGTGCTTTGTTAAGCGTGTTGACTGGACT | 18889 |
| NS1-30nt-T7 | CCTCGTGAAGGTGTCTTTGTTTCAAATGGC | 24830 |
| NS1-30nt-T8 | TTACACTAGCCATCCTTACTGCGCTTCGAT | 26330 |
| NS1-30nt-T9 | GACCCCAAAATCAGCGAAATGCACCCCGCA | 28287 |
| NS1-30nt-T10 | TCGGAATGTCGCGCATTGGCATGGAAGTCA | 29217 |
| NS1-30nt-T11 | GTAACTCGTCTATCTTCTGCAGGCTGCTTA | 167 |
| NS1-30nt-T12 | CCAAATGTGCCTTTCAACTCTCATGAAGTG | 1204 |
| NS1-30nt-T13 | CTAGTAATCCTACCACATTCCACCTAGATG | 4878 |
| NS1-30nt-T14 | TCATCTCGCAAAGGCTCTCAATGACTTCAG | 9961 |
| NS1-30nt-T15 | CTGAGGACAAGAGGGCAAAAGTTACTAGTG | 12318 |
| NS1-30nt-T16 | GTTCCCACCTACAAGTTTTGGACCACTAGT | 14400 |
| NS1-30nt-T17 | CAGGAGATGCCACAACTGCTTATGCTAATA | 15485 |
| NS1-30nt-T18 | CATCACAGGGCTCAGAATATGACTATGTCA | 17840 |
| NS1-30nt-T19 | GCAACCATGATCTGTATTGTCAAGTCCATG | 18803 |
| NS1-30nt-T20 | ATGTCTCTCAGCCTTTTCTTATGGACCTTG | 22071 |
| NS1-30nt-T21 | TCTTATGTCCTTCCCTCAGTCAGCACCTCA | 24706 |
| NS1-30nt-T22 | CGACGAGCTTGGCACTGATCCTTATGAAGA | 703 |
| NS1-30nt-T23 | CTATGCACGCTGCTTCTGGTAATCTATTAC | 14576 |
| NS1-30nt-T24 | TTCTTGGCACTGATAACACTCGCTACTTGT | 27409 |
| **Nanoswitch 2 30 nt targets** | |  |
| NS2-30nt-T1 | CCTGGTTTCAACGAGAAAACACACGTCCAA | 281 |
| NS2-30nt-T2 | CCTATGTGTTCATCAAACGTTCGGATGCTC | 465 |
| NS2-30nt-T3 | ACGGTAATAAAGGAGCTGGTGGCCATAGTT | 642 |
| NS2-30nt-T4 | TGGTAAAGCTTCATGCACTTTGTCCGAACA | 889 |
| NS2-30nt-T5 | CTGCTGCCGTGAACATGAGCATGAAATTGC | 952 |
| NS2-30nt-T6 | TGCCATAACAAGTGTGCCTATTGGGTTCCA | 1511 |
| NS2-30nt-T7 | TTCGCAGTGGCTAACTAACATCTTTGGCAC | 2092 |
| NS2-30nt-T8 | CCTGTGCAAAGGAAATTAAGGAGAGTGTTC | 2262 |
| NS2-30nt-T9 | CACGCACTCAAAGGGATTGTACAGAAAGTG | 2395 |
| NS2-30nt-T10 | GCCCTTGCACCTAATATGATGGTAACAAAC | 2666 |
| NS2-30nt-T11 | TCGGTACAGAAGTAAATGAGTTCGCCTGTG | 2856 |
| NS2-30nt-T12 | CAGTATCTGAATTACTTACACCACTGGGCA | 2919 |
| NS2-30nt-T13 | GAATTTGGTGCCACTTCTGCTGCTCTTCAA | 3146 |
| NS2-30nt-T14 | CAAGACGGCAGTGAGGACAATCAGACAACT | 3236 |
| NS2-30nt-T15 | CGGACACAATCTTGCTAAACACTGTCTTCA | 3583 |
| NS2-30nt-T16 | ACTGTTCGCACAAATGTCTACTTAGCTGTC | 3767 |
| NS2-30nt-T17 | GCAATCTTCATCCAGATTCTGCCACTCTTG | 4047 |
| NS2-30nt-T18 | TTGGAAGAAGCTGCTCGGTATATGAGATCT | 4640 |
| NS2-30nt-T19 | GAAGTTTAATCCACCTGCTCTACAAGATGC | 5329 |
| NS2-30nt-T20 | GTCAGAGGACGCGCAGGGAATGGATAATCT | 6352 |
| NS2-30nt-T21 | CCTTGCTACTCATGGTTTAGCTGCTGTTAA | 6628 |
| NS2-30nt-T22 | TCTTGTACAAATGGCCCCGATTTCAGCTAT | 7375 |
| NS2-30nt-T23 | TTGCCTGGCACGATATTACGCACAACTAAT | 8870 |
| NS2-30nt-T24 | TGAAAGTTTACGCCCTGACACACGTTATGT | 9100 |
| **Nanoswitch 3 30 nt targets** | |  |
| NS3-30nt-T1 | TACTGTAGGCACGGCACTTGTGAAAGATCA | 9212 |
| NS3-30nt-T2 | TCAACCTATTGGTGCTTTGGACATATCAGC | 9370 |
| NS3-30nt-T3 | GTACTTTTGAAGAAGCTGCGCTGTGCACCT | 9783 |
| NS3-30nt-T4 | CGGTCTTTGGCTTGATGACGTAGTTTACTG | 10138 |
| NS3-30nt-T5 | GGTACAGGCTGGTAATGTTCAACTCAGGGT | 10255 |
| NS3-30nt-T6 | TGACATACTAGGACCTCTTTCTGCTCAAAC | 10795 |
| NS3-30nt-T7 | GCAAGAACTGTGTATGATGATGGTGCTAGG | 11354 |
| NS3-30nt-T8 | ACTCAACCGCTACTTTAGACTGACTCTTGG | 11662 |
| NS3-30nt-T9 | CAAGCTATAGCCTCAGAGTTTAGTTCCCTT | 12089 |
| NS3-30nt-T10 | TGGCTAAATCTGAATTTGACCGTGATGCAG | 12222 |
| NS3-30nt-T11 | GGACAATTCACCTAATTTAGCATGGCCTCT | 12613 |
| NS3-30nt-T12 | AGTTTAGCTGCCACAGTACGTCTACAAGCT | 12998 |
| NS3-30nt-T13 | TAACAGTTACACCGGAAGCCAATATGGATC | 13188 |
| NS3-30nt-T14 | CAACTCCGCGAACCCATGCTTCAGTCAGCT | 13418 |
| NS3-30nt-T15 | ATACACAATGGCAGACCTCGTCTATGCTTT | 13803 |
| NS3-30nt-T16 | AATTCTGTGATGCCATGCGAAATGCTGGTA | 14012 |
| NS3-30nt-T17 | CCAAGTCATCGTCAACAACCTAGACAAATC | 14913 |
| NS3-30nt-T18 | ATGCCATTAGTGCAAAGAATAGAGCTCGCA | 15077 |
| NS3-30nt-T19 | CCTTACCCAGATCCATCAAGAATCCTAGGG | 15928 |
| NS3-30nt-T20 | CTATGTACACACCGCATACAGTCTTACAGG | 16208 |
| NS3-30nt-T21 | GCAGCAGAAACGCTCAAAGCTACTGAGGAG | 16636 |
| NS3-30nt-T22 | TCTTTCATGGGAAGTTGGTAAACCTAGACC | 16728 |
| NS3-30nt-T23 | AGCTCTCTACTACCCTTCTGCTCGCATAGT | 17121 |
| NS3-30nt-T24 | CTGTAAATGCATTGCCTGAGACGACAGCAG | 17312 |
| **Nanoswitch 4 30 nt targets** | |  |
| NS4-30nt-T1 | CACTATGTGTACATTGGCGACCCTGCTCAA | 17419 |
| NS4-30nt-T2 | GAAAACTATAGGTCCAGACATGTTCCTCGG | 17523 |
| NS4-30nt-T3 | TCCTTACACGTAACCCTGCTTGGAGAAAAG | 17732 |
| NS4-30nt-T4 | ATCCTACACAGGCACCTACACACCTCAGTG | 18095 |
| NS4-30nt-T5 | GTTGACATACCTGGCATACCTAAGGACATG | 18157 |
| NS4-30nt-T6 | GGTTACCCTAACATGTTTATCACCCGCGAA | 18241 |
| NS4-30nt-T7 | GTGTTAACCTAGTTGCTGTACCTACAGGTT | 18380 |
| NS4-30nt-T8 | GTGCTAAACCACCGCCTGGAGATCAATTTA | 18449 |
| NS4-30nt-T9 | ATAGGACCTGAGCGCACCTGTTGTCTATGT | 18640 |
| NS4-30nt-T10 | ATTACTCTGACAGTCCATGTGAGTCTCATG | 19376 |
| NS4-30nt-T11 | GATGCTTATAACATGATGATCTCAGCTGGC | 19525 |
| NS4-30nt-T12 | CTTTGATGGACAACAGGGTGAAGTACCAGT | 19662 |
| NS4-30nt-T13 | ACTGTGATCTGGGACTACAAAAGAGATGCT | 19867 |
| NS4-30nt-T14 | ATTACAATCTAGTCAAGCGTGGCAACCGGG | 20652 |
| NS4-30nt-T15 | GTACGCTGCTTGTCGATTCAGATCTTAATG | 20933 |
| NS4-30nt-T16 | CAACAAAAGCTAGCTCTTGGAGGTTCCGTG | 21130 |
| NS4-30nt-T17 | CCCCCTGCATACACTAATTCTTTCACACGT | 21635 |
| NS4-30nt-T18 | GGACCAATGGTACTAAGAGGTTTGATAACC | 21777 |
| NS4-30nt-T19 | GATTCGAAGACCCAGTCCCTACTTATTGTT | 21893 |
| NS4-30nt-T20 | CTGTAGACTGTGCACTTGACCCTCTCTCAG | 22425 |
| NS4-30nt-T21 | CTATCAGGCCGGTAGCACACCTTGTAATGG | 22978 |
| NS4-30nt-T22 | CTTTTGAACTTCTACATGCACCAGCAACTG | 23103 |
| NS4-30nt-T23 | CCAGGAACAAATACTTCTAACCAGGTTGCT | 23360 |
| NS4-30nt-T24 | CATGCAGATCAACTTACTCCTACTTGGCGT | 23435 |
| **Nanoswitch 5 30 nt targets** | |  |
| NS5-30nt-T1 | GGTATATGCGCTAGTTATCAGACTCAGACT | 23567 |
| NS5-30nt-T2 | GTCAATCCATCATTGCCTACACTATGTCAC | 23628 |
| NS5-30nt-T3 | TTACCAGATCCATCAAAACCAAGCAAGAGG | 23978 |
| NS5-30nt-T4 | TCAAGATGTGGTCAACCAAAATGCACAAGC | 24406 |
| NS5-30nt-T5 | CAACCTGAATTAGACTCATTCAAGGAGGAG | 24986 |
| NS5-30nt-T6 | ACTGCAACGATACCGATACAAGCCTCACTC | 25486 |
| NS5-30nt-T7 | GTTTACTCACACCTTTTGCTCGTTGCTGCT | 25660 |
| NS5-30nt-T8 | GATGAACCGACGACGACTACTAGCGTGCCT | 26185 |
| NS5-30nt-T9 | GGCACTATTCTGACCAGACCGCTTCTAGAA | 26898 |
| NS5-30nt-T10 | CCTTCGTGGACATCTTCGTATTGCTGGACA | 26954 |
| NS5-30nt-T11 | AGGACCTGCCTAAAGAAATCACTGTTGCTA | 27007 |
| NS5-30nt-T12 | GCACTCAATTTGCTTTTGCTTGTCCTGACG | 27572 |
| NS5-30nt-T13 | ATGTAGTTGATGACCCGTGTCCTATTCACT | 27985 |
| NS5-30nt-T14 | GCGCGATCAAAACAACGTCGGCCCCAAGGT | 28376 |
| NS5-30nt-T15 | TCACCGCTCTCACTCAACATGGCAAGGAAG | 28431 |
| NS5-30nt-T16 | GCAGTCCAGATGACCAAATTGGCTACTACC | 28506 |
| NS5-30nt-T17 | CAGAAGCTGGACTTCCCTATGGTGCTAACA | 28623 |
| NS5-30nt-T18 | CAAAAGATCACATTGGCACCCGCAATCCTG | 28698 |
| NS5-30nt-T19 | CTCTTCTCGTTCCTCATCACGTAGTCGCAA | 28819 |
| NS5-30nt-T20 | CGGCAGACGTGGTCCAGAACAAACCCAAGG | 29095 |
| NS5-30nt-T21 | TGGGGACCAGGAACTAATCAGACAAGGAAC | 29131 |
| NS5-30nt-T22 | CTCAAGCCTTACCGCAGAGACAGAAGAAAC | 29409 |
| NS5-30nt-T23 | GACTCAACTCAGGCCTAAACTCATGCAGAC | 29516 |
| NS5-30nt-T24 | TACAGTGAACAATGCTAGGGAGAGCTGCCT | 29760 |
| **120 Target (60 nt)** | |  |
| **Nanoswitch 1 60 nt targets** | |  |
| NS1-60nt-T1 | CAAACCAACCAACTTTCGATCTCTTGTAGATCTGTTCTCTAAACGAACTTTAAAATCTGT | 28 |
| NS1-60nt-T2 | GAGCCTTGTCCCTGGTTTCAACGAGAAAACACACGTCCAACTCAGTTTGCCTGTTTTACA | 271 |
| NS1-60nt-T3 | TCATTTGACTTAGGCGACGAGCTTGGCACTGATCCTTATGAAGATTTTCAAGAAAACTGG | 689 |
| NS1-60nt-T4 | AATGTCCAAATTTTGTATTTCCCTTAAATTCCATAATCAAGACTATTCAACCAAGGGTTG | 1071 |
| NS1-60nt-T5 | TTACTTACCCCAAAATGCTGTTGTTAAAATTTATTGTCCAGCATGTCACAATTCAGAAGT | 1339 |
| NS1-60nt-T6 | GGTCTTAATGACAACCTTCTTGAAATACTCCAAAAAGAGAAAGTCAACATCAATATTGTT | 1598 |
| NS1-60nt-T7 | AATTTTAAAGTTACAAAAGGAAAAGCTAAAAAAGGTGCCTGGAATATTGGTGAACAGAAA | 1787 |
| NS1-60nt-T8 | TAACTAACATCTTTGGCACTGTTTATGAAAAACTCAAACCCGTCCTTGATTGGCTTGAAG | 2103 |
| NS1-60nt-T9 | GTTAAATCCAGAGAAGAAACTGGCCTACTCATGCCTCTAAAAGCCCCAAAAGAAATTATC | 2426 |
| NS1-60nt-T10 | AGAGTGTGAATATCACTTTTGAACTTGATGAAAGGATTGATAAAGTACTTAATGAGAAGT | 2775 |
| NS1-60nt-T11 | GAGGATGAAGAAGAAGGTGATTGTGAAGAAGAAGAGTTTGAGCCATCAACTCAATATGAG | 3050 |
| NS1-60nt-T12 | ACAAGACGGCAGTGAGGACAATCAGACAACTACTATTCAAACAATTGTTGAGGTTCAACC | 3235 |
| NS1-60nt-T13 | ACTAACAATGCCATGCAAGTTGAATCTGATGATTACATAGCTACTAATGGACCACTTAAA | 3500 |
| NS1-60nt-T14 | GCTTATGAAAATTTTAATCAGCACGAAGTTCTACTTGCACCATTATTATCAGCTGGTATT | 3665 |
| NS1-60nt-T15 | TTTTGGAAATGAAGAGTGAAAAGCAAGTTGAACAAAAGATCGCTGAGATTCCTAAAGAGG | 3834 |
| NS1-60nt-T16 | AAGTTCCTCACAGAAAACTTGTTACTTTATATTGACATTAATGGCAATCTTCATCCAGAT | 4004 |
| NS1-60nt-T17 | GAAATGCTAGCGAAAGCTTTGAGAAAAGTGCCAACAGACAATTATATAACCACTTACCCG | 4196 |
| NS1-60nt-T18 | AAACTAAAGCCATAGTTTCAACTATACAGCGTAAATATAAGGGTATTAAAATACAAGAGG | 4446 |
| NS1-60nt-T19 | TATCTTACTTCTTCTTCTAAAACACCTGAAGAACATTTTATTGAAACCATCTCACTTGCT | 4730 |
| NS1-60nt-T20 | GATGGAGCTGATGTTACTAAAATAAAACCTCATAATTCACATGAAGGTAAAACATTTTAT | 5063 |
| NS1-60nt-T21 | TAACACTCCAACAAATAGAGTTGAAGTTTAATCCACCTGCTCTACAAGATGCTTATTACA | 5307 |
| NS1-60nt-T22 | TGCAAAAGAGTCTTGAACGTGGTGTGTAAAACTTGTGGACAACAGCAGACAACCCTTAAG | 5495 |
| NS1-60nt-T23 | TTACCAGTGTGGTCACTATAAACATATAACTTCTAAAGAAACTTTGTATTGCATAGACGG | 5755 |
| NS1-60nt-T24 | ATTCTTATTTCACAGAGCAACCAATTGATCTTGTACCAAACCAACCATATCCAAACGCAA | 5976 |
| **Nanoswitch 2 60 nt targets** | |  |
| NS2-60nt-T1 | TGATTATAAACACTACACACCCTCTTTTAAGAAAGGAGCTAAATTGTTACATAAACCTAT | 6175 |
| NS2-60nt-T2 | ACTTAAACCAGCAAATAATAGTTTAAAAATTACAGAAGAGGTTGGCCACACAGATCTAAT | 6493 |
| NS2-60nt-T3 | AAACCGTGTTTGTACTAATTATATGCCTTATTTCTTTACTTTATTGCTACAATTGTGTAC | 6748 |
| NS2-60nt-T4 | AAACTGATAAATATTATAATTTGGTTTTTACTATTAAGTGTTTGCCTAGGTTCTTTAATC | 6938 |
| NS2-60nt-T5 | TTCTTTAGACACCTATCCTTCTTTAGAAACTATACAAATTACCATTTCATCTTTTAAATG | 7153 |
| NS2-60nt-T6 | GGCCCCGATTTCAGCTATGGTTAGAATGTACATCTTCTTTGCATCATTTTATTATGTATG | 7387 |
| NS2-60nt-T7 | AGTTTAAAAGACCAATAAATCCTACTGACCAGTCTTCTTACATCGTTGATAGTGTTACAG | 7692 |
| NS2-60nt-T8 | TATTAATGTTATAGTTTTTGATGGTAAATCAAAATGTGAAGAATCATCTGCAAAATCAGC | 7876 |
| NS2-60nt-T9 | AATACGTTTTCATCAACTTTTAACGTACCAATGGAAAAACTCAAAACACTAGTTGCAACT | 8051 |
| NS2-60nt-T10 | TTGTAATAACTATATGCTCACCTATAACAAAGTTGAAAACATGACACCCCGTGACCTTGG | 8272 |
| NS2-60nt-T11 | GTTCCTTTTTGTTGCTGCTATTTTCTATTTAATAACACCTGTTCATGTCATGTCTAAACA | 8602 |
| NS2-60nt-T12 | AACATCTGTTACACACCATCAAAACTTATAGAGTACACTGACTTTGCAACATCAGCTTGT | 8945 |
| NS2-60nt-T13 | TGACACACGTTATGTGCTCATGGATGGCTCTATTATTCAATTTCCTAACACCTACCTTGA | 9115 |
| NS2-60nt-T14 | TCATGTAGTTGCCTTTAATACTTTACTATTCCTTATGTCATTCACTGTACTCTGTTTAAC | 9490 |
| NS2-60nt-T15 | GTTATGTTCACACCTTTAGTACCTTTCTGGATAACAATTGCTTATATCATTTGTATTTCC | 9653 |
| NS2-60nt-T16 | ACTTCAGTAACTCAGGTTCTGATGTTCTTTACCAACCACCACAAACCTCTATCACCTCAG | 9984 |
| NS2-60nt-T17 | TTGATACAGCCAATCCTAAGACACCTAAGTATAAGTTTGTTCGCATTCAACCAGGACAGA | 10326 |
| NS2-60nt-T18 | AATGGTTCATGTGGTAGTGTTGGTTTTAACATAGATTATGACTGTGTCTCTTTTTGTTAC | 10478 |
| NS2-60nt-T19 | CTATGAAGTACAATTATGAACCTCTAACACAAGACCATGTTGACATACTAGGACCTCTTT | 10755 |
| NS2-60nt-T20 | GTACTCAATGGTCTTTGTTCTTTTTTTTGTATGAAAATGCCTTTTTACCTTTTGCTATGG | 11055 |
| NS2-60nt-T21 | GTTTTAAGCTAAAAGACTGTGTTATGTATGCATCAGCTGTAGTGTTACTAATCCTTATGA | 11292 |
| NS2-60nt-T22 | TCATGTTTTTGGCCAGAGGTATTGTTTTTATGTGTGTTGAGTATTGCCCTATTTTCTTCA | 11517 |
| NS2-60nt-T23 | TACAGTCTAAAATGTCAGATGTAAAGTGCACATCAGTAGTCTTACTCTCAGTTTTGCAAC | 11838 |
| NS2-60nt-T24 | GGCAACCTTACAAGCTATAGCCTCAGAGTTTAGTTCCCTTCCATCATATGCAGCTTTTGC | 12079 |
| **Nanoswitch 3 60 nt targets** | |  |
| NS3-60nt-T1 | TATGCAGACAATGCTTTTCACTATGCTTAGAAAGTTGGATAATGATGCACTCAACAACAT | 12349 |
| NS3-60nt-T2 | TGAAATTAGTATGGACAATTCACCTAATTTAGCATGGCCTCTTATTGTAACAGCTTTAAG | 12601 |
| NS3-60nt-T3 | TTTACAGGATTTGAAATGGGCTAGATTCCCTAAGAGTGATGGAACTGGTACTATCTATAC | 12826 |
| NS3-60nt-T4 | GCCTGCCAATTCAACTGTATTATCTTTCTGTGCTTTTGCTGTAGATGCTGCTAAAGCTTA | 13045 |
| NS3-60nt-T5 | ACAAATACCTACAACTTGTGCTAATGACCCTGTGGGTTTTACACTTAAAAACACAGTCTG | 13315 |
| NS3-60nt-T6 | ATAAAGTAGCTGGTTTTGCTAAATTCCTAAAAACTAATTGTTGTCGCTTCCAAGAAAAGG | 13559 |
| NS3-60nt-T7 | CCACATATATCACGTCAACGTCTTACTAAATACACAATGGCAGACCTCGTCTATGCTTTA | 13774 |
| NS3-60nt-T8 | TATTTCAATAAAAAGGACTGGTATGATTTTGTAGAAAACCCAGATATATTACGCGTATAC | 13906 |
| NS3-60nt-T9 | GGTAGTGGAGTTCCTGTTGTAGATTCTTATTATTCATTGTTAATGCCTATATTAACCTTG | 14122 |
| NS3-60nt-T10 | CATTGTGCAAACTTTAATGTTTTATTCTCTACAGTGTTCCCACCTACAAGTTTTGGACCA | 14365 |
| NS3-60nt-T11 | TTACTAACAATGTTGCTTTTCAAACTGTCAAACCCGGTAATTTTAACAAAGACTTCTATG | 14642 |
| NS3-60nt-T12 | CTGTATTAATGCTAACCAAGTCATCGTCAACAACCTAGACAAATCAGCTGGTTTTCCATT | 14898 |
| NS3-60nt-T13 | CATCCCTACTATAACTCAAATGAATCTTAAGTATGCCATTAGTGCAAAGAATAGAGCTCG | 15045 |
| NS3-60nt-T14 | CATGCCTAACATGCTTAGAATTATGGCCTCACTTGTTCTTGCTCGCAAACATACAACGTG | 15315 |
| NS3-60nt-T15 | ATTTGCGTAAACATTTCTCAATGATGATACTCTCTGACGATGCTGTTGTGTGTTTCAATA | 15683 |
| NS3-60nt-T16 | TGATTATGTGTACCTTCCTTACCCAGATCCATCAAGAATCCTAGGGGCCGGCTGTTTTGT | 15912 |
| NS3-60nt-T17 | ATTTGTACTTACAATACATAAGAAAGCTACATGATGAGTTAACAGGACACATGTTAGACA | 16085 |
| NS3-60nt-T18 | AAATGCTGTTACGACCATGTCATATCAACATCACATAAATTAGTCTTGTCTGTTAATCCG | 16318 |
| NS3-60nt-T19 | GTGACTGGACAAATGCTGGTGATTACATTTTAGCTAACACCTGTACTGAAAGACTCAAGC | 16571 |
| NS3-60nt-T20 | AACCTAGACCACCACTTAACCGAAATTATGTCTTTACTGGTTATCGTGTAACTAAAAACA | 16748 |
| NS3-60nt-T21 | TCATTTTGCTATTGGCCTAGCTCTCTACTACCCTTCTGCTCGCATAGTGTATACAGCTTG | 17103 |
| NS3-60nt-T22 | CAATTACCTGCACCACGCACATTGCTAACTAAGGGCACACTAGAACCAGAATATTTCAAT | 17446 |
| NS3-60nt-T23 | AATTCCTTACACGTAACCCTGCTTGGAGAAAAGCTGTCTTTATTTCACCTTATAATTCAC | 17729 |
| NS3-60nt-T24 | GTAGGAATGTGGCAACTTTACAAGCTGAAAATGTAACAGGACTCTTTAAAGATTGTAGTA | 18017 |
| **Nanoswitch 4 60 nt targets** | |  |
| NS4-60nt-T1 | TTTTAAAATGAATTATCAAGTTAATGGTTACCCTAACATGTTTATCACCCGCGAAGAAGC | 18216 |
| NS4-60nt-T2 | GCTAAACCACCGCCTGGAGATCAATTTAAACACCTCATACCACTTATGTACAAAGGACTT | 18451 |
| NS4-60nt-T3 | GTTGACATCTATGAAGTATTTTGTGAAAATAGGACCTGAGCGCACCTGTTGTCTATGTGA | 18612 |
| NS4-60nt-T4 | TGTTAAGCGTGTTGACTGGACTATTGAATATCCTATAATTGGTGATGAACTGAAGATTAA | 18897 |
| NS4-60nt-T5 | ACAAAGCTTATAAAATAGAAGAATTATTCTATTCTTATGCCACACATTCTGACAAATTCA | 19112 |
| NS4-60nt-T6 | AAGTGCTTTTGTTAATTTAAAACAATTACCATTTTTCTATTACTCTGACAGTCCATGTGA | 19338 |
| NS4-60nt-T7 | TTAGCTTGTGGGTTTACAAACAATTTGATACTTATAACCTCTGGAACACTTTTACAAGAC | 19556 |
| NS4-60nt-T8 | TGACTGACATAGCCAAGAAACCAACTGAAACGATTTGTGCACCACTCACTGTCTTTTTTG | 19931 |
| NS4-60nt-T9 | TTGTCCAACAATTACCTGAAACTTACTTTACTCAGAGTAGAAATTTACAAGAATTTAAAC | 20174 |
| NS4-60nt-T10 | AATTAGAAGATTTTATTCCTATGGACAGTACAGTTAAAAACTATTTCATAACAGATGCGC | 20411 |
| NS4-60nt-T11 | TGGTGTAAAGATGGCCATGTAGAAACATTTTACCCAAAATTACAATCTAGTCAAGCGTGG | 20614 |
| NS4-60nt-T12 | TCAATATTTAAACACATTAACATTAGCTGTACCCTATAATATGAGAGTTATACATTTTGG | 20811 |
| NS4-60nt-T13 | TTATTAGTGATATGTACGACCCTAAGACTAAAAATGTTACAAAAGAAAATGACTCTAAAG | 21038 |
| NS4-60nt-T14 | ACGCGAACAAATAGATGGTTATGTCATGCATGCAAATTACATATTTTGGAGGAATACAAA | 21303 |
| NS4-60nt-T15 | TGTTAATCTTACAACCAGAACTCAATTACCCCCTGCATACACTAATTCTTTCACACGTGG | 21607 |
| NS4-60nt-T16 | GTACTACTTTAGATTCGAAGACCCAGTCCCTACTTATTGTTAATAACGCTACTAATGTTG | 21882 |
| NS4-60nt-T17 | TTTAAGAATATTGATGGTTATTTTAAAATATATTCTAAGCACACGCCTATTAATTTAGTG | 22142 |
| NS4-60nt-T18 | TCTTCAACCTAGGACTTTTCTATTAAAATATAATGAAAATGGAACCATTACAGATGCTGT | 22369 |
| NS4-60nt-T19 | TAAGTGTTATGGAGTGTCTCCTACTAAATTAAATGATCTCTGCTTTACTAATGTCTATGC | 22693 |
| NS4-60nt-T20 | AGGTTTTAATTGTTACTTTCCTTTACAATCATATGGTTTCCAACCCACTAATGGTGTTGG | 23014 |
| NS4-60nt-T21 | CTTTCCAACAATTTGGCAGAGACATTGCTGACACTACTGATGCTGTCCGTGATCCACAGA | 23244 |
| NS4-60nt-T22 | TGCAGATCAACTTACTCCTACTTGGCGTGTTTATTCTACAGGTTCTAATGTTTTTCAAAC | 23437 |
| NS4-60nt-T23 | TAATAACTCTATTGCCATACCCACAAATTTTACTATTAGTGTTACCACAGAAATTCTACC | 23686 |
| NS4-60nt-T24 | AACACCCAAGAAGTTTTTGCACAAGTCAAACAAATTTACAAAACACCACCAATTAAAGAT | 23891 |
| **Nanoswitch 5 60 nt targets** | |  |
| NS5-60nt-T1 | TAACGGCCTTACTGTTTTGCCACCTTTGCTCACAGATGAAATGATTGCTCAATACACTTC | 24127 |
| NS5-60nt-T2 | CTCCAATTTTGGTGCAATTTCAAGTGTTTTAAATGATATCCTTTCACGTCTTGACAAAGT | 24463 |
| NS5-60nt-T3 | TATGTCCCTGCACAAGAAAAGAACTTCACAACTGCTCCTGCCATTTGTCATGATGGAAAA | 24761 |
| NS5-60nt-T4 | TAATAGGAATTGTCAACAACACAGTTTATGATCCTTTGCAACCTGAATTAGACTCATTCA | 24948 |
| NS5-60nt-T5 | GACCGCCTCAATGAGGTTGCCAAGAATTTAAATGAATCTCTCATCGATCTCCAAGAACTT | 25112 |
| NS5-60nt-T6 | TTTGTTTATGAGAATCTTCACAATTGGAACTGTAACTTTGAAGCAAGGTGAAATCAAGGA | 25398 |
| NS5-60nt-T7 | TTTACTCACACCTTTTGCTCGTTGCTGCTGGCCTTGAAGCCCCTTTTCTCTATCTTTATG | 25661 |
| NS5-60nt-T8 | CCCATTACTTTATGATGCCAACTATTTTCTTTGCTGGCATACTAATTGTTACGACTATTG | 25803 |
| NS5-60nt-T9 | TCAATTGAGTACAGACACTGGTGTTGAACATGTTACCTTCTTCATCTACAATAAAATTGT | 26043 |
| NS5-60nt-T10 | TTAATAGTTAATAGCGTACTTCTTTTTCTTGCTTTCGTGGTATTCTTGCTAGTTACACTA | 26278 |
| NS5-60nt-T11 | TAAATATTATATTAGTTTTTCTGTTTGGAACTTTAATTTTAGCCATGGCAGATTCCAACG | 26479 |
| NS5-60nt-T12 | CGCGTTCCATGTGGTCATTCAATCCAGAAACTAACATTCTTCTCAACGTGCCACTCCATG | 26839 |
| NS5-60nt-T13 | GGACCTGCCTAAAGAAATCACTGTTGCTACATCACGAACGCTTTCTTATTACAAATTGGG | 27008 |
| NS5-60nt-T14 | TGGAATCTTGATTACATCATAAACCTCATAATTAAAAATTTATCTAAGTCACTAACTGAG | 27280 |
| NS5-60nt-T15 | TTCACCATTTCATCCTCTAGCTGATAACAAATTTGCACTGACTTGCTTTAGCACTCAATT | 27522 |
| NS5-60nt-T16 | CTTCTATTTGTGCTTTTTAGCCTTTCTGCTATTCCTTGTTTTAATTATGCTTATTATCTT | 27779 |
| NS5-60nt-T17 | CTAAATCACCCATTCAGTACATCGATATCGGTAATTATACAGTTTCCTGTTTACCTTTTA | 28093 |
| NS5-60nt-T18 | CCCCAAGGTTTACCCAATAATACTGCGTCTTGGTTCACCGCTCTCACTCAACATGGCAAG | 28397 |
| NS5-60nt-T19 | CAGACGAATTCGTGGTGGTGACGGTAAAATGAAAGATCTCAGTCCAAGATGGTATTTCTA | 28546 |
| NS5-60nt-T20 | TCAAGCCTCTTCTCGTTCCTCATCACGTAGTCGCAACAGTTCAAGAAATTCAACTCCAGG | 28813 |
| NS5-60nt-T21 | CATACAATGTAACACAAGCTTTCGGCAGACGTGGTCCAGAACAAACCCAAGGAAATTTTG | 29073 |
| NS5-60nt-T22 | CATATTGACGCATACAAAACATTCCCACCAACAGAGCCTAAAAAGGACAAAAAGAAGAAG | 29339 |
| NS5-60nt-T23 | TAAACGTTTTCGCTTTTCCGTTTACGATATATAGTCTACTCTTGTGCAGAATGAATTCTC | 29568 |
| NS5-60nt-T24 | GAAGAGCCCTAATGTGTAAAATTAATTTTAGTAGTGCTATCCCCATGTGATTTTAATAGC | 29795 |

***Table S2 – detector oligo sequences***

| **Target** | **Detector 1 sequence** **(nt 1-30 hybridize M13, 31-45 with target)** | **Detector 2 sequence (nt 1-15 hybridize with target, 16-45 with M13)** |
| --- | --- | --- |
| Single-CDC | CAATACTTCTTTGATTAGTAATAACATCACACATGACCATGAGGT | GCAGTTCGAGCATCCTCAACCGATTGAGGGAGGGAAGGTAAATAT |
| **5 Target** |  |  |
| 5T-1 | TTGCCTGAGTAGAAGAACTCAAACTATCGGACATGACCATGAGGT | GCAGTTCGAGCATCCTCACCGTCACCGACTTGAGCCATTTGGGAA |
| 5T-2 | CCTTGCTGGTAATATCCAGAACAATATTACCAGGGCCACAGAAGT | TGTTATCGACATAGCTTAGAGCCAGCAAAATCACCAGTAGCACCA |
| 5T-3 | CGCCAGCCATTGCAACAGGAAAAACGCTCATGTAGTACCGGCAGC | ACAAGACATCTGTCGTTACCATTAGCAAGGCCGGAAACGTCACCA |
| 5T-4 | TGGAAATACCTACATTTTGACGCTCAATCGGCCATAACCTTTCCA | CATACCGCAGACGGTATGAAACCATCGATAGCAGCACCGTAATCA |
| 5T-5 | TCTGAAATGGATTATTTACATTGGCAGATTCCACTACCTGGCGTG | GTTTGTATGAAATCAGTAGCGACAGAATCAAGTTTGCCTTTAGCG |
| **120 Target (30 nt)** | |  |
| **Nanoswitch 1 30 nt targets** | |  |
| NS1-30nt-T1 | TTGCCTGAGTAGAAGAACTCAAACTATCGGACATGACCATGAGGT | GCAGTTCGAGCATCCTCACCGTCACCGACTTGAGCCATTTGGGAA |
| NS1-30nt-T2 | CCTTGCTGGTAATATCCAGAACAATATTACCAGGGCCACAGAAGT | TGTTATCGACATAGCTTAGAGCCAGCAAAATCACCAGTAGCACCA |
| NS1-30nt-T3 | CGCCAGCCATTGCAACAGGAAAAACGCTCATGTAGTACCGGCAGC | ACAAGACATCTGTCGTTACCATTAGCAAGGCCGGAAACGTCACCA |
| NS1-30nt-T4 | TGGAAATACCTACATTTTGACGCTCAATCGGCCATAACCTTTCCA | CATACCGCAGACGGTATGAAACCATCGATAGCAGCACCGTAATCA |
| NS1-30nt-T5 | TCTGAAATGGATTATTTACATTGGCAGATTCCACTACCTGGCGTG | GTTTGTATGAAATCAGTAGCGACAGAATCAAGTTTGCCTTTAGCG |
| NS1-30nt-T6 | CACCAGTCACACGACCAGTAATAAAAGGGAAGTCCAGTCAACACG | CTTAACAAAGCACTCTCAGACTGTAGCGCGTTTTCATCGGCATTT |
| NS1-30nt-T7 | CATTCTGGCCAACAGAGATAGAACCCTTCTGCCATTTGAAACAAA | GACACCTTCACGAGGTCGGTCATAGCCCCCTTATTAGCGTTTGCC |
| NS1-30nt-T8 | GACCTGAAAGCGTAAGAATACGTGGCACAGATCGAAGCGCAGTAA | GGATGGCTAGTGTAAATCTTTTCATAATCAAAATCACCGGAACCA |
| NS1-30nt-T9 | ACAATATTTTTGAATGGCTATTAGTCTTTATGCGGGGTGCATTTC | GCTGATTTTGGGGTCGAGCCACCACCGGAACCGCCTCCCTCAGAG |
| NS1-30nt-T10 | ATGCGCGAACTGATAGCCCTAAAACATCGCTGACTTCCATGCCAA | TGCGCGACATTCCGACCGCCACCCTCAGAACCGCCACCCTCAGAG |
| NS1-30nt-T11 | CATTAAAAATACCGAACGAACCACCAGCAGTAAGCAGCCTGCAGA | AGATAGACGAGTTACCCACCACCCTCAGAGCCGCCACCAGAACCA |
| NS1-30nt-T12 | AAGATAAAACAGAGGTGAGGCGGTCAGTATCACTTCATGAGAGTT | GAAAGGCACATTTGGCCACCAGAGCCGCCGCCAGCATTGACAGGA |
| NS1-30nt-T13 | TAACACCGCCTGCAACAGTGCCACGCTGAGCATCTAGGTGGAATG | TGGTAGGATTACTAGGGTTGAGGCAGGTCAGACGATTGGCCTTGA |
| NS1-30nt-T14 | AGCCAGCAGCAAATGAAAAATCTAAAGCATCTGAAGTCATTGAGA | GCCTTTGCGAGATGATATTCACAAACAAATAAATCCTCATTAAAG |
| NS1-30nt-T15 | CACCTTGCTGAACCTCAAATATCAAACCCTCACTAGTAACTTTTG | CCCTCTTGTCCTCAGCCAGAATGGAAAGCGCAGTCTCTGAATTTA |
| NS1-30nt-T16 | CAATCAATATCTGGTCAGTTGGCAAATCAAACTAGTGGTCCAAAA | CTTGTAGGTGGGAACCCGTTCCAGTAAGCGTCATACATGGCTTTT |
| NS1-30nt-T17 | CAGTTGAAAGGAATTGAGGAAGGTTATCTATATTAGCATAAGCAG | TTGTGGCATCTCCTGGATGATACAGGAGTGTACTGGTAATAAGTT |
| NS1-30nt-T18 | AAATATCTTTAGGAGCACTAACAACTAATATGACATAGTCATATT | CTGAGCCCTGTGATGTTAACGGGGTCAGTGCCTTGAGTAACAGTG |
| NS1-30nt-T19 | GATTAGAGCCGTCAATAGATAATACATTTGCATGGACTTGACAAT | ACAGATCATGGTTGCCCCGTATAAACAGTTAATGCCCCCTGCCTA |
| NS1-30nt-T20 | AGGATTTAGAAGTATTAGACTTTACAAACACAAGGTCCATAAGAA | AAGGCTGAGAGACATTTTCGGAACCTATTATTCTGAAACATGAAA |
| NS1-30nt-T21 | TCTGTCCATCACGCAAATTAACCGTTGTAGTGAGGTGCTGACTGA | GGGAAGGACATAAGATCAACCGATTGAGGGAGGGAAGGTAAATAT |
| NS1-30nt-T22 | CAATACTTCTTTGATTAGTAATAACATCACTCTTCATAAGGATCA | GTGCCAAGCTCGTCGTGACGGAAATTATTCATTAAAGGTGAATTA |
| NS1-30nt-T23 | ATTCGACAACTCGTATTAAATCCTTTGCCCGTAATAGATTACCAG | AAGCAGCGTGCATAGGTATTAAGAGGCTGAGACTCCTCAAGAGAA |
| NS1-30nt-T24 | GAACGTTATTAATTTTAAAAGTTTGAGTAAACAAGTAGCGAGTGT | TATCAGTGCCAAGAAGGATTAGGATTAGCGGGGTTTTGCTCAGTA |
| **Nanoswitch 2 30 nt targets** | |  |
| NS2-30nt-T1 | TCTGTCCATCACGCAAATTAACCGTTGTAGTTGGACGTGTGTTTT | CTCGTTGAAACCAGGTCAACCGATTGAGGGAGGGAAGGTAAATAT |
| NS2-30nt-T2 | CAATACTTCTTTGATTAGTAATAACATCACGAGCATCCGAACGTT | TGATGAACACATAGGTGACGGAAATTATTCATTAAAGGTGAATTA |
| NS2-30nt-T3 | TTGCCTGAGTAGAAGAACTCAAACTATCGGAACTATGGCCACCAG | CTCCTTTATTACCGTTCACCGTCACCGACTTGAGCCATTTGGGAA |
| NS2-30nt-T4 | CCTTGCTGGTAATATCCAGAACAATATTACTGTTCGGACAAAGTG | CATGAAGCTTTACCATTAGAGCCAGCAAAATCACCAGTAGCACCA |
| NS2-30nt-T5 | CGCCAGCCATTGCAACAGGAAAAACGCTCAGCAATTTCATGCTCA | TGTTCACGGCAGCAGTTACCATTAGCAAGGCCGGAAACGTCACCA |
| NS2-30nt-T6 | TGGAAATACCTACATTTTGACGCTCAATCGTGGAACCCAATAGGC | ACACTTGTTATGGCAATGAAACCATCGATAGCAGCACCGTAATCA |
| NS2-30nt-T7 | TCTGAAATGGATTATTTACATTGGCAGATTGTGCCAAAGATGTTA | GTTAGCCACTGCGAAGTAGCGACAGAATCAAGTTTGCCTTTAGCG |
| NS2-30nt-T8 | CACCAGTCACACGACCAGTAATAAAAGGGAGAACACTCTCCTTAA | TTTCCTTTGCACAGGTCAGACTGTAGCGCGTTTTCATCGGCATTT |
| NS2-30nt-T9 | CATTCTGGCCAACAGAGATAGAACCCTTCTCACTTTCTGTACAAT | CCCTTTGAGTGCGTGTCGGTCATAGCCCCCTTATTAGCGTTTGCC |
| NS2-30nt-T10 | GACCTGAAAGCGTAAGAATACGTGGCACAGGTTTGTTACCATCAT | ATTAGGTGCAAGGGCATCTTTTCATAATCAAAATCACCGGAACCA |
| NS2-30nt-T11 | ACAATATTTTTGAATGGCTATTAGTCTTTACACAGGCGAACTCAT | TTACTTCTGTACCGAGAGCCACCACCGGAACCGCCTCCCTCAGAG |
| NS2-30nt-T12 | ATGCGCGAACTGATAGCCCTAAAACATCGCTGCCCAGTGGTGTAA | GTAATTCAGATACTGCCGCCACCCTCAGAACCGCCACCCTCAGAG |
| NS2-30nt-T13 | CATTAAAAATACCGAACGAACCACCAGCAGTTGAAGAGCAGCAGA | AGTGGCACCAAATTCCCACCACCCTCAGAGCCGCCACCAGAACCA |
| NS2-30nt-T14 | AAGATAAAACAGAGGTGAGGCGGTCAGTATAGTTGTCTGATTGTC | CTCACTGCCGTCTTGCCACCAGAGCCGCCGCCAGCATTGACAGGA |
| NS2-30nt-T15 | TAACACCGCCTGCAACAGTGCCACGCTGAGTGAAGACAGTGTTTA | GCAAGATTGTGTCCGGGTTGAGGCAGGTCAGACGATTGGCCTTGA |
| NS2-30nt-T16 | AGCCAGCAGCAAATGAAAAATCTAAAGCATGACAGCTAAGTAGAC | ATTTGTGCGAACAGTTATTCACAAACAAATAAATCCTCATTAAAG |
| NS2-30nt-T17 | CACCTTGCTGAACCTCAAATATCAAACCCTCAAGAGTGGCAGAAT | CTGGATGAAGATTGCCCAGAATGGAAAGCGCAGTCTCTGAATTTA |
| NS2-30nt-T18 | CAATCAATATCTGGTCAGTTGGCAAATCAAAGATCTCATATACCG | AGCAGCTTCTTCCAACCGTTCCAGTAAGCGTCATACATGGCTTTT |
| NS2-30nt-T19 | CAGTTGAAAGGAATTGAGGAAGGTTATCTAGCATCTTGTAGAGCA | GGTGGATTAAACTTCGATGATACAGGAGTGTACTGGTAATAAGTT |
| NS2-30nt-T20 | AAATATCTTTAGGAGCACTAACAACTAATAAGATTATCCATTCCC | TGCGCGTCCTCTGACTTAACGGGGTCAGTGCCTTGAGTAACAGTG |
| NS2-30nt-T21 | GATTAGAGCCGTCAATAGATAATACATTTGTTAACAGCAGCTAAA | CCATGAGTAGCAAGGCCCGTATAAACAGTTAATGCCCCCTGCCTA |
| NS2-30nt-T22 | AGGATTTAGAAGTATTAGACTTTACAAACAATAGCTGAAATCGGG | GCCATTTGTACAAGATTTCGGAACCTATTATTCTGAAACATGAAA |
| NS2-30nt-T23 | ATTCGACAACTCGTATTAAATCCTTTGCCCATTAGTTGTGCGTAA | TATCGTGCCAGGCAAGTATTAAGAGGCTGAGACTCCTCAAGAGAA |
| NS2-30nt-T24 | GAACGTTATTAATTTTAAAAGTTTGAGTAAACATAACGTGTGTCA | GGGCGTAAACTTTCAGGATTAGGATTAGCGGGGTTTTGCTCAGTA |
| **Nanoswitch 3 30 nt targets** | |  |
| NS3-30nt-T1 | TCTGTCCATCACGCAAATTAACCGTTGTAGTGATCTTTCACAAGT | GCCGTGCCTACAGTATCAACCGATTGAGGGAGGGAAGGTAAATAT |
| NS3-30nt-T2 | CAATACTTCTTTGATTAGTAATAACATCACGCTGATATGTCCAAA | GCACCAATAGGTTGATGACGGAAATTATTCATTAAAGGTGAATTA |
| NS3-30nt-T3 | TTGCCTGAGTAGAAGAACTCAAACTATCGGAGGTGCACAGCGCAG | CTTCTTCAAAAGTACTCACCGTCACCGACTTGAGCCATTTGGGAA |
| NS3-30nt-T4 | CCTTGCTGGTAATATCCAGAACAATATTACCAGTAAACTACGTCA | TCAAGCCAAAGACCGTTAGAGCCAGCAAAATCACCAGTAGCACCA |
| NS3-30nt-T5 | CGCCAGCCATTGCAACAGGAAAAACGCTCAACCCTGAGTTGAACA | TTACCAGCCTGTACCTTACCATTAGCAAGGCCGGAAACGTCACCA |
| NS3-30nt-T6 | TGGAAATACCTACATTTTGACGCTCAATCGGTTTGAGCAGAAAGA | GGTCCTAGTATGTCAATGAAACCATCGATAGCAGCACCGTAATCA |
| NS3-30nt-T7 | TCTGAAATGGATTATTTACATTGGCAGATTCCTAGCACCATCATC | ATACACAGTTCTTGCGTAGCGACAGAATCAAGTTTGCCTTTAGCG |
| NS3-30nt-T8 | CACCAGTCACACGACCAGTAATAAAAGGGACCAAGAGTCAGTCTA | AAGTAGCGGTTGAGTTCAGACTGTAGCGCGTTTTCATCGGCATTT |
| NS3-30nt-T9 | CATTCTGGCCAACAGAGATAGAACCCTTCTAAGGGAACTAAACTC | TGAGGCTATAGCTTGTCGGTCATAGCCCCCTTATTAGCGTTTGCC |
| NS3-30nt-T10 | GACCTGAAAGCGTAAGAATACGTGGCACAGCTGCATCACGGTCAA | ATTCAGATTTAGCCAATCTTTTCATAATCAAAATCACCGGAACCA |
| NS3-30nt-T11 | ACAATATTTTTGAATGGCTATTAGTCTTTAAGAGGCCATGCTAAA | TTAGGTGAATTGTCCGAGCCACCACCGGAACCGCCTCCCTCAGAG |
| NS3-30nt-T12 | ATGCGCGAACTGATAGCCCTAAAACATCGCAGCTTGTAGACGTAC | TGTGGCAGCTAAACTCCGCCACCCTCAGAACCGCCACCCTCAGAG |
| NS3-30nt-T13 | CATTAAAAATACCGAACGAACCACCAGCAGGATCCATATTGGCTT | CCGGTGTAACTGTTACCACCACCCTCAGAGCCGCCACCAGAACCA |
| NS3-30nt-T14 | AAGATAAAACAGAGGTGAGGCGGTCAGTATAGCTGACTGAAGCAT | GGGTTCGCGGAGTTGCCACCAGAGCCGCCGCCAGCATTGACAGGA |
| NS3-30nt-T15 | TAACACCGCCTGCAACAGTGCCACGCTGAGAAAGCATAGACGAGG | TCTGCCATTGTGTATGGTTGAGGCAGGTCAGACGATTGGCCTTGA |
| NS3-30nt-T16 | AGCCAGCAGCAAATGAAAAATCTAAAGCATTACCAGCATTTCGCA | TGGCATCACAGAATTTATTCACAAACAAATAAATCCTCATTAAAG |
| NS3-30nt-T17 | CACCTTGCTGAACCTCAAATATCAAACCCTGATTTGTCTAGGTTG | TTGACGATGACTTGGCCAGAATGGAAAGCGCAGTCTCTGAATTTA |
| NS3-30nt-T18 | CAATCAATATCTGGTCAGTTGGCAAATCAATGCGAGCTCTATTCT | TTGCACTAATGGCATCCGTTCCAGTAAGCGTCATACATGGCTTTT |
| NS3-30nt-T19 | CAGTTGAAAGGAATTGAGGAAGGTTATCTACCCTAGGATTCTTGA | TGGATCTGGGTAAGGGATGATACAGGAGTGTACTGGTAATAAGTT |
| NS3-30nt-T20 | AAATATCTTTAGGAGCACTAACAACTAATACCTGTAAGACTGTAT | GCGGTGTGTACATAGTTAACGGGGTCAGTGCCTTGAGTAACAGTG |
| NS3-30nt-T21 | GATTAGAGCCGTCAATAGATAATACATTTGCTCCTCAGTAGCTTT | GAGCGTTTCTGCTGCCCCGTATAAACAGTTAATGCCCCCTGCCTA |
| NS3-30nt-T22 | AGGATTTAGAAGTATTAGACTTTACAAACAGGTCTAGGTTTACCA | ACTTCCCATGAAAGATTTCGGAACCTATTATTCTGAAACATGAAA |
| NS3-30nt-T23 | ATTCGACAACTCGTATTAAATCCTTTGCCCACTATGCGAGCAGAA | GGGTAGTAGAGAGCTGTATTAAGAGGCTGAGACTCCTCAAGAGAA |
| NS3-30nt-T24 | GAACGTTATTAATTTTAAAAGTTTGAGTAACTGCTGTCGTCTCAG | GCAATGCATTTACAGGGATTAGGATTAGCGGGGTTTTGCTCAGTA |
| **Nanoswitch 4 30 nt targets** | |  |
| NS4-30nt-T1 | TCTGTCCATCACGCAAATTAACCGTTGTAGTTGAGCAGGGTCGCC | AATGTACACATAGTGTCAACCGATTGAGGGAGGGAAGGTAAATAT |
| NS4-30nt-T2 | CAATACTTCTTTGATTAGTAATAACATCACCCGAGGAACATGTCT | GGACCTATAGTTTTCTGACGGAAATTATTCATTAAAGGTGAATTA |
| NS4-30nt-T3 | TTGCCTGAGTAGAAGAACTCAAACTATCGGCTTTTCTCCAAGCAG | GGTTACGTGTAAGGATCACCGTCACCGACTTGAGCCATTTGGGAA |
| NS4-30nt-T4 | CCTTGCTGGTAATATCCAGAACAATATTACCACTGAGGTGTGTAG | GTGCCTGTGTAGGATTTAGAGCCAGCAAAATCACCAGTAGCACCA |
| NS4-30nt-T5 | CGCCAGCCATTGCAACAGGAAAAACGCTCACATGTCCTTAGGTAT | GCCAGGTATGTCAACTTACCATTAGCAAGGCCGGAAACGTCACCA |
| NS4-30nt-T6 | TGGAAATACCTACATTTTGACGCTCAATCGTTCGCGGGTGATAAA | CATGTTAGGGTAACCATGAAACCATCGATAGCAGCACCGTAATCA |
| NS4-30nt-T7 | TCTGAAATGGATTATTTACATTGGCAGATTAACCTGTAGGTACAG | CAACTAGGTTAACACGTAGCGACAGAATCAAGTTTGCCTTTAGCG |
| NS4-30nt-T8 | CACCAGTCACACGACCAGTAATAAAAGGGATAAATTGATCTCCAG | GCGGTGGTTTAGCACTCAGACTGTAGCGCGTTTTCATCGGCATTT |
| NS4-30nt-T9 | CATTCTGGCCAACAGAGATAGAACCCTTCTACATAGACAACAGGT | GCGCTCAGGTCCTATTCGGTCATAGCCCCCTTATTAGCGTTTGCC |
| NS4-30nt-T10 | GACCTGAAAGCGTAAGAATACGTGGCACAGCATGAGACTCACATG | GACTGTCAGAGTAATATCTTTTCATAATCAAAATCACCGGAACCA |
| NS4-30nt-T11 | ACAATATTTTTGAATGGCTATTAGTCTTTAGCCAGCTGAGATCAT | CATGTTATAAGCATCGAGCCACCACCGGAACCGCCTCCCTCAGAG |
| NS4-30nt-T12 | ATGCGCGAACTGATAGCCCTAAAACATCGCACTGGTACTTCACCC | TGTTGTCCATCAAAGCCGCCACCCTCAGAACCGCCACCCTCAGAG |
| NS4-30nt-T13 | CATTAAAAATACCGAACGAACCACCAGCAGAGCATCTCTTTTGTA | GTCCCAGATCACAGTCCACCACCCTCAGAGCCGCCACCAGAACCA |
| NS4-30nt-T14 | AAGATAAAACAGAGGTGAGGCGGTCAGTATCCCGGTTGCCACGCT | TGACTAGATTGTAATCCACCAGAGCCGCCGCCAGCATTGACAGGA |
| NS4-30nt-T15 | TAACACCGCCTGCAACAGTGCCACGCTGAGCATTAAGATCTGAAT | CGACAAGCAGCGTACGGTTGAGGCAGGTCAGACGATTGGCCTTGA |
| NS4-30nt-T16 | AGCCAGCAGCAAATGAAAAATCTAAAGCATCACGGAACCTCCAAG | AGCTAGCTTTTGTTGTATTCACAAACAAATAAATCCTCATTAAAG |
| NS4-30nt-T17 | CACCTTGCTGAACCTCAAATATCAAACCCTACGTGTGAAAGAATT | AGTGTATGCAGGGGGCCAGAATGGAAAGCGCAGTCTCTGAATTTA |
| NS4-30nt-T18 | CAATCAATATCTGGTCAGTTGGCAAATCAAGGTTATCAAACCTCT | TAGTACCATTGGTCCCCGTTCCAGTAAGCGTCATACATGGCTTTT |
| NS4-30nt-T19 | CAGTTGAAAGGAATTGAGGAAGGTTATCTAAACAATAAGTAGGGA | CTGGGTCTTCGAATCGATGATACAGGAGTGTACTGGTAATAAGTT |
| NS4-30nt-T20 | AAATATCTTTAGGAGCACTAACAACTAATACTGAGAGAGGGTCAA | GTGCACAGTCTACAGTTAACGGGGTCAGTGCCTTGAGTAACAGTG |
| NS4-30nt-T21 | GATTAGAGCCGTCAATAGATAATACATTTGCCATTACAAGGTGTG | CTACCGGCCTGATAGCCCGTATAAACAGTTAATGCCCCCTGCCTA |
| NS4-30nt-T22 | AGGATTTAGAAGTATTAGACTTTACAAACACAGTTGCTGGTGCAT | GTAGAAGTTCAAAAGTTTCGGAACCTATTATTCTGAAACATGAAA |
| NS4-30nt-T23 | ATTCGACAACTCGTATTAAATCCTTTGCCCAGCAACCTGGTTAGA | AGTATTTGTTCCTGGGTATTAAGAGGCTGAGACTCCTCAAGAGAA |
| NS4-30nt-T24 | GAACGTTATTAATTTTAAAAGTTTGAGTAAACGCCAAGTAGGAGT | AAGTTGATCTGCATGGGATTAGGATTAGCGGGGTTTTGCTCAGTA |
| **Nanoswitch 5 30 nt targets** | |  |
| NS5-30nt-T1 | TCTGTCCATCACGCAAATTAACCGTTGTAGAGTCTGAGTCTGATA | ACTAGCGCATATACCTCAACCGATTGAGGGAGGGAAGGTAAATAT |
| NS5-30nt-T2 | CAATACTTCTTTGATTAGTAATAACATCACGTGACATAGTGTAGG | CAATGATGGATTGACTGACGGAAATTATTCATTAAAGGTGAATTA |
| NS5-30nt-T3 | TTGCCTGAGTAGAAGAACTCAAACTATCGGCCTCTTGCTTGGTTT | TGATGGATCTGGTAATCACCGTCACCGACTTGAGCCATTTGGGAA |
| NS5-30nt-T4 | CCTTGCTGGTAATATCCAGAACAATATTACGCTTGTGCATTTTGG | TTGACCACATCTTGATTAGAGCCAGCAAAATCACCAGTAGCACCA |
| NS5-30nt-T5 | CGCCAGCCATTGCAACAGGAAAAACGCTCACTCCTCCTTGAATGA | GTCTAATTCAGGTTGTTACCATTAGCAAGGCCGGAAACGTCACCA |
| NS5-30nt-T6 | TGGAAATACCTACATTTTGACGCTCAATCGGAGTGAGGCTTGTAT | CGGTATCGTTGCAGTATGAAACCATCGATAGCAGCACCGTAATCA |
| NS5-30nt-T7 | TCTGAAATGGATTATTTACATTGGCAGATTAGCAGCAACGAGCAA | AAGGTGTGAGTAAACGTAGCGACAGAATCAAGTTTGCCTTTAGCG |
| NS5-30nt-T8 | CACCAGTCACACGACCAGTAATAAAAGGGAAGGCACGCTAGTAGT | CGTCGTCGGTTCATCTCAGACTGTAGCGCGTTTTCATCGGCATTT |
| NS5-30nt-T9 | CATTCTGGCCAACAGAGATAGAACCCTTCTTTCTAGAAGCGGTCT | GGTCAGAATAGTGCCTCGGTCATAGCCCCCTTATTAGCGTTTGCC |
| NS5-30nt-T10 | GACCTGAAAGCGTAAGAATACGTGGCACAGTGTCCAGCAATACGA | AGATGTCCACGAAGGATCTTTTCATAATCAAAATCACCGGAACCA |
| NS5-30nt-T11 | ACAATATTTTTGAATGGCTATTAGTCTTTATAGCAACAGTGATTT | CTTTAGGCAGGTCCTGAGCCACCACCGGAACCGCCTCCCTCAGAG |
| NS5-30nt-T12 | ATGCGCGAACTGATAGCCCTAAAACATCGCCGTCAGGACAAGCAA | AAGCAAATTGAGTGCCCGCCACCCTCAGAACCGCCACCCTCAGAG |
| NS5-30nt-T13 | CATTAAAAATACCGAACGAACCACCAGCAGAGTGAATAGGACACG | GGTCATCAACTACATCCACCACCCTCAGAGCCGCCACCAGAACCA |
| NS5-30nt-T14 | AAGATAAAACAGAGGTGAGGCGGTCAGTATACCTTGGGGCCGACG | TTGTTTTGATCGCGCCCACCAGAGCCGCCGCCAGCATTGACAGGA |
| NS5-30nt-T15 | TAACACCGCCTGCAACAGTGCCACGCTGAGCTTCCTTGCCATGTT | GAGTGAGAGCGGTGAGGTTGAGGCAGGTCAGACGATTGGCCTTGA |
| NS5-30nt-T16 | AGCCAGCAGCAAATGAAAAATCTAAAGCATGGTAGTAGCCAATTT | GGTCATCTGGACTGCTATTCACAAACAAATAAATCCTCATTAAAG |
| NS5-30nt-T17 | CACCTTGCTGAACCTCAAATATCAAACCCTTGTTAGCACCATAGG | GAAGTCCAGCTTCTGCCAGAATGGAAAGCGCAGTCTCTGAATTTA |
| NS5-30nt-T18 | CAATCAATATCTGGTCAGTTGGCAAATCAACAGGATTGCGGGTGC | CAATGTGATCTTTTGCCGTTCCAGTAAGCGTCATACATGGCTTTT |
| NS5-30nt-T19 | CAGTTGAAAGGAATTGAGGAAGGTTATCTATTGCGACTACGTGAT | GAGGAACGAGAAGAGGATGATACAGGAGTGTACTGGTAATAAGTT |
| NS5-30nt-T20 | AAATATCTTTAGGAGCACTAACAACTAATACCTTGGGTTTGTTCT | GGACCACGTCTGCCGTTAACGGGGTCAGTGCCTTGAGTAACAGTG |
| NS5-30nt-T21 | GATTAGAGCCGTCAATAGATAATACATTTGGTTCCTTGTCTGATT | AGTTCCTGGTCCCCACCCGTATAAACAGTTAATGCCCCCTGCCTA |
| NS5-30nt-T22 | AGGATTTAGAAGTATTAGACTTTACAAACAGTTTCTTCTGTCTCT | GCGGTAAGGCTTGAGTTTCGGAACCTATTATTCTGAAACATGAAA |
| NS5-30nt-T23 | ATTCGACAACTCGTATTAAATCCTTTGCCCGTCTGCATGAGTTTA | GGCCTGAGTTGAGTCGTATTAAGAGGCTGAGACTCCTCAAGAGAA |
| NS5-30nt-T24 | GAACGTTATTAATTTTAAAAGTTTGAGTAAAGGCAGCTCTCCCTA | GCATTGTTCACTGTAGGATTAGGATTAGCGGGGTTTTGCTCAGTA |

|  | **Detector 1 sequence (nt 1-30 hybridize M13, 31-60 hybridize with target)** |
| --- | --- |
| **Nanoswitch 1 60 nt targets** | |
| NS1-60nt-T1 | TCTGTCCATCACGCAAATTAACCGTTGTAGACAGATTTTAAAGTTCGTTTAGAGAACAGA |
| NS1-60nt-T2 | CAATACTTCTTTGATTAGTAATAACATCACTGTAAAACAGGCAAACTGAGTTGGACGTGT |
| NS1-60nt-T3 | TTGCCTGAGTAGAAGAACTCAAACTATCGGCCAGTTTTCTTGAAAATCTTCATAAGGATC |
| NS1-60nt-T4 | CCTTGCTGGTAATATCCAGAACAATATTACCAACCCTTGGTTGAATAGTCTTGATTATGG |
| NS1-60nt-T5 | CGCCAGCCATTGCAACAGGAAAAACGCTCAACTTCTGAATTGTGACATGCTGGACAATAA |
| NS1-60nt-T6 | TGGAAATACCTACATTTTGACGCTCAATCGAACAATATTGATGTTGACTTTCTCTTTTTG |
| NS1-60nt-T7 | TCTGAAATGGATTATTTACATTGGCAGATTTTTCTGTTCACCAATATTCCAGGCACCTTT |
| NS1-60nt-T8 | CACCAGTCACACGACCAGTAATAAAAGGGACTTCAAGCCAATCAAGGACGGGTTTGAGTT |
| NS1-60nt-T9 | CATTCTGGCCAACAGAGATAGAACCCTTCTGATAATTTCTTTTGGGGCTTTTAGAGGCAT |
| NS1-60nt-T10 | GACCTGAAAGCGTAAGAATACGTGGCACAGACTTCTCATTAAGTACTTTATCAATCCTTT |
| NS1-60nt-T11 | ACAATATTTTTGAATGGCTATTAGTCTTTACTCATATTGAGTTGATGGCTCAAACTCTTC |
| NS1-60nt-T12 | ATGCGCGAACTGATAGCCCTAAAACATCGCGGTTGAACCTCAACAATTGTTTGAATAGTA |
| NS1-60nt-T13 | CATTAAAAATACCGAACGAACCACCAGCAGTTTAAGTGGTCCATTAGTAGCTATGTAATC |
| NS1-60nt-T14 | AAGATAAAACAGAGGTGAGGCGGTCAGTATAATACCAGCTGATAATAATGGTGCAAGTAG |
| NS1-60nt-T15 | TAACACCGCCTGCAACAGTGCCACGCTGAGCCTCTTTAGGAATCTCAGCGATCTTTTGTT |
| NS1-60nt-T16 | AGCCAGCAGCAAATGAAAAATCTAAAGCATATCTGGATGAAGATTGCCATTAATGTCAAT |
| NS1-60nt-T17 | CACCTTGCTGAACCTCAAATATCAAACCCTCGGGTAAGTGGTTATATAATTGTCTGTTGG |
| NS1-60nt-T18 | CAATCAATATCTGGTCAGTTGGCAAATCAACCTCTTGTATTTTAATACCCTTATATTTAC |
| NS1-60nt-T19 | CAGTTGAAAGGAATTGAGGAAGGTTATCTAAGCAAGTGAGATGGTTTCAATAAAATGTTC |
| NS1-60nt-T20 | AAATATCTTTAGGAGCACTAACAACTAATAATAAAATGTTTTACCTTCATGTGAATTATG |
| NS1-60nt-T21 | GATTAGAGCCGTCAATAGATAATACATTTGTGTAATAAGCATCTTGTAGAGCAGGTGGAT |
| NS1-60nt-T22 | AGGATTTAGAAGTATTAGACTTTACAAACACTTAAGGGTTGTCTGCTGTTGTCCACAAGT |
| NS1-60nt-T23 | ATTCGACAACTCGTATTAAATCCTTTGCCCCCGTCTATGCAATACAAAGTTTCTTTAGAA |
| NS1-60nt-T24 | GAACGTTATTAATTTTAAAAGTTTGAGTAATTGCGTTTGGATATGGTTGGTTTGGTACAA |
| **Nanoswitch 2 60 nt targets** | |
| NS2-60nt-T1 | TCTGTCCATCACGCAAATTAACCGTTGTAGATAGGTTTATGTAACAATTTAGCTCCTTTC |
| NS2-60nt-T2 | CAATACTTCTTTGATTAGTAATAACATCACATTAGATCTGTGTGGCCAACCTCTTCTGTA |
| NS2-60nt-T3 | TTGCCTGAGTAGAAGAACTCAAACTATCGGGTACACAATTGTAGCAATAAAGTAAAGAAA |
| NS2-60nt-T4 | CCTTGCTGGTAATATCCAGAACAATATTACGATTAAAGAACCTAGGCAAACACTTAATAG |
| NS2-60nt-T5 | CGCCAGCCATTGCAACAGGAAAAACGCTCACATTTAAAAGATGAAATGGTAATTTGTATA |
| NS2-60nt-T6 | TGGAAATACCTACATTTTGACGCTCAATCGCATACATAATAAAATGATGCAAAGAAGATG |
| NS2-60nt-T7 | TCTGAAATGGATTATTTACATTGGCAGATTCTGTAACACTATCAACGATGTAAGAAGACT |
| NS2-60nt-T8 | CACCAGTCACACGACCAGTAATAAAAGGGAGCTGATTTTGCAGATGATTCTTCACATTTT |
| NS2-60nt-T9 | CATTCTGGCCAACAGAGATAGAACCCTTCTAGTTGCAACTAGTGTTTTGAGTTTTTCCAT |
| NS2-60nt-T10 | GACCTGAAAGCGTAAGAATACGTGGCACAGCCAAGGTCACGGGGTGTCATGTTTTCAACT |
| NS2-60nt-T11 | ACAATATTTTTGAATGGCTATTAGTCTTTATGTTTAGACATGACATGAACAGGTGTTATT |
| NS2-60nt-T12 | ATGCGCGAACTGATAGCCCTAAAACATCGCACAAGCTGATGTTGCAAAGTCAGTGTACTC |
| NS2-60nt-T13 | CATTAAAAATACCGAACGAACCACCAGCAGTCAAGGTAGGTGTTAGGAAATTGAATAATA |
| NS2-60nt-T14 | AAGATAAAACAGAGGTGAGGCGGTCAGTATGTTAAACAGAGTACAGTGAATGACATAAGG |
| NS2-60nt-T15 | TAACACCGCCTGCAACAGTGCCACGCTGAGGGAAATACAAATGATATAAGCAATTGTTAT |
| NS2-60nt-T16 | AGCCAGCAGCAAATGAAAAATCTAAAGCATCTGAGGTGATAGAGGTTTGTGGTGGTTGGT |
| NS2-60nt-T17 | CACCTTGCTGAACCTCAAATATCAAACCCTTCTGTCCTGGTTGAATGCGAACAAACTTAT |
| NS2-60nt-T18 | CAATCAATATCTGGTCAGTTGGCAAATCAAGTAACAAAAAGAGACACAGTCATAATCTAT |
| NS2-60nt-T19 | CAGTTGAAAGGAATTGAGGAAGGTTATCTAAAAGAGGTCCTAGTATGTCAACATGGTCTT |
| NS2-60nt-T20 | AAATATCTTTAGGAGCACTAACAACTAATACCATAGCAAAAGGTAAAAAGGCATTTTCAT |
| NS2-60nt-T21 | GATTAGAGCCGTCAATAGATAATACATTTGTCATAAGGATTAGTAACACTACAGCTGATG |
| NS2-60nt-T22 | AGGATTTAGAAGTATTAGACTTTACAAACATGAAGAAAATAGGGCAATACTCAACACACA |
| NS2-60nt-T23 | ATTCGACAACTCGTATTAAATCCTTTGCCCGTTGCAAAACTGAGAGTAAGACTACTGATG |
| NS2-60nt-T24 | GAACGTTATTAATTTTAAAAGTTTGAGTAAGCAAAAGCTGCATATGATGGAAGGGAACTA |
| **Nanoswitch 3 60 nt targets** | |
| NS3-60nt-T1 | TCTGTCCATCACGCAAATTAACCGTTGTAGATGTTGTTGAGTGCATCATTATCCAACTTT |
| NS3-60nt-T2 | CAATACTTCTTTGATTAGTAATAACATCACCTTAAAGCTGTTACAATAAGAGGCCATGCT |
| NS3-60nt-T3 | TTGCCTGAGTAGAAGAACTCAAACTATCGGGTATAGATAGTACCAGTTCCATCACTCTTA |
| NS3-60nt-T4 | CCTTGCTGGTAATATCCAGAACAATATTACTAAGCTTTAGCAGCATCTACAGCAAAAGCA |
| NS3-60nt-T5 | CGCCAGCCATTGCAACAGGAAAAACGCTCACAGACTGTGTTTTTAAGTGTAAAACCCACA |
| NS3-60nt-T6 | TGGAAATACCTACATTTTGACGCTCAATCGCCTTTTCTTGGAAGCGACAACAATTAGTTT |
| NS3-60nt-T7 | TCTGAAATGGATTATTTACATTGGCAGATTTAAAGCATAGACGAGGTCTGCCATTGTGTA |
| NS3-60nt-T8 | CACCAGTCACACGACCAGTAATAAAAGGGAGTATACGCGTAATATATCTGGGTTTTCTAC |
| NS3-60nt-T9 | CATTCTGGCCAACAGAGATAGAACCCTTCTCAAGGTTAATATAGGCATTAACAATGAATA |
| NS3-60nt-T10 | GACCTGAAAGCGTAAGAATACGTGGCACAGTGGTCCAAAACTTGTAGGTGGGAACACTGT |
| NS3-60nt-T11 | ACAATATTTTTGAATGGCTATTAGTCTTTACATAGAAGTCTTTGTTAAAATTACCGGGTT |
| NS3-60nt-T12 | ATGCGCGAACTGATAGCCCTAAAACATCGCAATGGAAAACCAGCTGATTTGTCTAGGTTG |
| NS3-60nt-T13 | CATTAAAAATACCGAACGAACCACCAGCAGCGAGCTCTATTCTTTGCACTAATGGCATAC |
| NS3-60nt-T14 | AAGATAAAACAGAGGTGAGGCGGTCAGTATCACGTTGTATGTTTGCGAGCAAGAACAAGT |
| NS3-60nt-T15 | TAACACCGCCTGCAACAGTGCCACGCTGAGTATTGAAACACACAACAGCATCGTCAGAGA |
| NS3-60nt-T16 | AGCCAGCAGCAAATGAAAAATCTAAAGCATACAAAACAGCCGGCCCCTAGGATTCTTGAT |
| NS3-60nt-T17 | CACCTTGCTGAACCTCAAATATCAAACCCTTGTCTAACATGTGTCCTGTTAACTCATCAT |
| NS3-60nt-T18 | CAATCAATATCTGGTCAGTTGGCAAATCAACGGATTAACAGACAAGACTAATTTATGTGA |
| NS3-60nt-T19 | CAGTTGAAAGGAATTGAGGAAGGTTATCTAGCTTGAGTCTTTCAGTACAGGTGTTAGCTA |
| NS3-60nt-T20 | AAATATCTTTAGGAGCACTAACAACTAATATGTTTTTAGTTACACGATAACCAGTAAAGA |
| NS3-60nt-T21 | GATTAGAGCCGTCAATAGATAATACATTTGCAAGCTGTATACACTATGCGAGCAGAAGGG |
| NS3-60nt-T22 | AGGATTTAGAAGTATTAGACTTTACAAACAATTGAAATATTCTGGTTCTAGTGTGCCCTT |
| NS3-60nt-T23 | ATTCGACAACTCGTATTAAATCCTTTGCCCGTGAATTATAAGGTGAAATAAAGACAGCTT |
| NS3-60nt-T24 | GAACGTTATTAATTTTAAAAGTTTGAGTAATACTACAATCTTTAAAGAGTCCTGTTACAT |
| **Nanoswitch 4 60 nt targets** | |
| NS4-60nt-T1 | TCTGTCCATCACGCAAATTAACCGTTGTAGGCTTCTTCGCGGGTGATAAACATGTTAGGG |
| NS4-60nt-T2 | CAATACTTCTTTGATTAGTAATAACATCACAAGTCCTTTGTACATAAGTGGTATGAGGTG |
| NS4-60nt-T3 | TTGCCTGAGTAGAAGAACTCAAACTATCGGTCACATAGACAACAGGTGCGCTCAGGTCCT |
| NS4-60nt-T4 | CCTTGCTGGTAATATCCAGAACAATATTACTTAATCTTCAGTTCATCACCAATTATAGGA |
| NS4-60nt-T5 | CGCCAGCCATTGCAACAGGAAAAACGCTCATGAATTTGTCAGAATGTGTGGCATAAGAAT |
| NS4-60nt-T6 | TGGAAATACCTACATTTTGACGCTCAATCGTCACATGGACTGTCAGAGTAATAGAAAAAT |
| NS4-60nt-T7 | TCTGAAATGGATTATTTACATTGGCAGATTGTCTTGTAAAAGTGTTCCAGAGGTTATAAG |
| NS4-60nt-T8 | CACCAGTCACACGACCAGTAATAAAAGGGACAAAAAAGACAGTGAGTGGTGCACAAATCG |
| NS4-60nt-T9 | CATTCTGGCCAACAGAGATAGAACCCTTCTGTTTAAATTCTTGTAAATTTCTACTCTGAG |
| NS4-60nt-T10 | GACCTGAAAGCGTAAGAATACGTGGCACAGGCGCATCTGTTATGAAATAGTTTTTAACTG |
| NS4-60nt-T11 | ACAATATTTTTGAATGGCTATTAGTCTTTACCACGCTTGACTAGATTGTAATTTTGGGTA |
| NS4-60nt-T12 | ATGCGCGAACTGATAGCCCTAAAACATCGCCCAAAATGTATAACTCTCATATTATAGGGT |
| NS4-60nt-T13 | CATTAAAAATACCGAACGAACCACCAGCAGCTTTAGAGTCATTTTCTTTTGTAACATTTT |
| NS4-60nt-T14 | AAGATAAAACAGAGGTGAGGCGGTCAGTATTTTGTATTCCTCCAAAATATGTAATTTGCA |
| NS4-60nt-T15 | TAACACCGCCTGCAACAGTGCCACGCTGAGCCACGTGTGAAAGAATTAGTGTATGCAGGG |
| NS4-60nt-T16 | AGCCAGCAGCAAATGAAAAATCTAAAGCATCAACATTAGTAGCGTTATTAACAATAAGTA |
| NS4-60nt-T17 | CACCTTGCTGAACCTCAAATATCAAACCCTCACTAAATTAATAGGCGTGTGCTTAGAATA |
| NS4-60nt-T18 | CAATCAATATCTGGTCAGTTGGCAAATCAAACAGCATCTGTAATGGTTCCATTTTCATTA |
| NS4-60nt-T19 | CAGTTGAAAGGAATTGAGGAAGGTTATCTAGCATAGACATTAGTAAAGCAGAGATCATTT |
| NS4-60nt-T20 | AAATATCTTTAGGAGCACTAACAACTAATACCAACACCATTAGTGGGTTGGAAACCATAT |
| NS4-60nt-T21 | GATTAGAGCCGTCAATAGATAATACATTTGTCTGTGGATCACGGACAGCATCAGTAGTGT |
| NS4-60nt-T22 | AGGATTTAGAAGTATTAGACTTTACAAACAGTTTGAAAAACATTAGAACCTGTAGAATAA |
| NS4-60nt-T23 | ATTCGACAACTCGTATTAAATCCTTTGCCCGGTAGAATTTCTGTGGTAACACTAATAGTA |
| NS4-60nt-T24 | GAACGTTATTAATTTTAAAAGTTTGAGTAAATCTTTAATTGGTGGTGTTTTGTAAATTTG |
| **Nanoswitch 5 60 nt targets** | |
| NS5-60nt-T1 | TCTGTCCATCACGCAAATTAACCGTTGTAGGAAGTGTATTGAGCAATCATTTCATCTGTG |
| NS5-60nt-T2 | CAATACTTCTTTGATTAGTAATAACATCACACTTTGTCAAGACGTGAAAGGATATCATTT |
| NS5-60nt-T3 | TTGCCTGAGTAGAAGAACTCAAACTATCGGTTTTCCATCATGACAAATGGCAGGAGCAGT |
| NS5-60nt-T4 | CCTTGCTGGTAATATCCAGAACAATATTACTGAATGAGTCTAATTCAGGTTGCAAAGGAT |
| NS5-60nt-T5 | CGCCAGCCATTGCAACAGGAAAAACGCTCAAAGTTCTTGGAGATCGATGAGAGATTCATT |
| NS5-60nt-T6 | TGGAAATACCTACATTTTGACGCTCAATCGTCCTTGATTTCACCTTGCTTCAAAGTTACA |
| NS5-60nt-T7 | TCTGAAATGGATTATTTACATTGGCAGATTCATAAAGATAGAGAAAAGGGGCTTCAAGGC |
| NS5-60nt-T8 | CACCAGTCACACGACCAGTAATAAAAGGGACAATAGTCGTAACAATTAGTATGCCAGCAA |
| NS5-60nt-T9 | CATTCTGGCCAACAGAGATAGAACCCTTCTACAATTTTATTGTAGATGAAGAAGGTAACA |
| NS5-60nt-T10 | GACCTGAAAGCGTAAGAATACGTGGCACAGTAGTGTAACTAGCAAGAATACCACGAAAGC |
| NS5-60nt-T11 | ACAATATTTTTGAATGGCTATTAGTCTTTACGTTGGAATCTGCCATGGCTAAAATTAAAG |
| NS5-60nt-T12 | ATGCGCGAACTGATAGCCCTAAAACATCGCCATGGAGTGGCACGTTGAGAAGAATGTTAG |
| NS5-60nt-T13 | CATTAAAAATACCGAACGAACCACCAGCAGCCCAATTTGTAATAAGAAAGCGTTCGTGAT |
| NS5-60nt-T14 | AAGATAAAACAGAGGTGAGGCGGTCAGTATCTCAGTTAGTGACTTAGATAAATTTTTAAT |
| NS5-60nt-T15 | TAACACCGCCTGCAACAGTGCCACGCTGAGAATTGAGTGCTAAAGCAAGTCAGTGCAAAT |
| NS5-60nt-T16 | AGCCAGCAGCAAATGAAAAATCTAAAGCATAAGATAATAAGCATAATTAAAACAAGGAAT |
| NS5-60nt-T17 | CACCTTGCTGAACCTCAAATATCAAACCCTTAAAAGGTAAACAGGAAACTGTATAATTAC |
| NS5-60nt-T18 | CAATCAATATCTGGTCAGTTGGCAAATCAACTTGCCATGTTGAGTGAGAGCGGTGAACCA |
| NS5-60nt-T19 | CAGTTGAAAGGAATTGAGGAAGGTTATCTATAGAAATACCATCTTGGACTGAGATCTTTC |
| NS5-60nt-T20 | AAATATCTTTAGGAGCACTAACAACTAATACCTGGAGTTGAATTTCTTGAACTGTTGCGA |
| NS5-60nt-T21 | GATTAGAGCCGTCAATAGATAATACATTTGCAAAATTTCCTTGGGTTTGTTCTGGACCAC |
| NS5-60nt-T22 | AGGATTTAGAAGTATTAGACTTTACAAACACTTCTTCTTTTTGTCCTTTTTAGGCTCTGT |
| NS5-60nt-T23 | ATTCGACAACTCGTATTAAATCCTTTGCCCGAGAATTCATTCTGCACAAGAGTAGACTAT |
| NS5-60nt-T24 | GAACGTTATTAATTTTAAAAGTTTGAGTAAGCTATTAAAATCACATGGGGATAGCACTAC |

|  | **Detector 2 sequence (nt 1-30 hybridize with target, 31-60 hybridizing M13)** |
| --- | --- |
| **Nanoswitch 1 60 nt targets** | |
| NS1-60nt-T1 | TCTACAAGAGATCGAAAGTTGGTTGGTTTGTCAACCGATTGAGGGAGGGAAGGTAAATAT |
| NS1-60nt-T2 | GTTTTCTCGTTGAAACCAGGGACAAGGCTCTGACGGAAATTATTCATTAAAGGTGAATTA |
| NS1-60nt-T3 | AGTGCCAAGCTCGTCGCCTAAGTCAAATGATCACCGTCACCGACTTGAGCCATTTGGGAA |
| NS1-60nt-T4 | AATTTAAGGGAAATACAAAATTTGGACATTTTAGAGCCAGCAAAATCACCAGTAGCACCA |
| NS1-60nt-T5 | ATTTTAACAACAGCATTTTGGGGTAAGTAATTACCATTAGCAAGGCCGGAAACGTCACCA |
| NS1-60nt-T6 | GAGTATTTCAAGAAGGTTGTCATTAAGACCATGAAACCATCGATAGCAGCACCGTAATCA |
| NS1-60nt-T7 | TTTAGCTTTTCCTTTTGTAACTTTAAAATTGTAGCGACAGAATCAAGTTTGCCTTTAGCG |
| NS1-60nt-T8 | TTTCATAAACAGTGCCAAAGATGTTAGTTATCAGACTGTAGCGCGTTTTCATCGGCATTT |
| NS1-60nt-T9 | GAGTAGGCCAGTTTCTTCTCTGGATTTAACTCGGTCATAGCCCCCTTATTAGCGTTTGCC |
| NS1-60nt-T10 | CATCAAGTTCAAAAGTGATATTCACACTCTATCTTTTCATAATCAAAATCACCGGAACCA |
| NS1-60nt-T11 | TTCTTCACAATCACCTTCTTCTTCATCCTCGAGCCACCACCGGAACCGCCTCCCTCAGAG |
| NS1-60nt-T12 | GTTGTCTGATTGTCCTCACTGCCGTCTTGTCCGCCACCCTCAGAACCGCCACCCTCAGAG |
| NS1-60nt-T13 | ATCAGATTCAACTTGCATGGCATTGTTAGTCCACCACCCTCAGAGCCGCCACCAGAACCA |
| NS1-60nt-T14 | AACTTCGTGCTGATTAAAATTTTCATAAGCCCACCAGAGCCGCCGCCAGCATTGACAGGA |
| NS1-60nt-T15 | CAACTTGCTTTTCACTCTTCATTTCCAAAAGGTTGAGGCAGGTCAGACGATTGGCCTTGA |
| NS1-60nt-T16 | ATAAAGTAACAAGTTTTCTGTGAGGAACTTTATTCACAAACAAATAAATCCTCATTAAAG |
| NS1-60nt-T17 | CACTTTTCTCAAAGCTTTCGCTAGCATTTCCCAGAATGGAAAGCGCAGTCTCTGAATTTA |
| NS1-60nt-T18 | GCTGTATAGTTGAAACTATGGCTTTAGTTTCCGTTCCAGTAAGCGTCATACATGGCTTTT |
| NS1-60nt-T19 | TTCAGGTGTTTTAGAAGAAGAAGTAAGATAGATGATACAGGAGTGTACTGGTAATAAGTT |
| NS1-60nt-T20 | AGGTTTTATTTTAGTAACATCAGCTCCATCTTAACGGGGTCAGTGCCTTGAGTAACAGTG |
| NS1-60nt-T21 | TAAACTTCAACTCTATTTGTTGGAGTGTTACCCGTATAAACAGTTAATGCCCCCTGCCTA |
| NS1-60nt-T22 | TTTACACACCACGTTCAAGACTCTTTTGCATTTCGGAACCTATTATTCTGAAACATGAAA |
| NS1-60nt-T23 | GTTATATGTTTATAGTGACCACACTGGTAAGTATTAAGAGGCTGAGACTCCTCAAGAGAA |
| NS1-60nt-T24 | GATCAATTGGTTGCTCTGTGAAATAAGAATGGATTAGGATTAGCGGGGTTTTGCTCAGTA |
| **Nanoswitch 2 60 nt targets** | |
| NS2-60nt-T1 | TTAAAAGAGGGTGTGTAGTGTTTATAATCATCAACCGATTGAGGGAGGGAAGGTAAATAT |
| NS2-60nt-T2 | ATTTTTAAACTATTATTTGCTGGTTTAAGTTGACGGAAATTATTCATTAAAGGTGAATTA |
| NS2-60nt-T3 | TAAGGCATATAATTAGTACAAACACGGTTTTCACCGTCACCGACTTGAGCCATTTGGGAA |
| NS2-60nt-T4 | TAAAAACCAAATTATAATATTTATCAGTTTTTAGAGCCAGCAAAATCACCAGTAGCACCA |
| NS2-60nt-T5 | GTTTCTAAAGAAGGATAGGTGTCTAAAGAATTACCATTAGCAAGGCCGGAAACGTCACCA |
| NS2-60nt-T6 | TACATTCTAACCATAGCTGAAATCGGGGCCATGAAACCATCGATAGCAGCACCGTAATCA |
| NS2-60nt-T7 | GGTCAGTAGGATTTATTGGTCTTTTAAACTGTAGCGACAGAATCAAGTTTGCCTTTAGCG |
| NS2-60nt-T8 | GATTTACCATCAAAAACTATAACATTAATATCAGACTGTAGCGCGTTTTCATCGGCATTT |
| NS2-60nt-T9 | TGGTACGTTAAAAGTTGATGAAAACGTATTTCGGTCATAGCCCCCTTATTAGCGTTTGCC |
| NS2-60nt-T10 | TTGTTATAGGTGAGCATATAGTTATTACAAATCTTTTCATAATCAAAATCACCGGAACCA |
| NS2-60nt-T11 | AAATAGAAAATAGCAGCAACAAAAAGGAACGAGCCACCACCGGAACCGCCTCCCTCAGAG |
| NS2-60nt-T12 | TATAAGTTTTGATGGTGTGTAACAGATGTTCCGCCACCCTCAGAACCGCCACCCTCAGAG |
| NS2-60nt-T13 | GAGCCATCCATGAGCACATAACGTGTGTCACCACCACCCTCAGAGCCGCCACCAGAACCA |
| NS2-60nt-T14 | AATAGTAAAGTATTAAAGGCAACTACATGACCACCAGAGCCGCCGCCAGCATTGACAGGA |
| NS2-60nt-T15 | CCAGAAAGGTACTAAAGGTGTGAACATAACGGTTGAGGCAGGTCAGACGATTGGCCTTGA |
| NS2-60nt-T16 | AAAGAACATCAGAACCTGAGTTACTGAAGTTATTCACAAACAAATAAATCCTCATTAAAG |
| NS2-60nt-T17 | ACTTAGGTGTCTTAGGATTGGCTGTATCAACCAGAATGGAAAGCGCAGTCTCTGAATTTA |
| NS2-60nt-T18 | GTTAAAACCAACACTACCACATGAACCATTCCGTTCCAGTAAGCGTCATACATGGCTTTT |
| NS2-60nt-T19 | GTGTTAGAGGTTCATAATTGTACTTCATAGGATGATACAGGAGTGTACTGGTAATAAGTT |
| NS2-60nt-T20 | ACAAAAAAAAGAACAAAGACCATTGAGTACTTAACGGGGTCAGTGCCTTGAGTAACAGTG |
| NS2-60nt-T21 | CATACATAACACAGTCTTTTAGCTTAAAACCCCGTATAAACAGTTAATGCCCCCTGCCTA |
| NS2-60nt-T22 | TAAAAACAATACCTCTGGCCAAAAACATGATTTCGGAACCTATTATTCTGAAACATGAAA |
| NS2-60nt-T23 | TGCACTTTACATCTGACATTTTAGACTGTAGTATTAAGAGGCTGAGACTCCTCAAGAGAA |
| NS2-60nt-T24 | AACTCTGAGGCTATAGCTTGTAAGGTTGCCGGATTAGGATTAGCGGGGTTTTGCTCAGTA |
| **Nanoswitch 3 60 nt targets** | |
| NS3-60nt-T1 | CTAAGCATAGTGAAAAGCATTGTCTGCATATCAACCGATTGAGGGAGGGAAGGTAAATAT |
| NS3-60nt-T2 | AAATTAGGTGAATTGTCCATACTAATTTCATGACGGAAATTATTCATTAAAGGTGAATTA |
| NS3-60nt-T3 | GGGAATCTAGCCCATTTCAAATCCTGTAAATCACCGTCACCGACTTGAGCCATTTGGGAA |
| NS3-60nt-T4 | CAGAAAGATAATACAGTTGAATTGGCAGGCTTAGAGCCAGCAAAATCACCAGTAGCACCA |
| NS3-60nt-T5 | GGGTCATTAGCACAAGTTGTAGGTATTTGTTTACCATTAGCAAGGCCGGAAACGTCACCA |
| NS3-60nt-T6 | TTAGGAATTTAGCAAAACCAGCTACTTTATATGAAACCATCGATAGCAGCACCGTAATCA |
| NS3-60nt-T7 | TTTAGTAAGACGTTGACGTGATATATGTGGGTAGCGACAGAATCAAGTTTGCCTTTAGCG |
| NS3-60nt-T8 | AAAATCATACCAGTCCTTTTTATTGAAATATCAGACTGTAGCGCGTTTTCATCGGCATTT |
| NS3-60nt-T9 | ATAAGAATCTACAACAGGAACTCCACTACCTCGGTCATAGCCCCCTTATTAGCGTTTGCC |
| NS3-60nt-T10 | AGAGAATAAAACATTAAAGTTTGCACAATGATCTTTTCATAATCAAAATCACCGGAACCA |
| NS3-60nt-T11 | TGACAGTTTGAAAAGCAACATTGTTAGTAAGAGCCACCACCGGAACCGCCTCCCTCAGAG |
| NS3-60nt-T12 | TTGACGATGACTTGGTTAGCATTAATACAGCCGCCACCCTCAGAACCGCCACCCTCAGAG |
| NS3-60nt-T13 | TTAAGATTCATTTGAGTTATAGTAGGGATGCCACCACCCTCAGAGCCGCCACCAGAACCA |
| NS3-60nt-T14 | GAGGCCATAATTCTAAGCATGTTAGGCATGCCACCAGAGCCGCCGCCAGCATTGACAGGA |
| NS3-60nt-T15 | GTATCATCATTGAGAAATGTTTACGCAAATGGTTGAGGCAGGTCAGACGATTGGCCTTGA |
| NS3-60nt-T16 | GGATCTGGGTAAGGAAGGTACACATAATCATATTCACAAACAAATAAATCCTCATTAAAG |
| NS3-60nt-T17 | GTAGCTTTCTTATGTATTGTAAGTACAAATCCAGAATGGAAAGCGCAGTCTCTGAATTTA |
| NS3-60nt-T18 | TGTTGATATGACATGGTCGTAACAGCATTTCCGTTCCAGTAAGCGTCATACATGGCTTTT |
| NS3-60nt-T19 | AAATGTAATCACCAGCATTTGTCCAGTCACGATGATACAGGAGTGTACTGGTAATAAGTT |
| NS3-60nt-T20 | CATAATTTCGGTTAAGTGGTGGTCTAGGTTTTAACGGGGTCAGTGCCTTGAGTAACAGTG |
| NS3-60nt-T21 | TAGTAGAGAGCTAGGCCAATAGCAAAATGACCCGTATAAACAGTTAATGCCCCCTGCCTA |
| NS3-60nt-T22 | AGTTAGCAATGTGCGTGGTGCAGGTAATTGTTTCGGAACCTATTATTCTGAAACATGAAA |
| NS3-60nt-T23 | TTCTCCAAGCAGGGTTACGTGTAAGGAATTGTATTAAGAGGCTGAGACTCCTCAAGAGAA |
| NS3-60nt-T24 | TTTCAGCTTGTAAAGTTGCCACATTCCTACGGATTAGGATTAGCGGGGTTTTGCTCAGTA |
| **Nanoswitch 4 60 nt targets** | |
| NS4-60nt-T1 | TAACCATTAACTTGATAATTCATTTTAAAATCAACCGATTGAGGGAGGGAAGGTAAATAT |
| NS4-60nt-T2 | TTTAAATTGATCTCCAGGCGGTGGTTTAGCTGACGGAAATTATTCATTAAAGGTGAATTA |
| NS4-60nt-T3 | ATTTTCACAAAATACTTCATAGATGTCAACTCACCGTCACCGACTTGAGCCATTTGGGAA |
| NS4-60nt-T4 | TATTCAATAGTCCAGTCAACACGCTTAACATTAGAGCCAGCAAAATCACCAGTAGCACCA |
| NS4-60nt-T5 | AGAATAATTCTTCTATTTTATAAGCTTTGTTTACCATTAGCAAGGCCGGAAACGTCACCA |
| NS4-60nt-T6 | GGTAATTGTTTTAAATTAACAAAAGCACTTATGAAACCATCGATAGCAGCACCGTAATCA |
| NS4-60nt-T7 | TATCAAATTGTTTGTAAACCCACAAGCTAAGTAGCGACAGAATCAAGTTTGCCTTTAGCG |
| NS4-60nt-T8 | TTTCAGTTGGTTTCTTGGCTATGTCAGTCATCAGACTGTAGCGCGTTTTCATCGGCATTT |
| NS4-60nt-T9 | TAAAGTAAGTTTCAGGTAATTGTTGGACAATCGGTCATAGCCCCCTTATTAGCGTTTGCC |
| NS4-60nt-T10 | TACTGTCCATAGGAATAAAATCTTCTAATTATCTTTTCATAATCAAAATCACCGGAACCA |
| NS4-60nt-T11 | AAATGTTTCTACATGGCCATCTTTACACCAGAGCCACCACCGGAACCGCCTCCCTCAGAG |
| NS4-60nt-T12 | ACAGCTAATGTTAATGTGTTTAAATATTGACCGCCACCCTCAGAACCGCCACCCTCAGAG |
| NS4-60nt-T13 | TAGTCTTAGGGTCGTACATATCACTAATAACCACCACCCTCAGAGCCGCCACCAGAACCA |
| NS4-60nt-T14 | TGCATGACATAACCATCTATTTGTTCGCGTCCACCAGAGCCGCCGCCAGCATTGACAGGA |
| NS4-60nt-T15 | GGTAATTGAGTTCTGGTTGTAAGATTAACAGGTTGAGGCAGGTCAGACGATTGGCCTTGA |
| NS4-60nt-T16 | GGGACTGGGTCTTCGAATCTAAAGTAGTACTATTCACAAACAAATAAATCCTCATTAAAG |
| NS4-60nt-T17 | TATTTTAAAATAACCATCAATATTCTTAAACCAGAATGGAAAGCGCAGTCTCTGAATTTA |
| NS4-60nt-T18 | TATTTTAATAGAAAAGTCCTAGGTTGAAGACCGTTCCAGTAAGCGTCATACATGGCTTTT |
| NS4-60nt-T19 | AATTTAGTAGGAGACACTCCATAACACTTAGATGATACAGGAGTGTACTGGTAATAAGTT |
| NS4-60nt-T20 | GATTGTAAAGGAAAGTAACAATTAAAACCTTTAACGGGGTCAGTGCCTTGAGTAACAGTG |
| NS4-60nt-T21 | CAGCAATGTCTCTGCCAAATTGTTGGAAAGCCCGTATAAACAGTTAATGCCCCCTGCCTA |
| NS4-60nt-T22 | ACACGCCAAGTAGGAGTAAGTTGATCTGCATTTCGGAACCTATTATTCTGAAACATGAAA |
| NS4-60nt-T23 | AAATTTGTGGGTATGGCAATAGAGTTATTAGTATTAAGAGGCTGAGACTCCTCAAGAGAA |
| NS4-60nt-T24 | TTTGACTTGTGCAAAAACTTCTTGGGTGTTGGATTAGGATTAGCGGGGTTTTGCTCAGTA |
| **Nanoswitch 5 60 nt targets** | |
| NS5-60nt-T1 | AGCAAAGGTGGCAAAACAGTAAGGCCGTTATCAACCGATTGAGGGAGGGAAGGTAAATAT |
| NS5-60nt-T2 | AAAACACTTGAAATTGCACCAAAATTGGAGTGACGGAAATTATTCATTAAAGGTGAATTA |
| NS5-60nt-T3 | TGTGAAGTTCTTTTCTTGTGCAGGGACATATCACCGTCACCGACTTGAGCCATTTGGGAA |
| NS5-60nt-T4 | CATAAACTGTGTTGTTGACAATTCCTATTATTAGAGCCAGCAAAATCACCAGTAGCACCA |
| NS5-60nt-T5 | TAAATTCTTGGCAACCTCATTGAGGCGGTCTTACCATTAGCAAGGCCGGAAACGTCACCA |
| NS5-60nt-T6 | GTTCCAATTGTGAAGATTCTCATAAACAAAATGAAACCATCGATAGCAGCACCGTAATCA |
| NS5-60nt-T7 | CAGCAGCAACGAGCAAAAGGTGTGAGTAAAGTAGCGACAGAATCAAGTTTGCCTTTAGCG |
| NS5-60nt-T8 | AGAAAATAGTTGGCATCATAAAGTAATGGGTCAGACTGTAGCGCGTTTTCATCGGCATTT |
| NS5-60nt-T9 | TGTTCAACACCAGTGTCTGTACTCAATTGATCGGTCATAGCCCCCTTATTAGCGTTTGCC |
| NS5-60nt-T10 | AAGAAAAAGAAGTACGCTATTAACTATTAAATCTTTTCATAATCAAAATCACCGGAACCA |
| NS5-60nt-T11 | TTCCAAACAGAAAAACTAATATAATATTTAGAGCCACCACCGGAACCGCCTCCCTCAGAG |
| NS5-60nt-T12 | TTTCTGGATTGAATGACCACATGGAACGCGCCGCCACCCTCAGAACCGCCACCCTCAGAG |
| NS5-60nt-T13 | GTAGCAACAGTGATTTCTTTAGGCAGGTCCCCACCACCCTCAGAGCCGCCACCAGAACCA |
| NS5-60nt-T14 | TATGAGGTTTATGATGTAATCAAGATTCCACCACCAGAGCCGCCGCCAGCATTGACAGGA |
| NS5-60nt-T15 | TTGTTATCAGCTAGAGGATGAAATGGTGAAGGTTGAGGCAGGTCAGACGATTGGCCTTGA |
| NS5-60nt-T16 | AGCAGAAAGGCTAAAAAGCACAAATAGAAGTATTCACAAACAAATAAATCCTCATTAAAG |
| NS5-60nt-T17 | CGATATCGATGTACTGAATGGGTGATTTAGCCAGAATGGAAAGCGCAGTCTCTGAATTTA |
| NS5-60nt-T18 | AGACGCAGTATTATTGGGTAAACCTTGGGGCCGTTCCAGTAAGCGTCATACATGGCTTTT |
| NS5-60nt-T19 | ATTTTACCGTCACCACCACGAATTCGTCTGGATGATACAGGAGTGTACTGGTAATAAGTT |
| NS5-60nt-T20 | CTACGTGATGAGGAACGAGAAGAGGCTTGATTAACGGGGTCAGTGCCTTGAGTAACAGTG |
| NS5-60nt-T21 | GTCTGCCGAAAGCTTGTGTTACATTGTATGCCCGTATAAACAGTTAATGCCCCCTGCCTA |
| NS5-60nt-T22 | TGGTGGGAATGTTTTGTATGCGTCAATATGTTTCGGAACCTATTATTCTGAAACATGAAA |
| NS5-60nt-T23 | ATATCGTAAACGGAAAAGCGAAAACGTTTAGTATTAAGAGGCTGAGACTCCTCAAGAGAA |
| NS5-60nt-T24 | TAAAATTAATTTTACACATTAGGGCTCTTCGGATTAGGATTAGCGGGGTTTTGCTCAGTA |

***Table S3 – Tiling oligo sequences***

| **Tiling oligos for 24 target nanoswitches (base set)** | |
| --- | --- |
| Var 1 | AACATCCAATAAATCATACAGGCAAGGCAAAGAATTAGCAAAATTAAGCAATAAAGCCTC |
| 1.01 | AGAGCATAAAGCTAAATCGGTTGTACCAAAAACATTATGACCCTGTAATACTTTTGCGGG |
| 1.02 | AGAAGCCTTTATTTCAACGCAAGGATAAAAATTTTTAGAACCCTCATATATTTTAAATGC |
| 1.03 | AATGCCTGAGTAATGTGTAGGTAAAGATTCAAAAGGGTGAGAAAGGCCGGAGACAGTCAA |
| 1.04 | ATCACCATCAATATGATATTCAACCGTTCTAGCTGATAAATTAATGCCGGAGAGGGTAGC |
| 1.05 | TATTTTTGAGAGATCTACAAAGGCTATCAGGTCATTGCCTGAGAGTCTGGAGCAAACAAG |
| 1.06 | AGAATCGATGAACGGTAATCGTAAAACTAGCATGTCAATCATATGTACCCCGGTTGATAA |
| 1.07 | TCAGAAAAGCCCCAAAAACAGGAAGATTGTATAAGCAAATATTTAAATTGTAAACGTTAA |
| 1.08 | TATTTTGTTAAAATTCGCATTAAATTTTTGTTAAATCAGCTCATTTTTTAACCAATAGGA |
| 1.09 | ACGCCATCAAAAATAATTCGCGTCTGGCCTTCCTGTAGCCAGCTTTCATCAACATTAAAT |
| Var 2 | GTGAGCGAGTAACAACCCGTCGGATTCTCCGTGGGAACAAACGGCGGATTGACCGTAATG |
| 2.01 | GGATAGGTCACGTTGGTGTAGATGGGCGCATCGTAACCGTGCATCTGCCAGTTTGAGGGG |
| 2.02 | ACGACGACAGTATCGGCCTCAGGAAGATCGCACTCCAGCCAGCTTTCCGGCACCGCTTCT |
| 2.03 | GGTGCCGGAAACCAGGCAAAGCGCCATTCGCCATTCAGGCTGCGCAACTGTTGGGAAGGG |
| 2.04 | CGATCGGTGCGGGCCTCTTCGCTATTACGCCAGCTGGCGAAAGGGGGATGTGCTGCAAGG |
| 2.05 | CGATTAAGTTGGGTAACGCCAGGGTTTTCCCAGTCACGACGTTGTAAAACGACGGCCAGT |
| 2.06 | GCCAAGCTTGCATGCCTGCAGGTCGACTCTAGAGGATCCCCGGGTACCGAGCTCGAATTC |
| 2.07 | GTAATCATGGTCATAGCTGTTTCCTGTGTGAAATTGTTATCCGCTCACAATTCCACACAA |
| 2.08 | CATACGAGCCGGAAGCATAAAGTGTAAAGCCTGGGGTGCCTAATGAGTGAGCTAACTCAC |
| 2.09 | ATTAATTGCGTTGCGCTCACTGCCCGCTTTCCAGTCGGGAAACCTGTCGTGCCAGCTGCA |
| 2.10 | TTAATGAATCGGCCAACGCGCGGGGAGAGGCGGTTTGCGTATTGGGCGCCAGGGTGGTTT |
| Var 3 | TTCTTTTCACCAGTGAGACGGGCAACAGCTGATTGCCCTTCACCGCCTGGCCCTGAGAGA |
| 3.01 | GTTGCAGCAAGCGGTCCACGCTGGTTTGCCCCAGCAGGCGAAAATCCTGTTTGATGGTGG |
| 3.02 | TTCCGAAATCGGCAAAATCCCTTATAAATCAAAAGAATAGCCCGAGATAGGGTTGAGTGT |
| 3.03 | TGTTCCAGTTTGGAACAAGAGTCCACTATTAAAGAACGTGGACTCCAACGTCAAAGGGCG |
| 3.04 | AAAAACCGTCTATCAGGGCGATGGCCCACTACGTGAACCATCACCCAAATCAAGTTTTTT |
| 3.05 | GGGGTCGAGGTGCCGTAAAGCACTAAATCGGAACCCTAAAGGGAGCCCCCGATTTAGAGC |
| 3.06 | TTGACGGGGAAAGCCGGCGAACGTGGCGAGAAAGGAAGGGAAGAAAGCGAAAGGAGCGGG |
| 3.07 | CGCTAGGGCGCTGGCAAGTGTAGCGGTCACGCTGCGCGTAACCACCACACCCGCCGCGCT |
| 3.08 | TAATGCGCCGCTACAGGGCGCGTACTATGGTTGCTTTGACGAGCACGTATAACGTGCTTT |
| 3.09 | CCTCGTTAGAATCAGAGCGGGAGCTAAACAGGAGGCCGATTAAAGGGATTTTAGACAGGA |
| 3.10 | ACGGTACGCCAGAATCCTGAGAAGTGTTTTTATAATCAGTGAGGCCACCGAGTAAAAGAG |
| 5.01 | CATTATCATTTTGCGGAACAAAGAAACCACCAGAAGGAGCGGAATTATCATCATATTCCT |
| 5.02 | GATTATCAGATGATGGCAATTCATCAATATAATCCTGATTGTTTGGATTATACTTCTGAA |
| 5.03 | TAATGGAAGGGTTAGAACCTACCATATCAAAATTATTTGCACGTAAAACAGAAATAAAGA |
| 5.04 | AATTGCGTAGATTTTCAGGTTTAACGTCAGATGAATATACAGTAACAGTACCTTTTACAT |
| 5.05 | CGGGAGAAACAATAACGGATTCGCCTGATTGCTTTGAATACCAAGTTACAAAATCGCGCA |
| 5.06 | GAGGCGAATTATTCATTTCAATTACCTGAGCAAAAGAAGATGATGAAACAAACATCAAGA |
| 5.07 | AAACAAAATTAATTACATTTAACAATTTCATTTGAATTACCTTTTTTAATGGAAACAGTA |
| 5.08 | CATAAATCAATATATGTGAGTGAATAACCTTGCTTCTGTAAATCGTCGCTATTAATTAAT |
| 5.09 | TTTCCCTTAGAATCCTTGAAAACATAGCGATAGCTTAGATTAAGACGCTGAGAAGAGTCA |
| 5.10 | ATAGTGAATTTATCAAAATCATAGGTCTGAGAGACTACCTTTTTAACCTCCGGCTTAGGT |
| Var 6 | TGGGTTATATAACTATATGTAAATGCTGATGCAAATCCAATCGCAAGACAAAGAACGCGA |
| 6.01 | GAAAACTTTTTCAAATATATTTTAGTTAATTTCATCTTCTGACCTAAATTTAATGGTTTG |
| 6.02 | AAATACCGACCGTGTGATAAATAAGGCGTTAAATAAGAATAAACACCGGAATCATAATTA |
| 6.03 | CTAGAAAAAGCCTGTTTAGTATCATATGCGTTATACAAATTCTTACCAGTATAAAGCCAA |
| 6.04 | CGCTCAACAGTAGGGCTTAATTGAGAATCGCCATATTTAACAACGCCAACATGTAATTTA |
| 6.05 | GGCAGAGGCATTTTCGAGCCAGTAATAAGAGAATATAAAGTACCGACAAAAGGTAAAGTA |
| 6.06 | ATTCTGTCCAGACGACGACAATAAACAACATGTTCAGCTAATGCAGAACGCGCCTGTTTA |
| 6.07 | TCAACAATAGATAAGTCCTGAACAAGAAAAATAATATCCCATCCTAATTTACGAGCATGT |
| 6.08 | AGAAACCAATCAATAATCGGCTGTCTTTCCTTATCATTCCAAGAACGGGTATTAAACCAA |
| 6.09 | GTACCGCACTCATCGAGAACAAGCAAGCCGTTTTTATTTTCATCGTAGGAATCATTACCG |
| 6.10 | CGCCCAATAGCAAGCAAATCAGATATAGAAGGCTTATCCGGTATTCTAAGAACGCGAGGC |
| Var 7 | GTTTTAGCGAACCTCCCGACTTGCGGGAGGTTTTGAAGCCTTAAATCAAGATTAGTTGCT |
| 7.01 | ATTTTGCACCCAGCTACAATTTTATCCTGAATCTTACCAACGCTAACGAGCGTCTTTCCA |
| 7.02 | GAGCCTAATTTGCCAGTTACAAAATAAACAGCCATATTATTTATCCCAATCCAAATAAGA |
| 7.03 | AACGATTTTTTGTTTAACGTCAAAAATGAAAATAGCAGCCTTTACAGAGAGAATAACATA |
| 7.04 | AAAACAGGGAAGCGCATTAGACGGGAGAATTAACTGAACACCCTGAACAAAGTCAGAGGG |
| 7.05 | TAATTGAGCGCTAATATCAGAGAGATAACCCACAAGAATTGAGTTAAGCCCAATAATAAG |
| 7.06 | AGCAAGAAACAATGAAATAGCAATAGCTATCTTACCGAAGCCCTTTTTAAGAAAAGTAAG |
| 7.07 | CAGATAGCCGAACAAAGTTACCAGAAGGAAACCGAGGAAACGCAATAATAACGGAATACC |
| 7.08 | CAAAAGAACTGGCATGATTAAGACTCCTTATTACGCAGTATGTTAGCAAACGTAGAAAAT |
| 7.09 | ACATACATAAAGGTGGCAACATATAAAAGAAACGCAAAGACACCACGGAATAAGTTTATT |
| 7.10 | TTGTCACAATCAATAGAAAATTCATATGGTTTACCAGCGCCAAAGACAAAAGGGCGACAT |
| 9.01 | CCAGGCGGATAAGTGCCGTCGAGAGGGTTGATATAAGTATAGCCCGGAATAGGTGTATCA |
| 9.02 | CCGTACTCAGGAGGTTTAGTACCGCCACCCTCAGAACCGCCACCCTCAGAACCGCCACCC |
| 9.03 | TCAGAGCCACCACCCTCATTTTCAGGGATAGCAAGCCCAATAGGAACCCATGTACCGTAA |
| 9.04 | CACTGAGTTTCGTCACCAGTACAAACTACAACGCCTGTAGCATTCCACAGACAGCCCTCA |
| 9.05 | TAGTTAGCGTAACGATCTAAAGTTTTGTCGTCTTTCCAGACGTTAGTAAATGAATTTTCT |
| 9.06 | GTATGGGATTTTGCTAAACAACTTTCAACAGTTTCAGCGGAGTGAGAATAGAAAGGAACA |
| 9.07 | ACTAAAGGAATTGCGAATAATAATTTTTTCACGTTGAAAATCTCCAAAAAAAAGGCTCCA |
| 9.08 | AAAGGAGCCTTTAATTGTATCGGTTTATCAGCTTGCTTTCGAGGTGAATTTCTTAAACAG |
| 9.09 | CTTGATACCGATAGTTGCGCCGACAATGACAACAACCATCGCCCACGCATAACCGATATA |
| 9.10 | TTCGGTCGCTGAGGCTTGCAGGGAGTTAAAGGCCGCTTTTGCGGGATCGTCACCCTCAGC |
| Var 10 | AGCGAAAGACAGCATCGGAACGAGGGTAGCAACGGCTACAGAGGCTTTGAGGACTAAAGA |
| 10.01 | CTTTTTCATGAGGAAGTTTCCATTAAACGGGTAAAATACGTAATGCCACTACGAAGGCAC |
| 10.02 | CAACCTAAAACGAAAGAGGCAAAAGAATACACTAAAACACTCATCTTTGACCCCCAGCGA |
| 10.03 | TTATACCAAGCGCGAAACAAAGTACAACGGAGATTTGTATCATCGCCTGATAAATTGTGT |
| 10.04 | CGAAATCCGCGACCTGCTCCATGTTACTTAGCCGGAACGAGGCGCAGACGGTCAATCATA |
| 10.05 | AGGGAACCGAACTGACCAACTTTGAAAGAGGACAGATGAACGGTGTACAGACCAGGCGCA |
| 10.06 | TAGGCTGGCTGACCTTCATCAAGAGTAATCTTGACAAGAACCGGATATTCATTACCCAAA |
| 10.07 | TCAACGTAACAAAGCTGCTCATTCAGTGAATAAGGCTTGCCCTGACGAGAAACACCAGAA |
| 10.08 | CGAGTAGTAAATTGGGCTTGAGATGGTTTAATTTCAACTTTAATCATTGTGAATTACCTT |
| 10.09 | ATGCGATTTTAAGAACTGGCTCATTATACCAGTCAGGACGTTGGGAAGAAAAATCTACGT |
| 10.10 | TAATAAAACGAACTAACGGAACAACATTATTACAGGTAGAAAGATTCATCAGTTGAGATT |
| Var 11 | TAGGAATACCACATTCAACTAATGCAGATACATAACGCCAAAAGGAATTACGAGGCATAG |
| 11.01 | TAAGAGCAACACTATCATAACCCTCGTTTACCAGACGACGATAAAAACCAAAATAGCGAG |
| 11.02 | AGGCTTTTGCAAAAGAAGTTTTGCCAGAGGGGGTAATAGTAAAATGTTTAGACTGGATAG |
| 11.03 | CGTCCAATACTGCGGAATCGTCATAAATATTCATTGAATCCCCCTCAAATGCTTTAAACA |
| 11.04 | GTTCAGAAAACGAGAATGACCATAAATCAAAAATCAGGTCTTTACCCTGACTATTATAGT |
| 11.05 | CAGAAGCAAAGCGGATTGCATCAAAAAGATTAAGAGGAAGCCCGAAAGACTTCAAATATC |
| 11.06 | GCGTTTTAATTCGAGCTTCAAAGCGAACCAGACCGGAAGCAAACTCCAACAGGTCAGGAT |
| 11.07 | TAGAGAGTACCTTTAATTGCTCCTTTTGATAAGAGGTCATTTTTGCGGATGGCTTAGAGC |
| 11.08 | TTAATTGCTGAATATAATGCTGTAGCTCAACATGTTTTAAATATGCAACTAAAGTACGGT |
| 11.09 | GTCTGGAAGTTTCATTCCATATAACAGTTGATTCCCAATTCTGCGAACGAGTAGATTTAG |
| 11.10 | TTTGACCATTAGATACATTTCGCAAATGGTCAATAACCTGTTTAGCTAT |
| Var 12 | ATTTTCATTTGGGGCGCGAGCTGAAAAGGTGGCATCAATTCTACTAATAGTAGTAGCATT |
| **Additional tiling oligos for 5 target nanoswitch (added to base set)** | |
| 4.025 | TGGAAATACCTACATTTTGACGCTCAATCG |
| 4.03 | TCTGAAATGGATTATTTACATTGGCAGATTCACCAGTCACACGACCAGTAATAAAAGGGA |
| 4.04 | CATTCTGGCCAACAGAGATAGAACCCTTCTGACCTGAAAGCGTAAGAATACGTGGCACAG |
| 4.05 | ACAATATTTTTGAATGGCTATTAGTCTTTAATGCGCGAACTGATAGCCCTAAAACATCGC |
| 4.06 | CATTAAAAATACCGAACGAACCACCAGCAGAAGATAAAACAGAGGTGAGGCGGTCAGTAT |
| 4.07 | TAACACCGCCTGCAACAGTGCCACGCTGAGAGCCAGCAGCAAATGAAAAATCTAAAGCAT |
| 4.08 | CACCTTGCTGAACCTCAAATATCAAACCCTCAATCAATATCTGGTCAGTTGGCAAATCAA |
| 4.09 | CAGTTGAAAGGAATTGAGGAAGGTTATCTAAAATATCTTTAGGAGCACTAACAACTAATA |
| 4.1 | GATTAGAGCCGTCAATAGATAATACATTTGAGGATTTAGAAGTATTAGACTTTACAAACA |
| Var5 | ATTCGACAACTCGTATTAAATCCTTTGCCCGAACGTTATTAATTTTAAAAGTTTGAGTAA |
| 8.025 | ATGAAACCATCGATAGCAGCACCGTAATCA |
| 8.03 | GTAGCGACAGAATCAAGTTTGCCTTTAGCGTCAGACTGTAGCGCGTTTTCATCGGCATTT |
| 8.04 | TCGGTCATAGCCCCCTTATTAGCGTTTGCCATCTTTTCATAATCAAAATCACCGGAACCA |
| 8.05 | GAGCCACCACCGGAACCGCCTCCCTCAGAGCCGCCACCCTCAGAACCGCCACCCTCAGAG |
| 8.06 | CCACCACCCTCAGAGCCGCCACCAGAACCACCACCAGAGCCGCCGCCAGCATTGACAGGA |
| 8.07 | GGTTGAGGCAGGTCAGACGATTGGCCTTGATATTCACAAACAAATAAATCCTCATTAAAG |
| 8.08 | CCAGAATGGAAAGCGCAGTCTCTGAATTTACCGTTCCAGTAAGCGTCATACATGGCTTTT |
| 8.09 | GATGATACAGGAGTGTACTGGTAATAAGTTTTAACGGGGTCAGTGCCTTGAGTAACAGTG |
| 8.1 | CCCGTATAAACAGTTAATGCCCCCTGCCTATTTCGGAACCTATTATTCTGAAACATGAAA |
| Var9 | GTATTAAGAGGCTGAGACTCCTCAAGAGAAGGATTAGGATTAGCGGGGTTTTGCTCAGTA |
| **Additional tiling oligos for single target nanoswitch (added to base set)** | |
| Var4.5 | CAATACTTCTTTGATTAGTAATAACATCAC |
| 4.01 | TTGCCTGAGTAGAAGAACTCAAACTATCGGCCTTGCTGGTAATATCCAGAACAATATTAC |
| 4.02 | CGCCAGCCATTGCAACAGGAAAAACGCTCATGGAAATACCTACATTTTGACGCTCAATCG |
| 4.03 | TCTGAAATGGATTATTTACATTGGCAGATTCACCAGTCACACGACCAGTAATAAAAGGGA |
| 4.04 | CATTCTGGCCAACAGAGATAGAACCCTTCTGACCTGAAAGCGTAAGAATACGTGGCACAG |
| 4.05 | ACAATATTTTTGAATGGCTATTAGTCTTTAATGCGCGAACTGATAGCCCTAAAACATCGC |
| 4.06 | CATTAAAAATACCGAACGAACCACCAGCAGAAGATAAAACAGAGGTGAGGCGGTCAGTAT |
| 4.07 | TAACACCGCCTGCAACAGTGCCACGCTGAGAGCCAGCAGCAAATGAAAAATCTAAAGCAT |
| 4.08 | CACCTTGCTGAACCTCAAATATCAAACCCTCAATCAATATCTGGTCAGTTGGCAAATCAA |
| 4.09 | CAGTTGAAAGGAATTGAGGAAGGTTATCTAAAATATCTTTAGGAGCACTAACAACTAATA |
| 4.1 | GATTAGAGCCGTCAATAGATAATACATTTGAGGATTTAGAAGTATTAGACTTTACAAACA |
| Var5 | ATTCGACAACTCGTATTAAATCCTTTGCCCGAACGTTATTAATTTTAAAAGTTTGAGTAA |
| Var8.5 | TGACGGAAATTATTCATTAAAGGTGAATTA |
| 8.01 | TCACCGTCACCGACTTGAGCCATTTGGGAATTAGAGCCAGCAAAATCACCAGTAGCACCA |
| 8.02 | TTACCATTAGCAAGGCCGGAAACGTCACCAATGAAACCATCGATAGCAGCACCGTAATCA |
| 8.03 | GTAGCGACAGAATCAAGTTTGCCTTTAGCGTCAGACTGTAGCGCGTTTTCATCGGCATTT |
| 8.04 | TCGGTCATAGCCCCCTTATTAGCGTTTGCCATCTTTTCATAATCAAAATCACCGGAACCA |
| 8.05 | GAGCCACCACCGGAACCGCCTCCCTCAGAGCCGCCACCCTCAGAACCGCCACCCTCAGAG |
| 8.06 | CCACCACCCTCAGAGCCGCCACCAGAACCACCACCAGAGCCGCCGCCAGCATTGACAGGA |
| 8.07 | GGTTGAGGCAGGTCAGACGATTGGCCTTGATATTCACAAACAAATAAATCCTCATTAAAG |
| 8.08 | CCAGAATGGAAAGCGCAGTCTCTGAATTTACCGTTCCAGTAAGCGTCATACATGGCTTTT |
| 8.09 | GATGATACAGGAGTGTACTGGTAATAAGTTTTAACGGGGTCAGTGCCTTGAGTAACAGTG |
| 8.1 | CCCGTATAAACAGTTAATGCCCCCTGCCTATTTCGGAACCTATTATTCTGAAACATGAAA |
| Var9 | GTATTAAGAGGCTGAGACTCCTCAAGAGAAGGATTAGGATTAGCGGGGTTTTGCTCAGTA |
